# Diagnostic accuracy of remote self-administered digital cognitive assessment by clinical diagnosis and Alzheimer’s disease PET imaging biomarkers

**DOI:** 10.64898/2026.09.20.26362296

**Authors:** Nikki H. Stricker, Elizabeth A. Boots, Rita Taylor, Winnie Z. Fan, Ryan D. Frank, Teresa J. Christianson, John L. Stricker, Walter K. Kremers, Mary M. Machulda, Paula A. Aduen, John A. Lucas, Morgan A. Hughes, Aimee J. Karstens, Gregory S. Day, Christian Lachner, Neill R. Graff-Radford, Jason Hassenstab, Michelle M. Mielke, Clifford R. Jack, Bradley F. Boeve, Jonathan Graff-Radford, Ronald C. Petersen

## Abstract

**INTRODUCTION:** Cognitive characterization is increasingly important for Alzheimer’s disease treatment decisions, highlighting the need for scalable cognitive assessment. We evaluated the diagnostic accuracy of a remote self-administered cognitive screening battery, Mayo Test Drive (MTD), across clinically and biologically defined groups.

**METHODS:** In this prospectively designed study, 2,131 cognitively unimpaired (CU) and 118 cognitively impaired participants, including 96 with mild cognitive impairment and 22 with mild dementia (MCI/DEM), were recruited from observational cohort studies and completed MTD remotely (mean age=70 years, SD=12; 48% male; mean education=16 years, SD=2). Clinical diagnoses were determined by multidisciplinary consensus independently of MTD. Amyloid- and tau-PET were available for 1,224 participants. Logistic regression estimated associations between MTD and CU versus MCI/DEM, CU A-versus MCI/DEM A+, and A-T-versus A+T+. MTD diagnostic accuracy was compared with person-administered cognitive measures. Sensitivity and specificity across score types and comparison groups were examined.

**RESULTS:** MTD showed excellent discrimination of CU versus MCI/DEM (AUC=.89; 95% CI=.87-.92), and discriminability remained high for CU versus MCI (AUC=.88; 95% CI=.85-.92). Discriminability was highest when comparing CU A- and MCI/DEM A+ groups (AUC=.95; 95% CI=.92-.97). MTD outperformed estimated MMSE (p=.02) and was comparable to Mayo-PACC (p=.71) for A-T-versus A+T+ discrimination (AUC=.76; 95% CI=.71-.81). Combining both raw score and normative score cutoffs helped balance sensitivity and specificity relative to use of either cutoff in isolation. A two-stage interpretation framework further improved sensitivity while maintaining clinically useful specificity and informed development of an interpretation guide.

**DISCUSSION:** A remote self-administered digital cognitive assessment showed strong diagnostic accuracy for detecting MCI and mild dementia in a large, primarily population-based cohort. The remote, self-administered MTD composite also showed comparable performance to established in-person cognitive composites for differentiating amyloid and tau-defined biomarker groups. Remote self-administered digital cognitive assessment may complement existing cognitive evaluation approaches and support scalable cognitive characterization.

**Highlights:**

- Remote self-administered cognitive assessment showed strong differentiation of cognitively unimpaired and cognitively impaired participants.
- Diagnostic accuracy improved when amyloid PET biomarker status was incorporated.
- Remote assessment demonstrated sensitivity to amyloid and tau PET-defined Alzheimer’s disease.
- Performance was comparable to established in-person composites and better than an in-person mental status screening measure.
- Digital cognitive screening may support scalable treatment and clinical trial workflows.

## Introduction

The emergence of monoclonal antibody treatments that target Alzheimer’s disease (AD) pathology has altered the clinical landscape, shifting the focus toward earlier identification of symptomatic patients to ensure timely access to disease-modifying interventions [1, 2]. This represents a paradigm shift, as there has previously been uncertainty regarding whether routine cognitive screening is necessary in the absence of available treatments for neurodegenerative disorders [3]. Accurate and reliable determination of cognitive status is a required component of clinical decision-making, with cognitive characterization and biomarker evidence of AD pathology explicitly required by monoclonal antibody treatment pathways. Currently, a diagnosis of mild cognitive impairment (MCI) or mild dementia is needed for treatment eligibility, although future treatment options may require confirmation of cognitively unimpaired (CU) status.

Comprehensive neuropsychological and neurological evaluations remain the gold standard for clinical characterization. However, access to these resources varies, remains limited in many clinical settings, and can be impacted by hesitance to order neuropsychological testing [4–7], creating challenges for timely assessment of individuals who may be candidates for treatment or clinical trial participation. Although cognitive screening measures alone are insufficient to diagnose MCI or dementia, scalable screening approaches may help inform decisions regarding downstream evaluation and improve access to cognitive characterization. Digital self-administered cognitive assessments that can be completed remotely or in clinic represent one approach to expanding access to cognitive characterization. To be clinically useful in the context of monoclonal antibody treatments, digital cognitive screening tools should demonstrate (1) diagnostic accuracy for identifying MCI and mild dementia, the clinical stages currently eligible for treatment, and (2) criterion validity for AD, including sensitivity to AD pathology across the clinical continuum.

Mayo Test Drive (MTD; Mayo Test Development through Rapid Iteration, Validation and Expansion, DRIVE) is a digital cognitive testing platform designed for remote self-administered assessment across multiple device types (smartphone, tablet, computer). The MTD screening battery requires approximately 15-20 minutes to complete and has previously established feasibility, reliability, validity and remote normative data [8–13]. The MTD screening battery includes a computer-adaptive measure of memory and a measure of processing speed/executive function requiring visuospatial matching. These measures are combined into a composite that has demonstrated strong associations with traditional in-person cognitive composites and AD biomarkers [9].

The present study evaluated the cross-sectional diagnostic accuracy of MTD across clinically and biologically defined groups relevant to AD clinical care and research in a large population-based cohort, with AD biomarkers available in more than half of participants. We examined the ability of MTD to identify early symptomatic AD by distinguishing (a) CU individuals from those with MCI or mild dementia, and (b) CU individuals without amyloid pathology (amyloid PET negative; A−) from individuals with MCI or mild dementia due to AD (amyloid PET positive; A+). MCI and mild dementia groups were combined for primary analyses to reflect detection of clinically meaningful cognitive impairment within the clinical stages currently considered for treatment; subgroup results are also reported. We also evaluated the criterion validity of the MTD composite by examining discrimination of individuals with normal amyloid and tau biomarkers (A−T−) from those with AD pathology (A+T+), independent of clinical syndrome, and assessed sensitivity to preclinical AD in CU individuals. To place performance in the context of established approaches to cognitive characterization, we directly compared MTD diagnostic accuracy with traditional person-administered cognitive measures for the A-T-versus A+T+ comparison. We additionally evaluated whether MTD was significantly associated with clinical and biomarker-defined group membership beyond demographic variables and examined sensitivity and specificity across score types and comparison groups to inform potential cut points and interpretation guidelines.

## Methods

### Participants and Study Design

Participants were recruited from observational cohort studies at Mayo Clinic, including the Mayo Clinic Study of Aging (MCSA) and the Mayo Clinic Alzheimer’s Disease Research Center (ADRC; Rochester, MN and Jacksonville, FL). Individuals completed clinical evaluations and neuroimaging as part of these parent studies and additionally participated in an ancillary remote digital cognitive assessment study. The MCSA is a population-based longitudinal study of aging in Olmsted County, Minnesota which recruits participants using age- and sex-stratified random sampling [14–16]. MCSA exclusion criteria are terminal illness or receiving hospice care. Participants without medical contraindications are offered enrollment in neuroimaging protocols. Amyloid PET imaging using Pittsburgh compound B (PiB) has been conducted since 2008, with tau PET imaging incorporated in 2016. All MCSA participants are invited to participate in this ancillary remote study unless they meet the additional exclusion criteria of the ancillary study (unable to read and speak English; unable to complete study activities). The ADRC recruits individuals with AD and related disorders as well as CU participants; ADRC recruitment for this ancillary remote digital cognitive assessment study focused on increasing sample representation of participants with cognitive impairment due to AD (Rochester, MN) and participants from groups not well represented in Olmsted County, including Black American participants (Jacksonville, FL).

This study was prospectively designed as a diagnostic accuracy investigation of remote digital cognitive assessment (see Supplemental Methods for details).

### Informed Consent

The study was conducted in accordance with the Declaration of Helsinki. Parent study procedures were approved by the Mayo Clinic and Olmsted Medical Center Institutional Review Boards. Written informed consent was obtained from participants for parent study protocols. Ancillary study procedures were approved by the Mayo Clinic Institutional Review Board. Consent for the ancillary MTD study was obtained orally or via electronic consent procedures. A detailed description of the ancillary study recruitment procedures is available [13, 17].

### Clinical Characterization

As part of the parent study protocols, participants underwent standardized clinical assessments including a physician examination, informant interview (Clinical Dementia Rating® scale)[18], and a multi-domain neuropsychological battery administered by a trained psychometrist. Diagnostic classification (CU, MCI, or dementia) was completed through multidisciplinary consensus using established criteria [14]. ADRC diagnostic determinations included consideration of prior diagnoses and biomarker data, when available. MCSA diagnostic determinations were based on in-person clinical, neurological, and neuropsychological data and were made without reference to prior diagnoses or biomarker status. All clinical diagnoses (ADRC and MCSA) were determined independently of the digital MTD cognitive data; MTD performance was not available to diagnostic raters and did not contribute to consensus diagnosis. Individuals with dementia severity greater than mild were excluded from the current analyses to focus on stages relevant to clinical screening and treatment eligibility.

### Neuroimaging Biomarkers

Amyloid and tau biomarker status was determined using Pittsburgh Compound-B (PiB) amyloid and flortaucipir tau positron emission tomography (PET) scans acquired within three years of the baseline MTD session. PET acquisition and processing methods have been previously described [19–22]. Amyloid positivity was defined using a Centiloid threshold of ≥ 25 [23], and tau positivity was defined using a global ROI SUVR threshold of ≥ 1.29 [24].

### Person-Administered Neuropsychological and Mental Status Screening Measures Completed in Clinic

Participants underwent in-person cognitive screening using either the Mini Mental Status Examination (MMSE) or the Short Test of Mental Status (STMS)[25, 26]. To equate cognitive screening measures across the full sample, STMS scores were converted to MMSE scores based on an unpublished conversion nomogram developed by Tang-Wai and colleagues as related to work comparing the STMS and MMSE in MCI [27]. Most participants also completed an in-person neuropsychological assessment comprised of nine neuropsychological tests, from which a Global Cognition z-score (Global-z) was derived. A subset of these tests was also used to calculate the Mayo Clinic Preclinical Alzheimer’s Cognitive Composite (Mayo-PACC). Further details regarding the neuropsychological test battery, composite calculations and reliability of these measures can be found elsewhere [9, 28].

### Remote Digital Cognitive Assessment

Participants completed MTD remotely at a self-selected time using a personalized link sent via email. The opportunity to complete MTD in clinic was often available upon request. Details about the MTD screening battery composite and subtest scores have been previously described [8–10]. Briefly, the battery includes two subtests: The Stricker Learning Span (SLS) is a novel word list memory test optimized for remote self-administration. The SLS employs a structured word list learning/recognition paradigm that emphasizes learning across repeated exposures following a computer-adaptive approach [10]. The Symbols Test (Symbols) is an open-source measure of visuospatial matching/discrimination, processing speed, and executive function adapted from the Ambulatory Research in Cognition application [29]. Symbols requires participants to identify which of two symbol pairs on the bottom of the screen matches one of three symbol pairs on the top of the screen; for MTD Symbols, there are four trials, with 12 items each [8]. An SLS delay follows the Symbols test. Participants also report test environment characteristics and potential distractions to support data quality evaluation. A total composite score is computed from both the SLS and Symbols (SLS Sum of Trials + Symbols Accuracy Weighted Score) and served as the primary MTD variable for analyses [8, 9]. Secondary variables included subtest scores [8, 9]. Demographically-adjusted T-scores were also used for select analyses [8]. MTD sessions were completed using MTD platforms v1 and v2. MTD test-retest reliability and direct reliability comparison to several of the traditional measures included in this study are reported in a recent study [12]; the sample overlaps with the one reported here, differing only based on the inclusion criteria applied to meet the aims of the separate manuscripts. There is also overlap of the current sample and that used to generate the normative data (N=1240) [8], but data collection for the current study represents approximately 1.5 additional years of participant accrual (normative data sample cutoff date was December 14, 2023) and there are 1,010 additional participants included here that were not in the normative sample. A subset of participants included in the current analyses have also been included in prior manuscripts addressing MTD criterion validity and biomarker associations. Prior publications examined continuous associations between MTD and neuroimaging biomarkers [9] or criterion validity of one MTD subtest (SLS) [10], whereas the present study evaluates diagnostic accuracy, clinical classification, sensitivity/specificity, and interpretation approaches in a substantially larger cohort.

### Inclusion Criteria

Participants from the MCSA and ADRC who completed a baseline MTD session between May 25, 2021 and June 15, 2025 were eligible for inclusion. Participants with evidence of invalid test performance (as previously described [13]) or CDR > 1 (i.e., moderate or severe dementia) were excluded. Subset analyses further required availability of amyloid and tau PET imaging within three years of MTD (for biomarker-defined groups) or availability of corresponding in-clinic neuropsychological testing (for direct comparisons of remote cognitive testing and traditional in-clinic cognitive testing).

### Statistical Methods

Patient characteristics were summarized using means and standard deviations for continuous variables and counts and percentages for categorical variables. Group comparisons were evaluated using t-tests or chi-square tests. Effect sizes were estimated using Hedge’s g.

Logistic regression models were used to examine associations between cognitive measures and group membership for the clinically defined and biomarker-defined groups. Models with and without age, sex, and education adjustment were performed to assess the incremental contribution of cognitive performance beyond demographic variables. Odds ratios and area under the receiver operating characteristic curves (AUC) were estimated with 95% confidence intervals. We hypothesized *a priori* that MTD would demonstrate excellent discrimination (AUC > 0.85) between CU and MCI/mild dementia participants (also see Table S1 and Supplemental Methods).

We also hypothesized that the odds ratio for MTD composite would be significantly associated with clinical and biomarker group membership after age/sex/education adjustment. Differences in AUCs between non-nested models were evaluated using concordance-based methods using the R survival package for select comparisons [30] and were used to test hypotheses directly comparing remote MTD with traditional person-administered measures.

Sensitivity, specificity, and corresponding confidence intervals were calculated across a range of empirically derived and clinically informed cutoffs. *A priori* clinically informed cutoffs of -1 SD were selected, consistent with our prior work [8], though data for multiple cutoffs are provided to characterize sensitivity and specificity across a range of commonly employed clinical cutoffs. Optimal cutoffs were identified using Youden’s index and informed proposed cutoffs, with *a priori* prioritization of standard clinical cutoffs (<-1 SD). A three-range classification framework was also developed; sensitivity and specificity were calculated using confusion matrices that retained all participants to avoid potentially biased estimates of diagnostic performance, consistent with FDA guidance regarding evaluation of diagnostic tests [31]. One-stage sensitivity was defined as true positive / (true positive + indeterminate positive + false negative) [TP / (TP + IP + FN)] and one-stage specificity was defined as true negative / (true negative + indeterminate negative + false positive) [TN / (TN + IN + FP)], whereas two-stage sensitivity was defined as (TP + IP) / (TP + IP + FN) and two-stage specificity was defined as (TN + IN) / (TN + IN + FP). For two-stage sensitivity and specificity, indeterminate cases represent individuals with test scores falling within a defined indeterminate range who would be identified as positive or negative through additional evaluation. For individuals with indeterminate MTD results in the current analyses, underlying positive or negative status was determined using the reference standard for the specific comparison (research diagnosis, PET imaging biomarkers, or both).

This study is reported in accordance with Standards for Reporting of Diagnostic Accuracy Studies (STARD); dementia-specific reporting considerations were addressed in accordance with the STARDdem Initiative [32, 33]. All analyses were conducted using R version 4.4.1, with two-sided p-values <0.05 considered statistically significant.

## Results

### Participant Characteristics

2,288 participants completed a baseline MTD session. Of these, 39 were excluded due to missing MTD composite data (n=29), dementia severity greater than mild (CDR>1, n=3), test validity concerns (n=5; [13]), or missing demographics (n=2) (see Figure S1). Excluded participants were more likely to have cognitive impairment (*p*<.001), and the exclusion of individuals with moderate dementia severity may have contributed to this difference. Excluded participants were also older (*p*<.001); no differences were observed in sex, education, race, ethnicity, or amyloid or tau PET status (*p*’s>.05).

The final analytic sample included 2,249 participants (Table 1). Most were CU (95%), with 4% classified as MCI and 1% as mild dementia. Most participants were non-Hispanic White (93%), college educated (mean years of education = 15.6, SD=2.3) and approximately half were male (48%). MTD was completed remotely by almost all participants (>99%). Amyloid and tau PET imaging data within three years of the MTD session were available in 54% of participants. Participants with available PET imaging were less likely to be Hispanic (*p*=.045), less likely to report English as their primary language (*p*=.004), and older than those without imaging (*p*=.003) but did not differ by sex, education, or race (*p*’s>.05; Table S2). Differences reflect parent study design, which offered imaging more frequently to older participants and to those with cognitive impairment. Among participants with amyloid PET data available, 27% of CU participants were A+ and 64% of MCI/DEM participants were A+. Most participants with cognitive impairment recruited from the ADRC with amyloid PET data available were A+ (85% MCI A+, 100% mild dementia A+), whereas those recruited from the MCSA showed lower A+ rates (42% MCI A+, 50% mild dementia A+), aligning with the population-based sampling design of the MCSA.

**Table 1.**
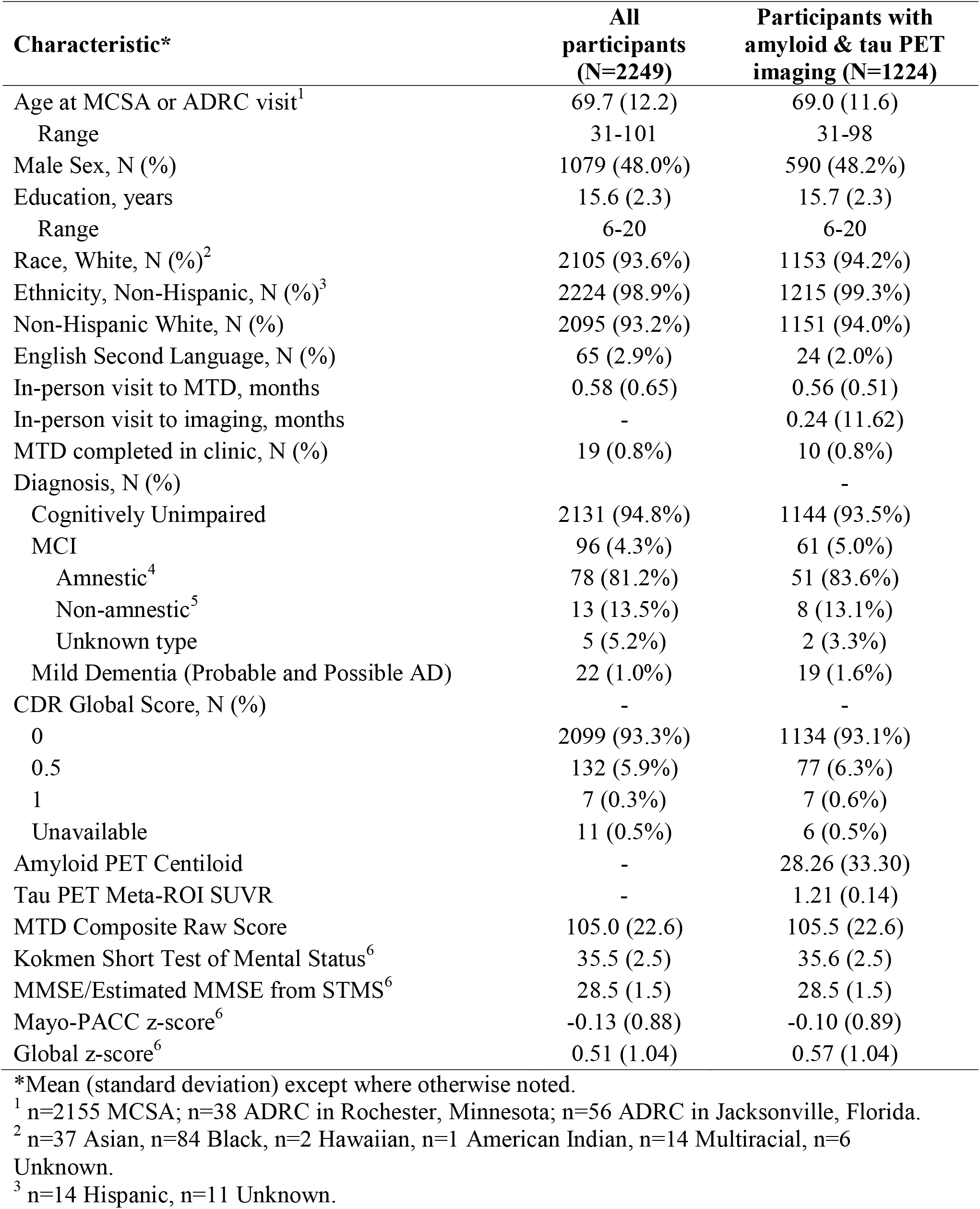
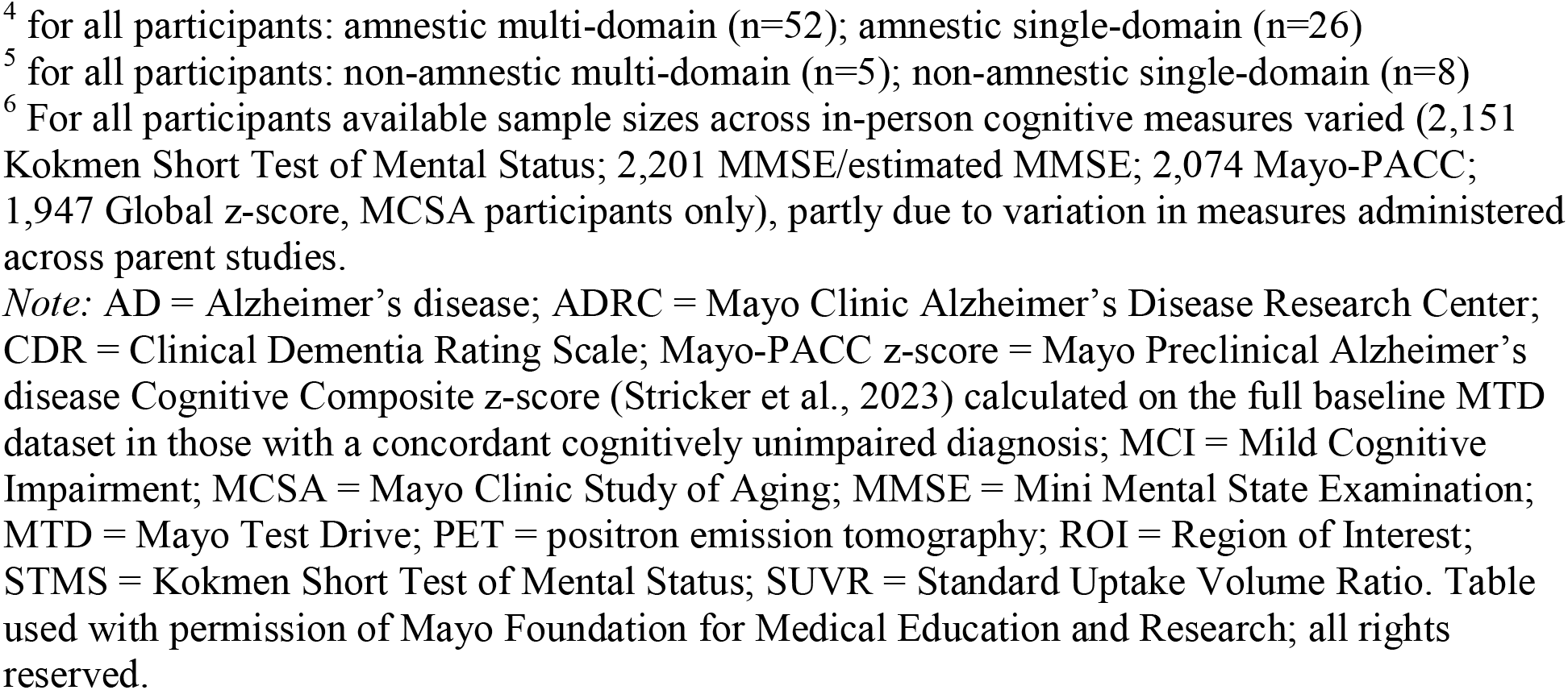
Demographic, clinical, and neuroimaging characteristics for all participants and for the participant subset with available amyloid and tau PET imaging.

| **Characteristic\*** | **All participants (N=2249)** | **Participants with amyloid & tau PET imaging (N=1224)** |
| --- | --- | --- |
| Age at MCSA or ADRC visit<sup>1</sup> | 69.7 (12.2) | 69.0 (11.6) |
| Range | 31-101 | 31-98 |
| Male Sex, N (%) | 1079 (48.0%) | 590 (48.2%) |
| Education, years | 15.6 (2.3) | 15.7 (2.3) |
| Range | 6-20 | 6-20 |
| Race, White, N (%)<sup>2</sup> | 2105 (93.6%) | 1153 (94.2%) |
| Ethnicity, Non-Hispanic, N (%)<sup>3</sup> | 2224 (98.9%) | 1215 (99.3%) |
| Non-Hispanic White, N (%) | 2095 (93.2%) | 1151 (94.0%) |
| English Second Language, N (%) | 65 (2.9%) | 24 (2.0%) |
| In-person visit to MTD, months | 0.58 (0.65) | 0.56 (0.51) |
| In-person visit to imaging, months | - | 0.24 (11.62) |
| MTD completed in clinic, N (%) | 19 (0.8%) | 10 (0.8%) |
| Diagnosis, N (%) |  | - |
| Cognitively Unimpaired | 2131 (94.8%) | 1144 (93.5%) |
| MCI | 96 (4.3%) | 61 (5.0%) |
| Amnestic<sup>4</sup> | 78 (81.2%) | 51 (83.6%) |
| Non-amnestic<sup>5</sup> | 13 (13.5%) | 8 (13.1%) |
| Unknown type | 5 (5.2%) | 2 (3.3%) |
| Mild Dementia (Probable and Possible AD) | 22 (1.0%) | 19 (1.6%) |
| CDR Global Score, N (%) | - | - |
| 0 | 2099 (93.3%) | 1134 (93.1%) |
| 0.5 | 132 (5.9%) | 77 (6.3%) |
| 1 | 7 (0.3%) | 7 (0.6%) |
| Unavailable | 11 (0.5%) | 6 (0.5%) |
| Amyloid PET Centiloid | - | 28.26 (33.30) |
| Tau PET Meta-ROI SUVR | - | 1.21 (0.14) |
| MTD Composite Raw Score | 105.0 (22.6) | 105.5 (22.6) |
| Kokmen Short Test of Mental Status<sup>6</sup> | 35.5 (2.5) | 35.6 (2.5) |
| MMSE/Estimated MMSE from STMS<sup>6</sup> | 28.5 (1.5) | 28.5 (1.5) |
| Mayo-PACC z-score<sup>6</sup> | -0.13 (0.88) | -0.10 (0.89) |
| Global z-score<sup>6</sup> | 0.51 (1.04) | 0.57 (1.04) |

\*Mean (standard deviation) except where otherwise noted.
<sup>1</sup> n=2155 MCSA; n=38 ADRC in Rochester, Minnesota; n=56 ADRC in Jacksonville, Florida.
<sup>2</sup> n=37 Asian, n=84 Black, n=2 Hawaiian, n=1 American Indian, n=14 Multiracial, n=6 Unknown.
<sup>3</sup> n=14 Hispanic, n=11 Unknown.
<sup>4</sup> for all participants: amnesic multi-domain (n=52); amnesic single-domain (n=26)
<sup>5</sup> for all participants: non-amnesic multi-domain (n=5); non-amnesic single-domain (n=8)
<sup>6</sup> For all participants available sample sizes across in-person cognitive measures varied (2,151 Kokmen Short Test of Mental Status; 2,201 MMSE/estimated MMSE; 2,074 Mayo-PACC; 1,947 Global z-score, MCSA participants only), partly due to variation in measures administered across parent studies.
*Note:* AD = Alzheimer's disease; ADRC = Mayo Clinic Alzheimer's Disease Research Center; CDR = Clinical Dementia Rating Scale; Mayo-PACC z-score = Mayo Preclinical Alzheimer's disease Cognitive Composite z-score (Stricker et al., 2023) calculated on the full baseline MTD dataset in those with a concordant cognitively unimpaired diagnosis; MCI = Mild Cognitive Impairment; MCSA = Mayo Clinic Study of Aging; MMSE = Mini Mental State Examination; MTD = Mayo Test Drive; PET = positron emission tomography; ROI = Region of Interest; STMS = Kokmen Short Test of Mental Status; SUVR = Standard Uptake Volume Ratio. Table used with permission of Mayo Foundation for Medical Education and Research; all rights reserved.

### Clinical Characterization

#### Clinical diagnosis

##### CU vs MCI/mild dementia

Unadjusted MTD composite demonstrated excellent discrimination between CU participants and those with MCI or mild dementia (AUC = 0.89; see Table 2 for 95% CIs). This AUC was significantly higher compared to a demographics-only model (AUC=0.69; *p*< .001; Figure 1). MTD subtests also showed good discrimination between CU and MCI/mild dementia participants, with AUC values > 0.85 for SLS variables and > .80 for Symbols Test variables (Figure 1; Figure S2). A higher MTD composite score was associated with significantly lower odds of MCI/mild dementia in unadjusted models (Odds Ratio=0.93, 95% CI=0.92-0.94, *p*<.001; Table S3). Each one standard deviation increase in MTD composite (using 1 SD of the sample, or a 23-point increase) is associated with 81% decreased odds of MCI/DEM (95% CI 76-85%). In models adjusted for demographics, MTD composite significantly contributed to classification beyond demographics (*p*<.001; Tables 2 and S3).

**Figure 1.**
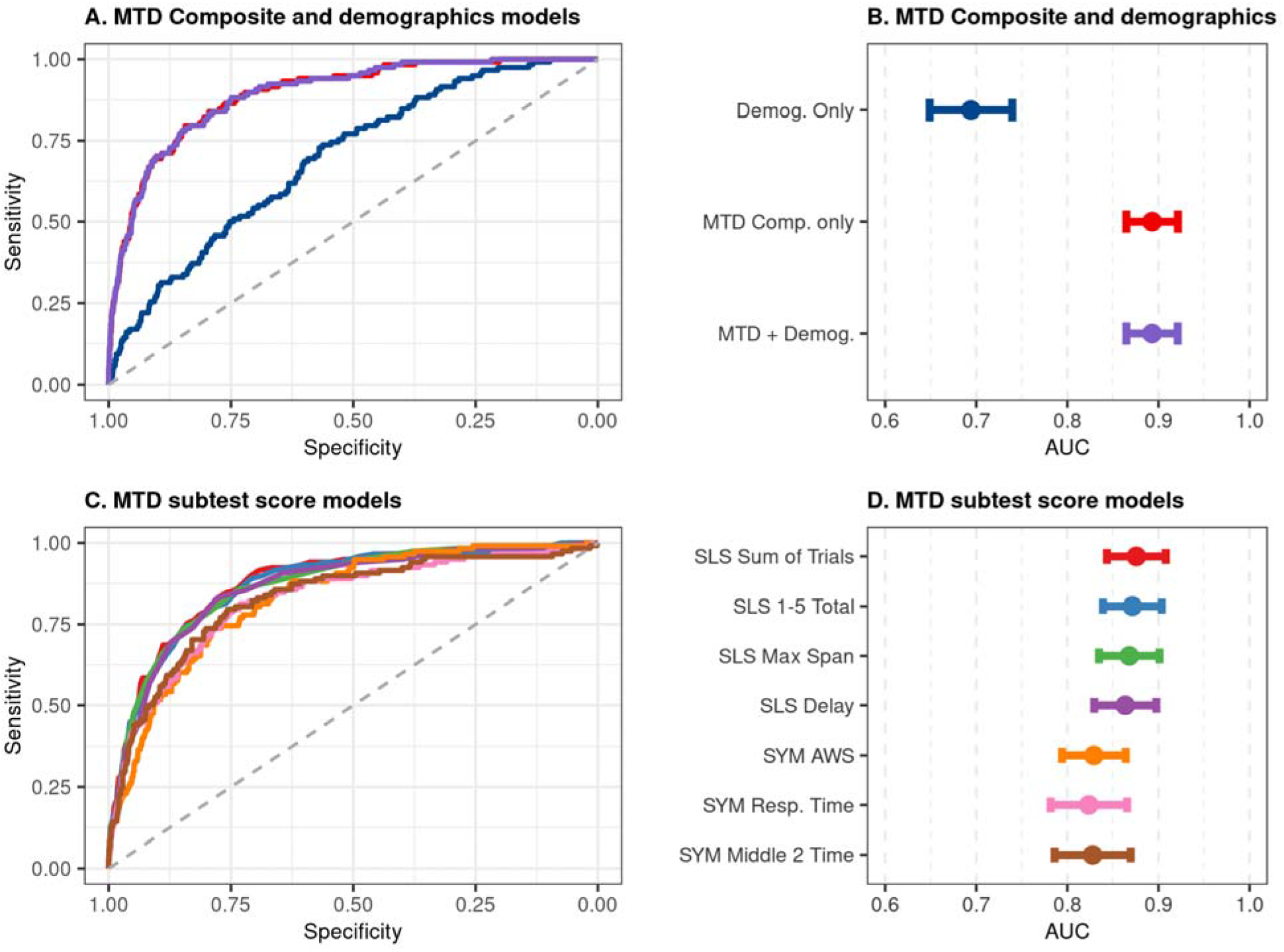
Model performance for predicting clinical diagnosis: cognitively unimpaired (n=2131) versus cognitively impaired (n=118 with MCI or mild dementia). *Note*. Panels A and B show the AUCs for the MTD Composite, demographics only, and MTD Composite plus demographics for differentiating cognitively unimpaired versus cognitively impaired groups. The cognitive impairment group includes 81% (n=96) with mild cognitive impairment (MCI) and 19% (n=22) with mild dementia. Panels C and D show the AUCs for the MTD subtests for differentiating cognitively unimpaired versus cognitively impaired groups. AUC = area under the receiver operating characteristic curve; AWS = Accuracy Weighted Score; MTD = Mayo Test Drive; SLS = Stricker Learning Span; SLS 1-5 Total = total words correctly recognized across all 5 learning trials; SLS Delay = number of words correctly recognized during the delay trial; SLS Max Span = maximum number of words correctly recognized across any of the 5 learning trials; SLS Sum of Trials = SLS Trials 1-5 Total Correct + SLS Delay Correct; SYM = Symbols Test; SYM Resp. Time = Symbols Average Response Time (seconds) for Correct Items across all 4 Symbols trials; SYM Middle 2 Time = average seconds to complete a trial when averaged across the middle two trials, excluding highest and lowest performances. Figure used with permission of Mayo Foundation for Medical Education and Research; all rights reserved.

**Table 2.**
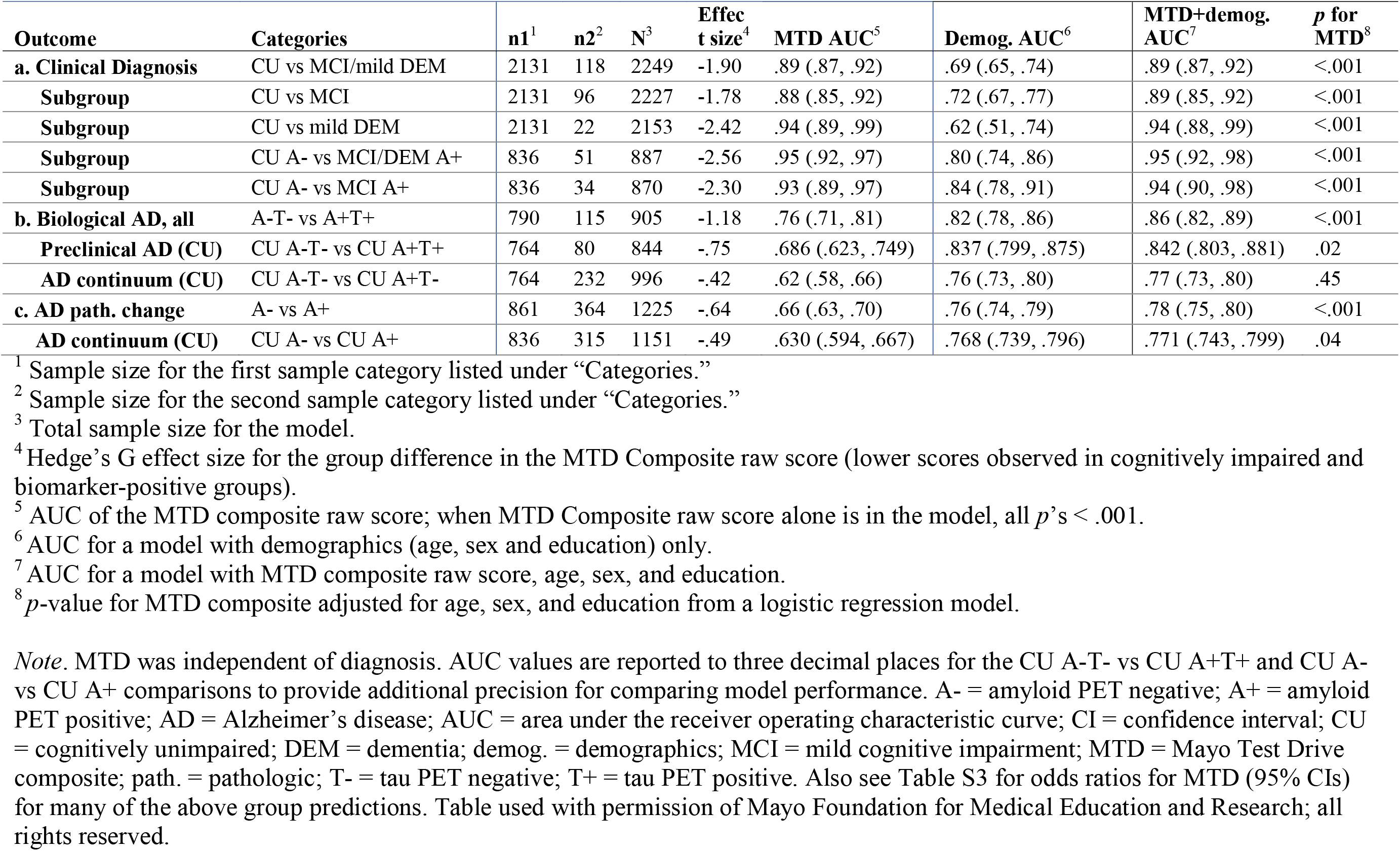
Subgroup sample sizes, group difference Hedge’s G effect sizes, and area under the receiving operating characteristic curves (AUC) with 95% CI for group comparisons for MTD Composite raw score alone, demographics (age, sex and education) alone, and MTD Composite raw score and demographics combined.

| Outcome | Categories | n1<sup>1</sup> | n2<sup>2</sup> | N<sup>3</sup> | Effect size<sup>4</sup> | MTD AUC<sup>5</sup> | Demog. AUC<sup>6</sup> | MTD+demog. AUC<sup>7</sup> | p for MTD<sup>8</sup> |
| --- | --- | --- | --- | --- | --- | --- | --- | --- | --- |
| **a. Clinical Diagnosis** | CU vs MCI/mild DEM | 2131 | 118 | 2249 | -1.90 | .89 (.87, .92) | .69 (.65, .74) | .89 (.87, .92) | <.001 |
| Subgroup | CU vs MCI | 2131 | 96 | 2227 | -1.78 | .88 (.85, .92) | .72 (.67, .77) | .89 (.85, .92) | <.001 |
| Subgroup | CU vs mild DEM | 2131 | 22 | 2153 | -2.42 | .94 (.89, .99) | .62 (.51, .74) | .94 (.88, .99) | <.001 |
| Subgroup | CU A- vs MCI/DEM A+ | 836 | 51 | 887 | -2.56 | .95 (.92, .97) | .80 (.74, .86) | .95 (.92, .98) | <.001 |
| Subgroup | CU A- vs MCI A+ | 836 | 34 | 870 | -2.30 | .93 (.89, .97) | .84 (.78, .91) | .94 (.90, .98) | <.001 |
| **b. Biological AD, all** | A-T- vs A+T+ | 790 | 115 | 905 | -1.18 | .76 (.71, .81) | .82 (.78, .86) | .86 (.82, .89) | <.001 |
| Preclinical AD (CU) | CU A-T- vs CU A+T+ | 764 | 80 | 844 | -.75 | .686 (.623, .749) | .837 (.799, .875) | .842 (.803, .881) | .02 |
| AD continuum (CU) | CU A-T- vs CU A+T- | 764 | 232 | 996 | -.42 | .62 (.58, .66) | .76 (.73, .80) | .77 (.73, .80) | .45 |
| **c. AD path. change** | A- vs A+ | 861 | 364 | 1225 | -.64 | .66 (.63, .70) | .76 (.74, .79) | .78 (.75, .80) | <.001 |
| AD continuum (CU) | CU A- vs CU A+ | 836 | 315 | 1151 | -.49 | .630 (.594, .667) | .768 (.739, .796) | .771 (.743, .799) | .04 |

<sup>1</sup> Sample size for the first sample category listed under “Categories.”
<sup>2</sup> Sample size for the second sample category listed under “Categories.”
<sup>3</sup> Total sample size for the model.
<sup>4</sup> Hedge’s G effect size for the group difference in the MTD Composite raw score (lower scores observed in cognitively impaired and biomarker-positive groups).
<sup>5</sup> AUC of the MTD composite raw score; when MTD Composite raw score alone is in the model, all $p$ ’s < .001.
<sup>6</sup> AUC for a model with demographics (age, sex and education) only.
<sup>7</sup> AUC for a model with MTD composite raw score, age, sex, and education.
<sup>8</sup> $p$ -value for MTD composite adjusted for age, sex, and education from a logistic regression model.
*Note.* MTD was independent of diagnosis. AUC values are reported to three decimal places for the CU A-T- vs CU A+T+ and CU A- vs CU A+ comparisons to provide additional precision for comparing model performance. A- = amyloid PET negative; A+ = amyloid PET positive; AD = Alzheimer’s disease; AUC = area under the receiver operating characteristic curve; CI = confidence interval; CU = cognitively unimpaired; DEM = dementia; demog. = demographics; MCI = mild cognitive impairment; MTD = Mayo Test Drive composite; path. = pathologic; T- = tau PET negative; T+ = tau PET positive. Also see Table S3 for odds ratios for MTD (95% CIs) for many of the above group predictions. Table used with permission of Mayo Foundation for Medical Education and Research; all rights reserved.

##### CU vs MCI

Discriminatory performance of the unadjusted MTD composite remained high for CU versus MCI (AUC=.88; Table 2), excluding participants with mild dementia from the comparison.

##### CU vs mild dementia

Discriminatory performance of the unadjusted MTD composite was excellent for CU versus mild dementia (AUC=.94; Table 2), excluding participants with MCI from the comparison.

##### MCI vs. mild dementia

Discriminatory performance of the unadjusted MTD composite for MCI versus mild dementia was significant but modest (AUC=.66; 95% CI=.52-.79).

#### Clinical diagnosis refined by amyloid status

##### CU A- vs MCI/DEM A+

Refining clinical diagnostic groups by amyloid PET status improved the discriminatory performance of the MTD composite. The MTD composite yielded the highest accuracy for CU A-versus MCI/mild dementia A+ participants (AUC=.95; Table 2).

##### CU A- vs MCI A+

The MTD composite maintained high accuracy for CU A- vs MCI A+ participants (AUC=.93; Table 2).

### Biological AD Classification

#### A-T-versus A+T+

The unadjusted MTD composite alone significantly differentiated biomarker-defined groups when all clinical groups were combined (AUC=.76; Table 2). As expected, given the strong association of age and AD biomarkers, a demographics-only model showed good group discrimination (AUC=.82; Table 2). Adding the MTD composite significantly improved classification beyond demographics (AUC=.86; *p*=.009 for AUC comparison and *p*<.001 for MTD in the adjusted model; Table 2).

#### Preclinical AD

When the sample was limited to CU participants only (CU A-T- vs CU A+T+), the unadjusted MTD composite showed sensitivity to preclinical AD (AUC=.69; Table 2). A demographics-only model again showed good group discrimination (AUC=.84). The MTD composite significantly contributed to the odds of group prediction beyond demographic variables (*p*=.02; Table 2; Table S3).

### Comparison with Traditional In-Person Cognitive Measures

We restricted A-T- and A+T+ groups to individuals with both the MTD composite and select in-person cognitive measures to enable direct AUC comparisons. Clinical diagnosis was not used to define these groups to avoid circularity of diagnosis and person-administered measures. The MTD composite discriminated biomarker-defined groups more effectively than the MMSE or STMS-estimated MMSE (AUC 0.76 vs 0.70; *p*=.02; Figure 2 and Table S4). A similar result was observed for the STMS (Table S4). The MTD composite was not significantly different from the Mayo-PACC (*p*=.71; Figure 2) or a global cognitive composite (*p*=.67; Table S4) for discriminating A-T- and A+T+ groups.

**Figure 2.**
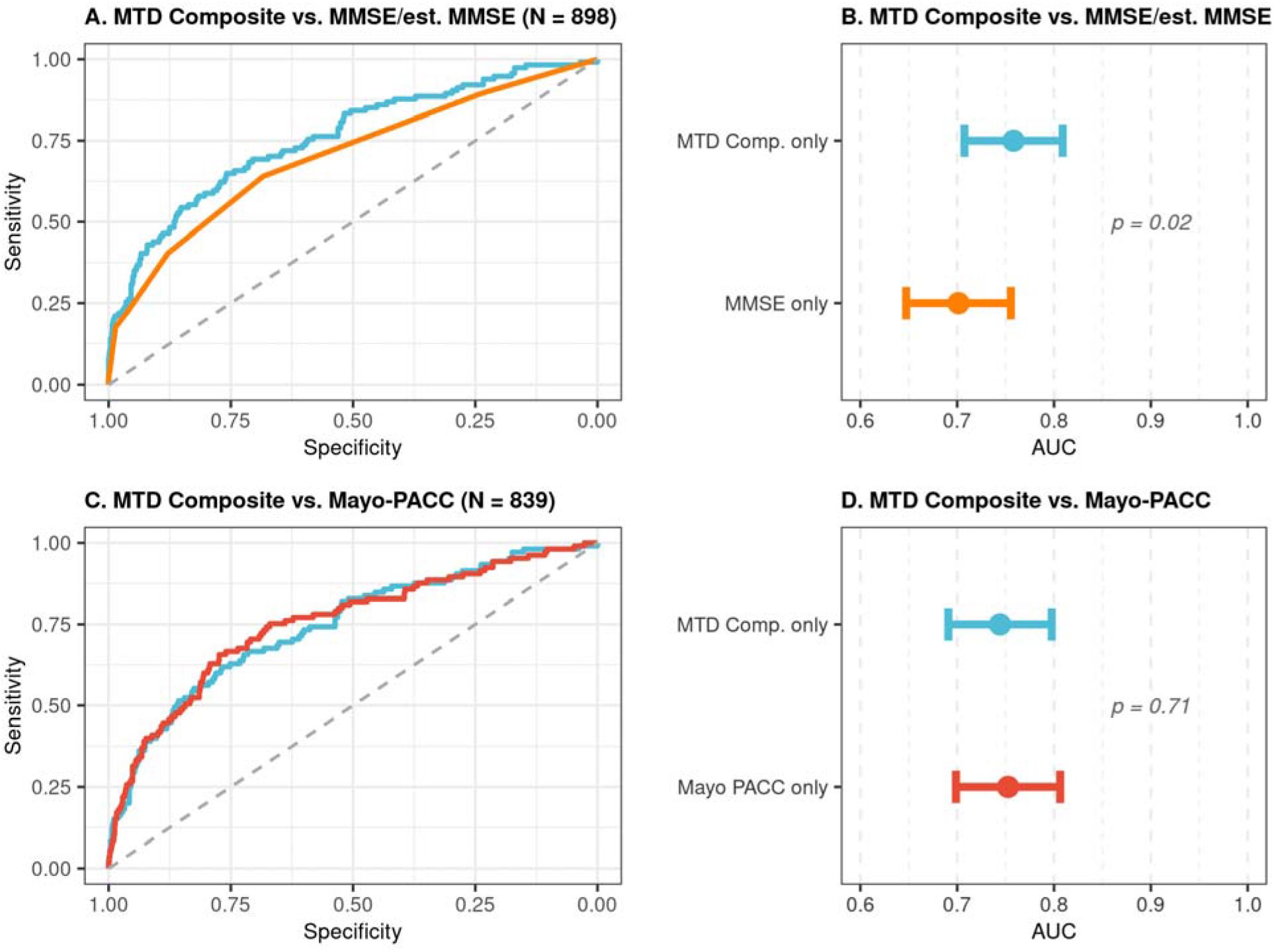
Model performance of MTD composite compared to in-person cognitive tests in predicting amyloid and tau PET positivity (A-T- versus A+T+). *Note.* Panels A and B show MTD Composite versus MMSE for differentiating A-T- (n=784) and A+T+ (n=114) independent of clinical diagnosis; MMSE scores represent the MMSE for participants from ADRC Florida (n=34) and the MMSE estimated from the Kokmen Short Test of Mental Status for all other participants. Panels C and D show MTD Composite versus Mayo-PACC for differentiating A-T- (n=734) and A+T+ (n=105) independent of clinical diagnosis. Analyses were completed independent of clinical diagnosis to avoid circularity as in-person cognitive measures are used to determine clinical diagnosis. MTD = Mayo Test Drive; MTD Comp. = Mayo Test Drive Composite; MMSE = Mini Mental Status Exam; Mayo-PACC = Mayo Clinic Preclinical Alzheimer’s Cognitive Composite; AUC = Area Under the Receiver Operating Characteristic Curve; PET = Positron Emission Tomography; A+ = Amyloid Positivity; A- = Amyloid Negativity; T+ = Tau Positivity; T- = Tau Negativity. Figure used with permission of Mayo Foundation for Medical Education and Research; all rights reserved.

### Additional Exploratory Analyses

Across additional amyloid-defined group predictions (A- versus A+, CU A- versus CU A+, CU A-T- versus CU A+T-), unadjusted MTD composite showed modest but significant discrimination, even among CU participants (AUCs=.62-.66; Table 2). Given the strong association of age and amyloid positivity, demographics-only models showed higher group discrimination performances (AUCs=.76-.77). The adjusted MTD composite was significantly associated with the group membership beyond demographic variables for the A- versus A+ (MTD *p*<.001) and CU A- versus CU A+ comparisons (MTD *p*=.04; Table 2). However, the MTD composite was not associated with group membership beyond demographic variables in individuals with early preclinical Alzheimer’s pathologic change (CU A-T- versus CU A+T-MTD *p*=.45; Table 2).

Select diagnostic accuracy analyses were also completed for subtest scores (Figure S2, Tables S5 and S6).

### Clinical Implications

#### Impact of demographically adjusted T-scores on diagnostic accuracy

The primary analyses above used the MTD composite raw score. We explored the impact of using a demographically-adjusted T-score (adjusted for age, sex and education) in a model with MTD alone as the predictor for select comparisons focusing on raw versus T-score differences for identification of treatment-relevant cognitive impairment and for early, preclinical cognitive change. Discriminatory ability remained strong when using the demographically-adjusted T-score for differentiating CU from MCI/mild dementia participants (AUC = 0.86; 95% CI = .84–.88). However, this AUC was significantly lower than that of the raw score model AUC of .89 (*p* < .001). Similarly, for differentiating CU A–T– from CU A+T+ groups, the adjusted T-score yielded a lower AUC (0.58; 95% CI = .51–.65) compared to the raw score’s AUC of 0.69 (*p* < .001 for AUC comparison).

#### Sensitivity, specificity, two-stage classification, and interpretation guidelines

Sensitivity, specificity, and corresponding 2x2 data (e.g., true positive, false positive, true negative, false negative categories) across a range of data-driven and clinically guided cutoffs and score types (raw, T-score) across several group comparisons are provided to inform clinical and research applications (Figure 3, Table 3, Tables S7-S10). Select subtest-level data are also included (Figure S2, Tables S11-S16). A selection of these values informative for deriving MTD composite cutoff scores is provided in Table 3.

**Figure 3.**
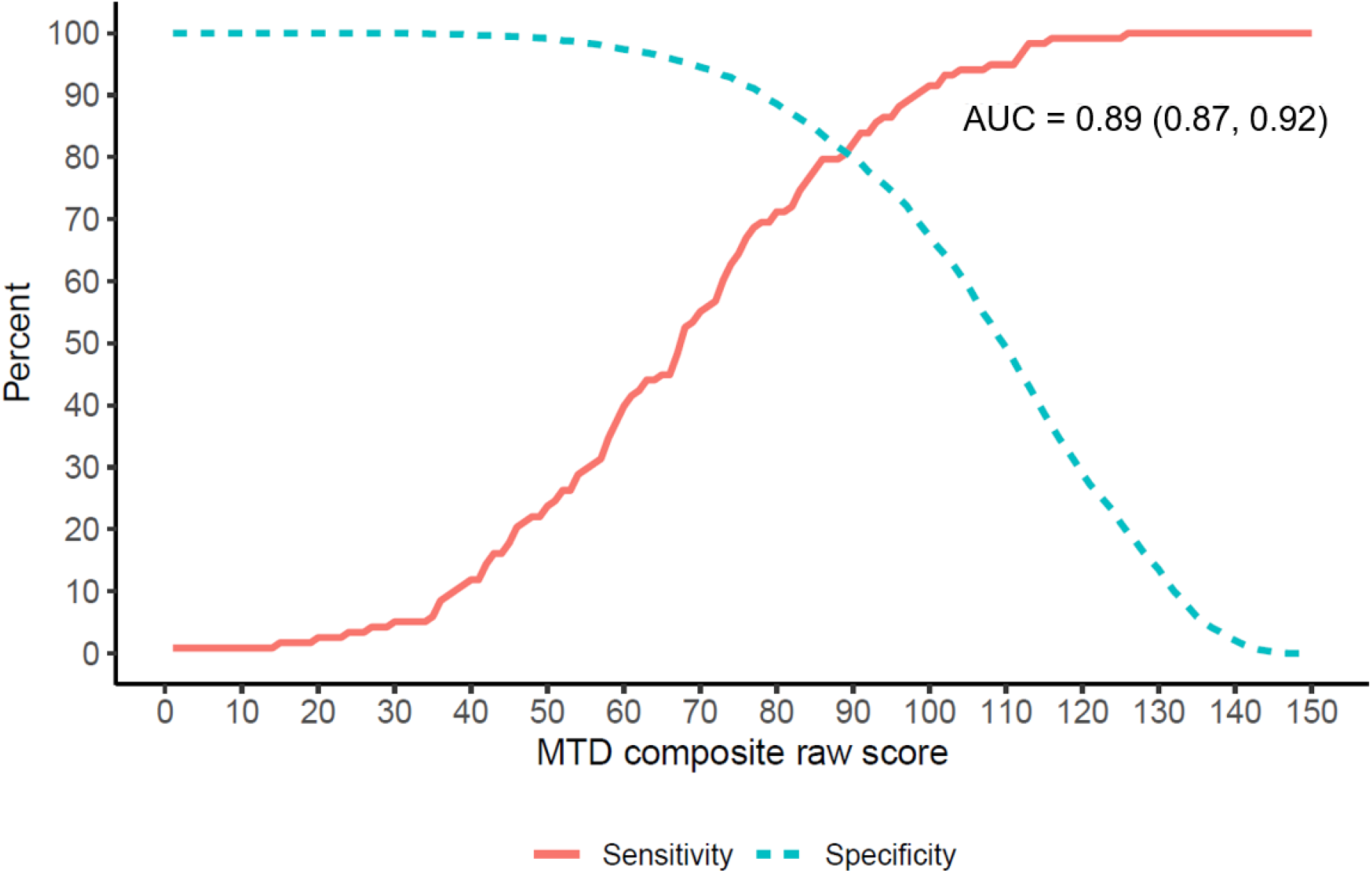
Sensitivity and specificity of MTD composite for differentiating cognitively unimpaired versus cognitively impaired (MCI or mild dementia). *Note.* AUC represents the area under the receiver operating characteristic curve with the 95% confidence interval in parenthesis; MCI = mild cognitive impairment; MTD = Mayo Test Drive. Figure used with permission of Mayo Foundation for Medical Education and Research; all rights reserved.

**Table 3.**
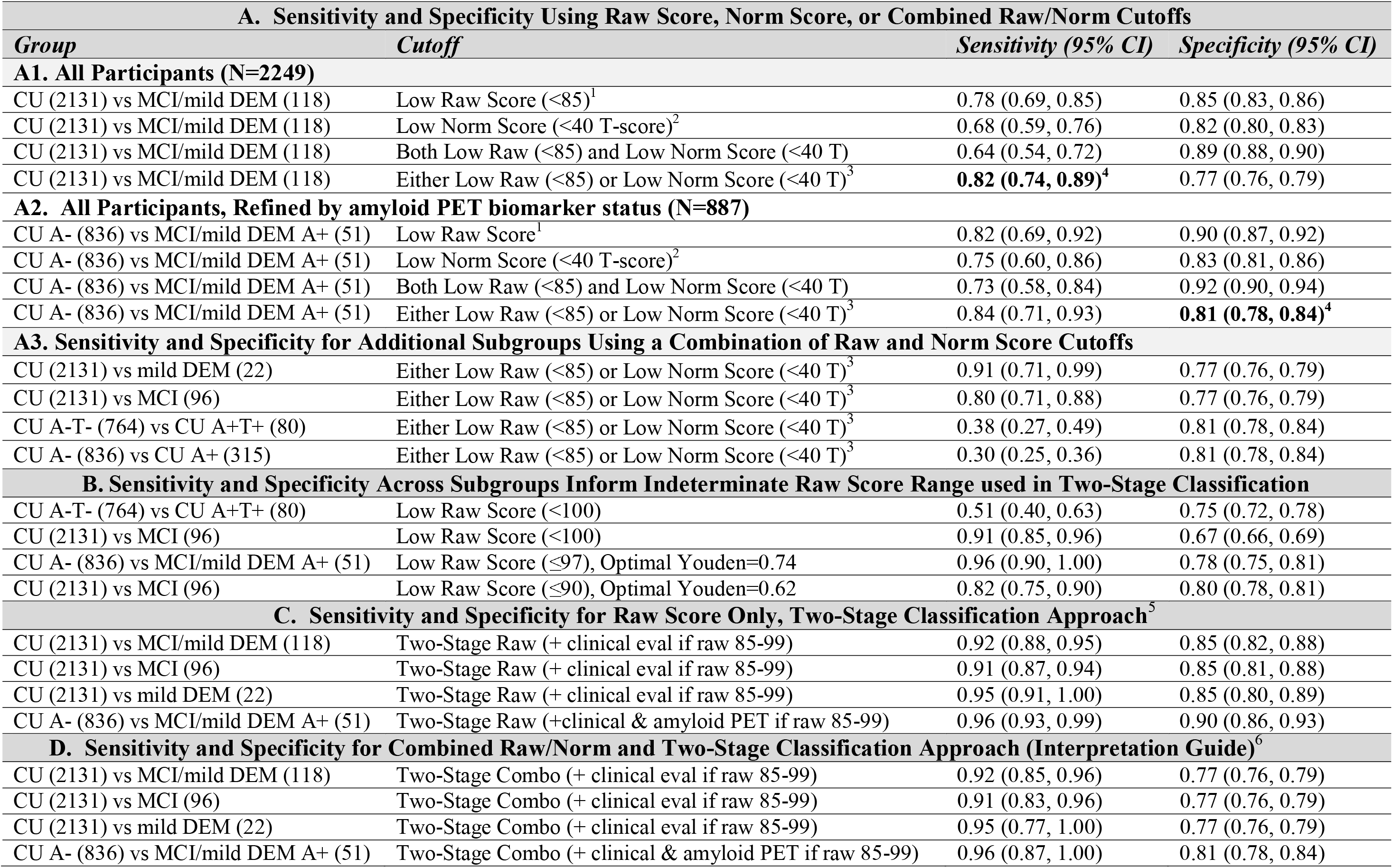
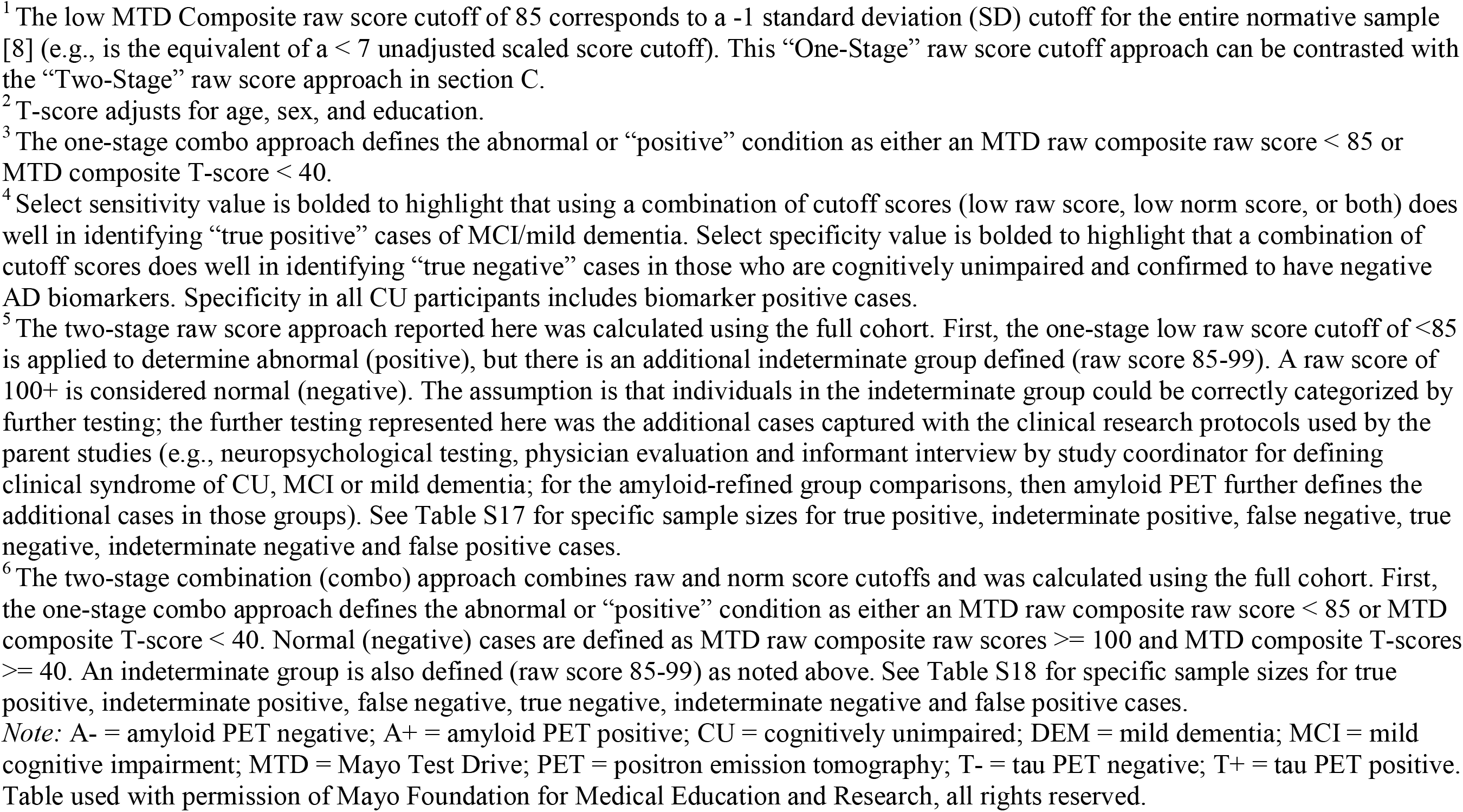
Select sensitivity and specificity values for the MTD composite across various cutoffs and subgroups.

| **A. Sensitivity and Specificity Using Raw Score, Norm Score, or Combined Raw/Norm Cutoffs** |  |  |  |
| --- | --- | --- | --- |
| **Group** | **Cutoff** | **Sensitivity (95% CI)** | **Specificity (95% CI)** |
| **A1. All Participants (N=2249)** |  |  |  |
| CU (2131) vs MCI/mild DEM (118) | Low Raw Score (<85)<sup>1</sup> | 0.78 (0.69, 0.85) | 0.85 (0.83, 0.86) |
| CU (2131) vs MCI/mild DEM (118) | Low Norm Score (<40 T-score)<sup>2</sup> | 0.68 (0.59, 0.76) | 0.82 (0.80, 0.83) |
| CU (2131) vs MCI/mild DEM (118) | Both Low Raw (<85) and Low Norm Score (<40 T) | 0.64 (0.54, 0.72) | 0.89 (0.88, 0.90) |
| CU (2131) vs MCI/mild DEM (118) | Either Low Raw (<85) or Low Norm Score (<40 T)<sup>3</sup> | **0.82 (0.74, 0.89)<sup>4</sup>** | 0.77 (0.76, 0.79) |
| **A2. All Participants, Refined by amyloid PET biomarker status (N=887)** |  |  |  |
| CU A- (836) vs MCI/mild DEM A+ (51) | Low Raw Score<sup>1</sup> | 0.82 (0.69, 0.92) | 0.90 (0.87, 0.92) |
| CU A- (836) vs MCI/mild DEM A+ (51) | Low Norm Score (<40 T-score)<sup>2</sup> | 0.75 (0.60, 0.86) | 0.83 (0.81, 0.86) |
| CU A- (836) vs MCI/mild DEM A+ (51) | Both Low Raw (<85) and Low Norm Score (<40 T) | 0.73 (0.58, 0.84) | 0.92 (0.90, 0.94) |
| CU A- (836) vs MCI/mild DEM A+ (51) | Either Low Raw (<85) or Low Norm Score (<40 T)<sup>3</sup> | 0.84 (0.71, 0.93) | **0.81 (0.78, 0.84)<sup>4</sup>** |
| **A3. Sensitivity and Specificity for Additional Subgroups Using a Combination of Raw and Norm Score Cutoffs** |  |  |  |
| CU (2131) vs mild DEM (22) | Either Low Raw (<85) or Low Norm Score (<40 T)<sup>3</sup> | 0.91 (0.71, 0.99) | 0.77 (0.76, 0.79) |
| CU (2131) vs MCI (96) | Either Low Raw (<85) or Low Norm Score (<40 T)<sup>3</sup> | 0.80 (0.71, 0.88) | 0.77 (0.76, 0.79) |
| CU A-T- (764) vs CU A+T+ (80) | Either Low Raw (<85) or Low Norm Score (<40 T)<sup>3</sup> | 0.38 (0.27, 0.49) | 0.81 (0.78, 0.84) |
| CU A- (836) vs CU A+ (315) | Either Low Raw (<85) or Low Norm Score (<40 T)<sup>3</sup> | 0.30 (0.25, 0.36) | 0.81 (0.78, 0.84) |
| **B. Sensitivity and Specificity Across Subgroups Inform Indeterminate Raw Score Range used in Two-Stage Classification** |  |  |  |
| CU A-T- (764) vs CU A+T+ (80) | Low Raw Score (<100) | 0.51 (0.40, 0.63) | 0.75 (0.72, 0.78) |
| CU (2131) vs MCI (96) | Low Raw Score (<100) | 0.91 (0.85, 0.96) | 0.67 (0.66, 0.69) |
| CU A- (836) vs MCI/mild DEM A+ (51) | Low Raw Score (≤97), Optimal Youden=0.74 | 0.96 (0.90, 1.00) | 0.78 (0.75, 0.81) |
| CU (2131) vs MCI (96) | Low Raw Score (≤90), Optimal Youden=0.62 | 0.82 (0.75, 0.90) | 0.80 (0.78, 0.81) |
| **C. Sensitivity and Specificity for Raw Score Only, Two-Stage Classification Approach<sup>5</sup>** |  |  |  |
| CU (2131) vs MCI/mild DEM (118) | Two-Stage Raw (+ clinical eval if raw 85-99) | 0.92 (0.88, 0.95) | 0.85 (0.82, 0.88) |
| CU (2131) vs MCI (96) | Two-Stage Raw (+ clinical eval if raw 85-99) | 0.91 (0.87, 0.94) | 0.85 (0.81, 0.88) |
| CU (2131) vs mild DEM (22) | Two-Stage Raw (+ clinical eval if raw 85-99) | 0.95 (0.91, 1.00) | 0.85 (0.80, 0.89) |
| CU A- (836) vs MCI/mild DEM A+ (51) | Two-Stage Raw (+clinical & amyloid PET if raw 85-99) | 0.96 (0.93, 0.99) | 0.90 (0.86, 0.93) |
| **D. Sensitivity and Specificity for Combined Raw/Norm and Two-Stage Classification Approach (Interpretation Guide)<sup>6</sup>** |  |  |  |
| CU (2131) vs MCI/mild DEM (118) | Two-Stage Combo (+ clinical eval if raw 85-99) | 0.92 (0.85, 0.96) | 0.77 (0.76, 0.79) |
| CU (2131) vs MCI (96) | Two-Stage Combo (+ clinical eval if raw 85-99) | 0.91 (0.83, 0.96) | 0.77 (0.76, 0.79) |
| CU (2131) vs mild DEM (22) | Two-Stage Combo (+ clinical eval if raw 85-99) | 0.95 (0.77, 1.00) | 0.77 (0.76, 0.79) |
| CU A- (836) vs MCI/mild DEM A+ (51) | Two-Stage Combo (+ clinical & amyloid PET if raw 85-99) | 0.96 (0.87, 1.00) | 0.81 (0.78, 0.84) |

<sup>1</sup> The low MTD Composite raw score cutoff of 85 corresponds to a -1 standard deviation (SD) cutoff for the entire normative sample [8] (e.g., is the equivalent of a < 7 unadjusted scaled score cutoff). This “One-Stage” raw score cutoff approach can be contrasted with the “Two-Stage” raw score approach in section C.
<sup>2</sup> T-score adjusts for age, sex, and education.
<sup>3</sup> The one-stage combo approach defines the abnormal or “positive” condition as either an MTD raw composite raw score < 85 or MTD composite T-score < 40.
<sup>4</sup> Select sensitivity value is bolded to highlight that using a combination of cutoff scores (low raw score, low norm score, or both) does well in identifying “true positive” cases of MCI/mild dementia. Select specificity value is bolded to highlight that a combination of cutoff scores does well in identifying “true negative” cases in those who are cognitively unimpaired and confirmed to have negative AD biomarkers. Specificity in all CU participants includes biomarker positive cases.
<sup>5</sup> The two-stage raw score approach reported here was calculated using the full cohort. First, the one-stage low raw score cutoff of <85 is applied to determine abnormal (positive), but there is an additional indeterminate group defined (raw score 85-99). A raw score of 100+ is considered normal (negative). The assumption is that individuals in the indeterminate group could be correctly categorized by further testing; the further testing represented here was the additional cases captured with the clinical research protocols used by the parent studies (e.g., neuropsychological testing, physician evaluation and informant interview by study coordinator for defining clinical syndrome of CU, MCI or mild dementia; for the amyloid-refined group comparisons, then amyloid PET further defines the additional cases in those groups). See Table S17 for specific sample sizes for true positive, indeterminate positive, false negative, true negative, indeterminate negative and false positive cases.
<sup>6</sup> The two-stage combination (combo) approach combines raw and norm score cutoffs and was calculated using the full cohort. First, the one-stage combo approach defines the abnormal or “positive” condition as either an MTD raw composite raw score < 85 or MTD composite T-score < 40. Normal (negative) cases are defined as MTD raw composite raw scores $\geq 100$ and MTD composite T-scores $\geq 40$ . An indeterminate group is also defined (raw score 85-99) as noted above. See Table S18 for specific sample sizes for true positive, indeterminate positive, false negative, true negative, indeterminate negative and false positive cases.
*Note:* A- = amyloid PET negative; A+ = amyloid PET positive; CU = cognitively unimpaired; DEM = mild dementia; MCI = mild cognitive impairment; MTD = Mayo Test Drive; PET = positron emission tomography; T- = tau PET negative; T+ = tau PET positive. Table used with permission of Mayo Foundation for Medical Education and Research, all rights reserved.

##### Two-stage raw score approach

A two-stage framework was developed for the MTD composite raw score to reduce the risk of potential false negative errors and evaluate the performance of a raw score only approach. To support this approach, a three-range classification scheme informed by observed sensitivity and specificity patterns was defined. Scores <85 were classified as abnormal (positive), scores ≥100 were classified as normal (negative), and scores between 85 and 99 were considered indeterminate and would require additional evaluation in the second stage of the framework. For the current analyses, indeterminate cases were classified as positive or negative using the reference standard for the specific comparison (research diagnosis, PET imaging biomarkers, or both). Relative to a single-threshold approach, the two-stage framework improved sensitivity while preserving clinically useful specificity (Table 3; Table S17).

##### Combined raw and norm score approach

A novel approach that combines both raw score and normative score cutoffs (“one-stage combo” approach) helped balance sensitivity and specificity relative to use of either a raw score or norm score cutoff in isolation (Table 3, Figure 4). Specifically, this approach classifies individuals based on concordant consideration of both raw score and normative score thresholds rather than either score type in isolation.

**Figure 4.**
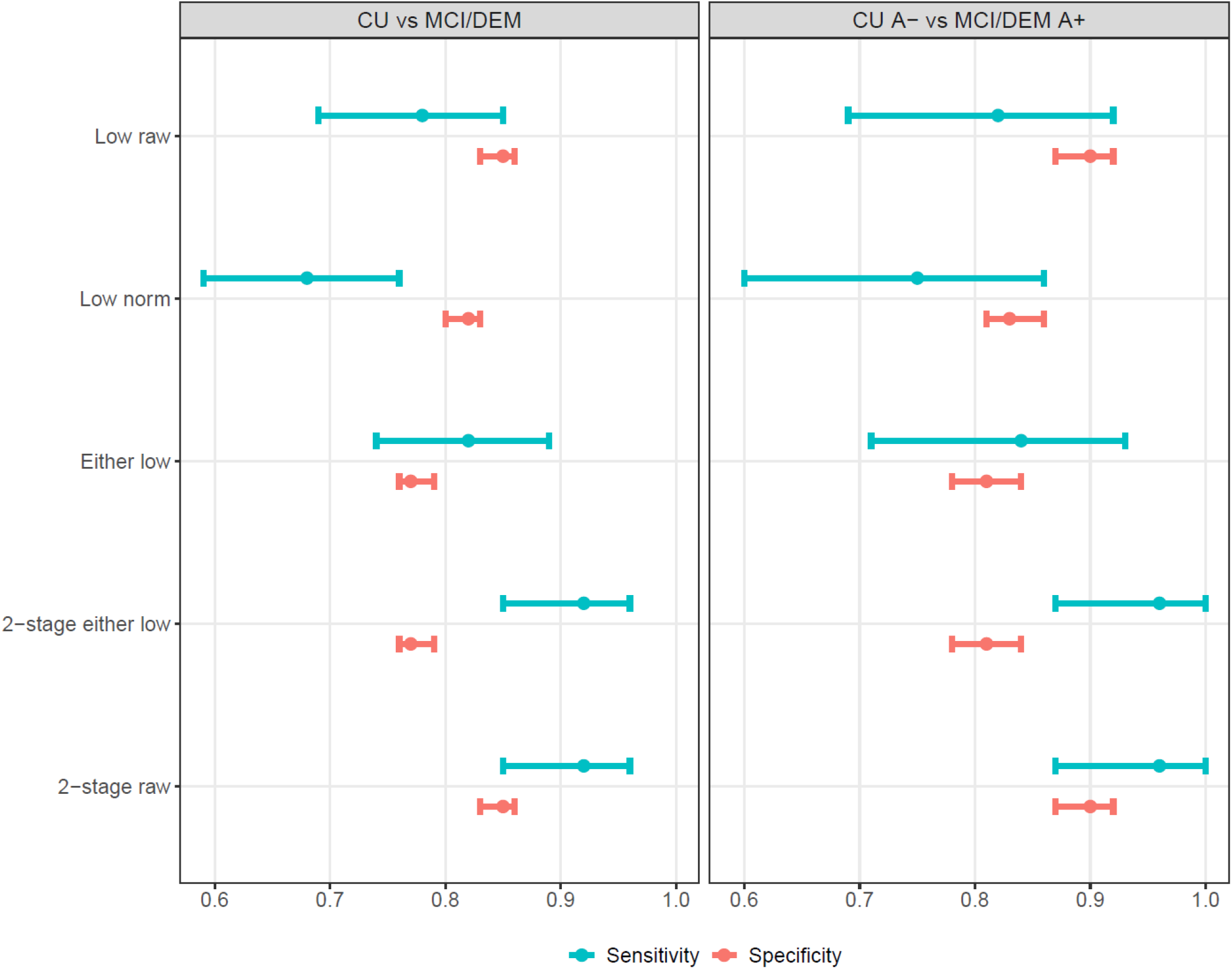
Sensitivity and specificity (with 95% confidence intervals) of various MTD composite cutoff approaches for CU vs MCI/DEM and CU A- vs MCI/DEM A+ comparisons. *Note.* Low raw = MTD composite raw score < 85 (also represents one-stage raw score approach); Low norm = MTD composite age-, sex-, and education-adjusted T-score < 40; Either low = either low raw or low norm (also represents one-stage either low approach also known as one-stage combo approach); 2-stage either low = MTD raw composite raw scores >= 100 AND MTD composite T-scores >= 40 were categorized as belonging to a “normal” or control population, MTD raw composite raw scores < 85 were categorized as having the abnormal or “positive” condition, MTD composite T-scores < 40 were categorized as having the abnormal or “positive” condition, MTD raw composite raw scores between 85-99 (AND not classified as positive by the normative score cutoff), inclusive, were not categorized; 2-stage raw = MTD raw composite raw scores >= 100 were categorized as belonging to a “normal” or control population, MTD raw composite raw scores < 85 were categorized as having the abnormal or “positive” condition, MTD raw composite scores between 85-99, inclusive, were not categorized. Figure used with permission of Mayo Foundation for Medical Education and Research; all rights reserved.

##### Two-stage combined raw and norm score approach

Adding the indeterminate range from the two-stage raw score approach to the combined approach (“two-stage combo” approach) further improved sensitivity (Table 3; Table S18). This approach combines raw score and normative score cutoffs with selective application of a two-stage classification framework for individuals whose raw scores fall within the indeterminate range, thereby minimizing false negative classifications. The resulting framework was incorporated into an interpretation guide (Table 4).

**Table 4.** Interpretation guide for combined raw and normative score approach in assessing Mayo Test Drive Composite performance.

| Raw Score | Normative Score | Raw/Norm Agreement | Result |
| --- | --- | --- | --- |
| 100+ (above cutoff) | 40+ (above cutoff) | Concordant | Within expected range |
| 85+ and <100 (above cutoff) | 40+ (above cutoff) | Concordant | Within expected range but consider follow-up if concern present* |
| 85+ (above cutoff) | <40 (below cutoff) | Discordant | Below expected range – low norm score |
| <85 (below cutoff) | 40+ (above cutoff) | Discordant | Below expected range – low raw score |
| <85 (below cutoff) | <40 (below cutoff) | Concordant | Below expected range – both low norm and low raw scores |

\*Two-stage classification is embedded into the combined raw and normative score approach. If results are normal but the raw score is 85-99 and there is clinical context that raises concern, consider additional steps to avoid a potential false negative (e.g., neuropsychologist reviews Mayo Test Drive subtest results in context of available clinical history and session characteristics, referral for full neuropsychological evaluation, behavioral neurology evaluation, plasma biomarker testing, longitudinal monitoring, etc., as appropriate for the setting; also see Table 3).
*Note.* The raw score cutoff of <85 is equivalent to a cutoff of -1 standard deviation (SD) based on the unadjusted scaled score from the normative sample of < 7 scaled score (mean 10, SD 3; citation). The < 40 demographically adjusted T-score also represents a < -1 SD cutoff, based on the age-, sex-, and education-adjusted T-score (mean 50, SD 10). Table used with permission of Mayo Foundation for Medical Education and Research; all rights reserved.

## Discussion

A remote, self-administered digital cognitive screening battery demonstrated strong diagnostic accuracy for detecting MCI and mild dementia, providing evidence of criterion validity when evaluated against amyloid- and tau-PET-defined biomarker groups in this large, well-characterized, population-based cohort. The MTD composite also showed comparable performance to established in-person cognitive composites, supporting its potential role as a scalable method of identifying individuals who may benefit from further cognitive and biomarker evaluation.

### Clinical sensitivity and treatment relevance

Consistent with our prespecified analytic plan, MTD exceeded the *a priori* threshold of .85 for identifying MCI and mild dementia (AUC=.89; 95% CI=.87-.92). Performance improved further when diagnostic groups were refined by amyloid status (CU A- vs MCI/DEM A+ AUC=.95), highlighting the complementary value of cognitive characterization and biomarker information. Diagnostic accuracy remained strong when excluding individuals with mild dementia from the clinical group comparison (CU vs MCI AUC=.88), indicating that performance was not driven solely by inclusion of participants with dementia. Prior studies have shown that digital cognitive assessments differentiate individuals with and without cognitive impairment, though most were conducted on-site or hybrid approaches [34–36], and remote registry-based samples typically rely on self-reported cognitive status [37]. To our knowledge, no prior study has combined all of these elements, including fully remote self-administered, multi-device-compatible cognitive assessment with detailed clinical characterization, amyloid- and tau-defined subgroups, and direct comparisons with established cognitive composites in a large, primarily population-based cohort [34, 38–41]. Further, although many digital cognitive assessment studies have reported AUCs demonstrating discrimination between cognitively unimpaired and cognitively impaired individuals, fewer have provided clinically actionable cutoffs or reported sensitivity and specificity estimates necessary for clinical implementation [34, 42–44].

The current findings support the use of remote digital cognitive tools for early detection and clinical characterization, particularly in settings where access to comprehensive neuropsychological evaluation is limited. For example, MTD has promise for use as an early detection tool in primary care clinics; the two-stage sensitivity and specificity values meet the criteria specified for a digital cognitive assessment for detecting cognitive impairment [45], and the availability of normative scores to consider alongside raw scores allow for a more nuanced understanding of test performance relative to use of a raw score in isolation. Evaluation of sensitivity and specificity also needs to consider the population represented in the study sample; population-based samples with broad inclusion/exclusion criteria, such as that a majority of participants represented in this study, may yield lower sensitivity and specificity than samples enrolled from specialty clinics due to referral bias and disease severity effects in those specialty clinics, contributing to spectrum effects (i.e., variation in test performance across populations that differ in disease severity representation) [46–48].

### Criterion validity

The MTD composite demonstrated moderate discrimination of PET-defined biomarker groups (A-T- vs A+T+) and meaningfully improved classification beyond demographic variables. This finding supports MTD criterion validity when amyloid and tau PET biomarkers are used as an independent reference standard and aligns with prior work showing that cognition captures disease-relevant variance not fully explained by demographics such as age, sex or education alone [29, 49, 50]. Importantly, the MTD composite performed similarly to more time-intensive, in-person cognitive composites in differentiating A+T+ individuals from those without AD pathology and outperformed a brief mental status screening measure.

### Preclinical AD and early AD pathologic change

Sensitivity of MTD to preclinical AD was modest, particularly when analyses were restricted to CU individuals. This attenuation in sensitivity in the preclinical phase is expected, as cognitive differences are variably reported across CU A+ versus CU A- groups [51, 52], and earlier work by our group demonstrated that neurodegenerative changes drive early cognitive symptoms [53]. Other investigations have shown that neuropsychological tests do not enhance prediction of amyloid positivity beyond demographics and genetic information in CU individuals [54], so it is notable that MTD demonstrated any sensitivity, albeit modest. Ultimately, these findings underscore that remote cognitive screening should not be interpreted as a stand-alone biomarker substitute in preclinical AD, but rather as a complementary tool that may help contextualize risk and inform downstream evaluation.

### Sample characteristics impact diagnostic accuracy

The clear pattern of strong discriminative performance of MTD for clinically defined groups, contrasted with the predominant role of age in predicting biomarker-defined groups, particularly among CU individuals, suggests that cognitive measures are most informative when the comparison groups differ in the clinical expression of cognitive impairment. Cognitive measures perform best when the groups under comparison are cognitively distinct. For example, in a predominantly CU cohort such as the present sample (95% CU), A+ and A- individuals are largely cognitively normal and cognitive differences associated with biomarker status are often subtle, limiting the discriminative performance of cognitive measures. In contrast, studies that evaluate A+ and A- individuals across a broader spectrum of clinical severity may include A+ participants with greater cognitive impairment and A- participants with relatively preserved cognition, resulting in larger cognitive differences between groups and, consequently, stronger discriminative performance [55, 56]. Thus, the observed performance of a cognitive measure depends largely on the extent to which the groups being compared differ cognitively. Taken together, these findings emphasize the importance of considering sample composition when interpreting diagnostic accuracy estimates and suggest that head-to-head comparisons remain the most informative approach for comparing cognitive measures. The direct comparison of digital and traditional person-administered measures is therefore a strength of the current study.

### Raw versus normative scores and clinical interpretation

The MTD composite raw score outperformed the demographically-adjusted normative score in both clinically and biomarker-defined group comparisons. This pattern suggests that demographic adjustment may remove variance related to disease risk that is relevant for screening purposes [57]. Sensitivity and specificity were examined across multiple data-driven and clinically-guided cutoffs, score types, and group comparisons to inform clinical and research applications. This multi-subgroup view is advantageous because preferred MTD cutoff scores may vary by context of use. Interpretation of specificity varies significantly by CU subgroup given the inclusion of individuals with positive AD biomarkers or other unmeasured early neurodegenerative changes in CU participants. Common clinical standards informed the initial suggested MTD cutoffs (Table 4), along with sensitivity and specificity estimates across subgroups. We provide a unique interpretation guide that combines raw and normative score cutoffs to provide a better balance of sensitivity and specificity than use of either score type in isolation. We additionally incorporate elements of a two-stage cutoff approach for scores in a range that may benefit from additional clinical consideration or work-up in settings where high sensitivity is needed. Screening measures frequently suffer from a stark imbalance in sensitivity and specificity when trying to rely on overly simplistic single cutoffs that do not reflect the imperfect nature of cognitive tests and the nuanced realities of clinical decision-making. Accordingly, our interpretation guide was designed to provide a more flexible framework for balancing these competing considerations across different clinical and research contexts. This interpretation guide informs initial clinical interpretation of MTD.

These guidelines will require independent validation, are likely to evolve over time, and may need adjustment for specific settings, contexts of use, or individual characteristics. Local verification of cutoffs is similarly recommended for plasma biomarkers [58]. We also characterize a two-stage MTD composite raw score approach, as is commonly done in AD plasma biomarker studies [59–61]. Raw scores offer consistency and transparency because normative data are likely to be updated over time. Raw scores may therefore facilitate longitudinal consistency. In addition, practical considerations can complicate use of normative scores for screening measures (e.g., demographic information must be available for real-time use), and demographic adjustment may reduce sensitivity for detecting early cognitive change in older adults [62]. Caution is needed when applying these guidelines and cutoffs to individuals not well represented in the current sample. Future work will examine whether modifications are needed for individuals not well represented in the current analytic or normative samples based on race, ethnicity, education, and linguistic background.

### Strengths, Limitations, and Future Directions

Key strengths of this study include the large, well-characterized, predominantly population-based sample, availability of amyloid and tau PET imaging in over half of participants, independence of MTD to diagnosis, avoidance of circularity in comparisons to traditional neuropsychological measures through use of AD-biomarker defined groups, and presentation of multiple sample subgroups that help illustrate the important role of sample characteristics in diagnostic accuracy results. Limitations include the predominantly non-Hispanic White, highly educated sample, ancillary and voluntary nature of the digital cognitive assessment component of the study that may contribute to sample bias, and cross-sectional design. Work is underway to expand MTD use and enable validation in more heterogeneous populations. This study was not designed or sufficiently powered to determine how well MTD differentiates earlier from later stages of symptomatic AD, and individuals with moderate dementia were excluded. Future work is needed to address prognostic accuracy, refinement of interpretation guidelines for specific use cases, and evaluation within real-world clinical workflows.

## Conclusions

These findings support MTD’s role as a scalable digital cognitive screening tool that may complement biomarker testing, inform referral for additional evaluation, and support clinical trial enrichment in the evolving landscape of AD care.

## Supporting information

Supplemental Online Material for Diagnostic accuracy of remote self administered digital cognitive assessment with STARDDem checklist

## Data Availability

All data produced in the present study are available upon reasonable request to the authors.

## Funding and Acknowledgments

Research reported in this publication was supported by the National Institute on Aging of the National Institutes of Health under Award Numbers R01AG081955, R21 AG073967, P30 AG062677, U01 AG006786, U19 AG102336, and R01 AG034676 (the Rochester Epidemiology Project). This work was also supported by the Kevin Merszei Career Development Award in Neurodegenerative Diseases Research IHO Janet Vittone, MD, the GHR Foundation, Gates Ventures and the Mayo Foundation for Medical Education and Research. The content is solely the responsibility of the authors and does not necessarily represent the official views of the National Institutes of Health or other sponsors. A Mayo Clinic invention disclosure has been submitted for the Stricker Learning Span and the Mayo Test Drive platform (NHS, JLS). We have no other conflicts of interest to disclose related to this work. The authors wish to thank the participants and staff at the Mayo Clinic Study of Aging and Mayo Alzheimer’s Disease Research Center.

