## Supplemental Online Material for Diagnostic accuracy of remote self administered digital cognitive assessment with STARDDem checklist for "Diagnostic accuracy of remote self-administered digital cognitive assessment by clinical diagnosis and Alzheimer’s disease PET imaging biomarkers"

**This is supplemental online materials for a preprint. This manuscript has not yet been peer reviewed by a journal.**

**Copyright 2026 Mayo Foundation for Medical Education and Research. All rights reserved.**

**Supplemental Methods**

*Prospective Study Design*

This study was prospectively designed as a diagnostic accuracy investigation of remote digital cognitive assessment. Preliminary analyses were conducted on a small subset of participants (13% of the current sample; N=298, including 20 participants with mild cognitive impairment (MCI)) using an early z-score composite presented in a conference abstract that differed from the Mayo Test Drive (MTD) raw score composite examined in the present study [1]. Initial data collection and analyses were focused primarily on establishing the criterion validity of the Stricker Learning Span (SLS, NIA R21 AG073967) and also included the Symbols test (SYM).

The current MTD composite was theoretically developed based on clinical considerations and review of preclinical Alzheimer's disease cognitive composite studies and implemented in May 2022. Development also incorporated observed frequencies of SYM matching errors to determine weighting applied to the Symbols Accuracy Weighted Score. No modifications were made to the MTD raw score composite after its implementation.

The current diagnostic accuracy aims were subsequently incorporated into an NIH R01 application first submitted in June 2022 and resubmitted in March 2023. Thus, primary analyses reported herein were conducted within a prospectively designed study framework in which the index test and major study aims were specified before completion of data collection. Preliminary data from a subset of participants were used to inform sample size targets and power calculations using the Power Analysis and Sample Size (PASS) software ([2, 3]).

*A priori*, we hypothesized that the MTD composite raw score would demonstrate excellent discrimination between CU and MCI participants (AUC > .85). Sample size planning targeted approximately 120 MCI participants. If this recruitment target was not achieved, a prespecified alternative analysis combined MCI and mild dementia participants into a broader cognitively impaired group for evaluation of diagnostic accuracy. CU sample size planning was based on the overall sample size of the MCSA and MTD ancillary study accrual rates observed at the time of the R01 submission; we projected 1780 CU participants would be available by the time of analysis. Sample sizes of 1780 CU participants and 120 MCI/mild dementia participants had 83% projected power to reject the null hypothesis AUC of 0.85 (a confidence interval with a lower limit that does not include 0.85). For the amyloid-refined clinical diagnosis groups, sample sizes of 662 CU A- and 40 MCI/mild dementia A+ participants had 81% projected power to reject the null hypothesis of AUC of 0.85.

We implemented the prespecified alternative analysis plan of using both MCI and mild dementia participants in our cognitive impairment group to stay on track with planned analysis timelines for funding and because this also aligns more closely with contemporary monoclonal antibody treatment decision-making pathways, which include both MCI and mild dementia stages [4]. Participants with MCI and mild dementia were therefore combined to create a cognitively impaired group (n=118) for the primary diagnostic accuracy analysis. The majority of participants in this group (81%) were MCI participants, and a smaller proportion (19%) were mild dementia participants. We also provide results using the CU vs MCI comparison.

The primary hypothesis for the *a priori* A-T- vs A+T+ participant comparison was that MTD would show similar ability to differentiate A-T- from A+T+ participants compared to traditional in-person measures. This analysis was based on all participants (instead of CU only) to avoid introducing circularity for the in-person measures that are considered for diagnosis. We additionally pre-specified an AUC to aid sample size projections, but considered the predominance of CU participants expected in our sample based on the primary parent study (MCSA) known sample composition when selecting the AUC target for hypothesis testing of 0.60, anchored by our previously published work using data collected previously in the same parent study cohort (MCSA) that reported AUC for measures from the Cogstate Brief Battery, the Rey's Auditory Verbal Learning Test and the Kokmen Short Test of Mental Status for CU A-T- (n=146) vs CU A+T+ (n=33) participants (all testing done in person in that prior study; Learning/Working Memory Composite age-corrected z-score AUC= 0.60, 95% CI = 0.49 – 0.71; One Card Learning test accuracy raw score AUC = 0.62, 95% CI = .51, .73; AVLT delay raw score AUC = 0.67, 95% CI = .56 - .78, Kokmen STMS AUC = 0.57, 95% CI = 0.46 – 0.68) [5]. For CU A-T- vs CU A+T+ comparisons, we pre-specified an AUC of 0.55 for sample size projections and hypothesis testing for sensitivity to preclinical AD. Sample sizes of 481 A-T- and 136 A+T+ had 93% projected power to reject the null hypothesis AUC of 0.60 (a confidence interval with a lower limit that does not include 0.60). For the CU only refinement, sample sizes of 442 CU A-T- and 100 CU A+T+ participants had 90% projected power to reject the null hypothesis of AUC of 0.55. Of note, at the time the R01 application was written, we defined amyloid and tau positivity as SUVR ≥ 1.48 (centiloid 22) [6] and ≥ 1.25 [7] for power projections. For the current analyses, we applied updated cutoffs in use at Mayo Clinic in our group, as reported in the manuscript. The older cutoffs yielded larger sample sizes of A+ and T+ individuals, thus the new imaging cutoffs result in fewer individuals labeled as A+T+ and more individuals labeled as A-T- relative to old cutoffs. Thus, although this leads to a slightly lower number of A+T+ individuals than projected in power analyses with the new cutoffs (which would have been met with the old cutoffs), there is increased confidence in the amyloid and tau positive status for individuals meeting current updated cutoffs thus those were selected for implementation in analyses.

**Supplemental Results**

No adverse events were reported during this ancillary, minimal risk study.

**Figure S1.** STARDdem participant flow and diagnostic classification performance of the Mayo Test Drive (MTD) composite using the either low raw score (MTD composite < 85) or low normative score (MTD composite T-score < 40) cutoff approach for discrimination of cognitively unimpaired (CU) participants from participants with mild cognitive impairment or mild dementia (MCI/DEM). Percentages represent sensitivity and specificity components calculated within the corresponding reference-standard group. Raw counts and denominators are shown beneath each percentage.

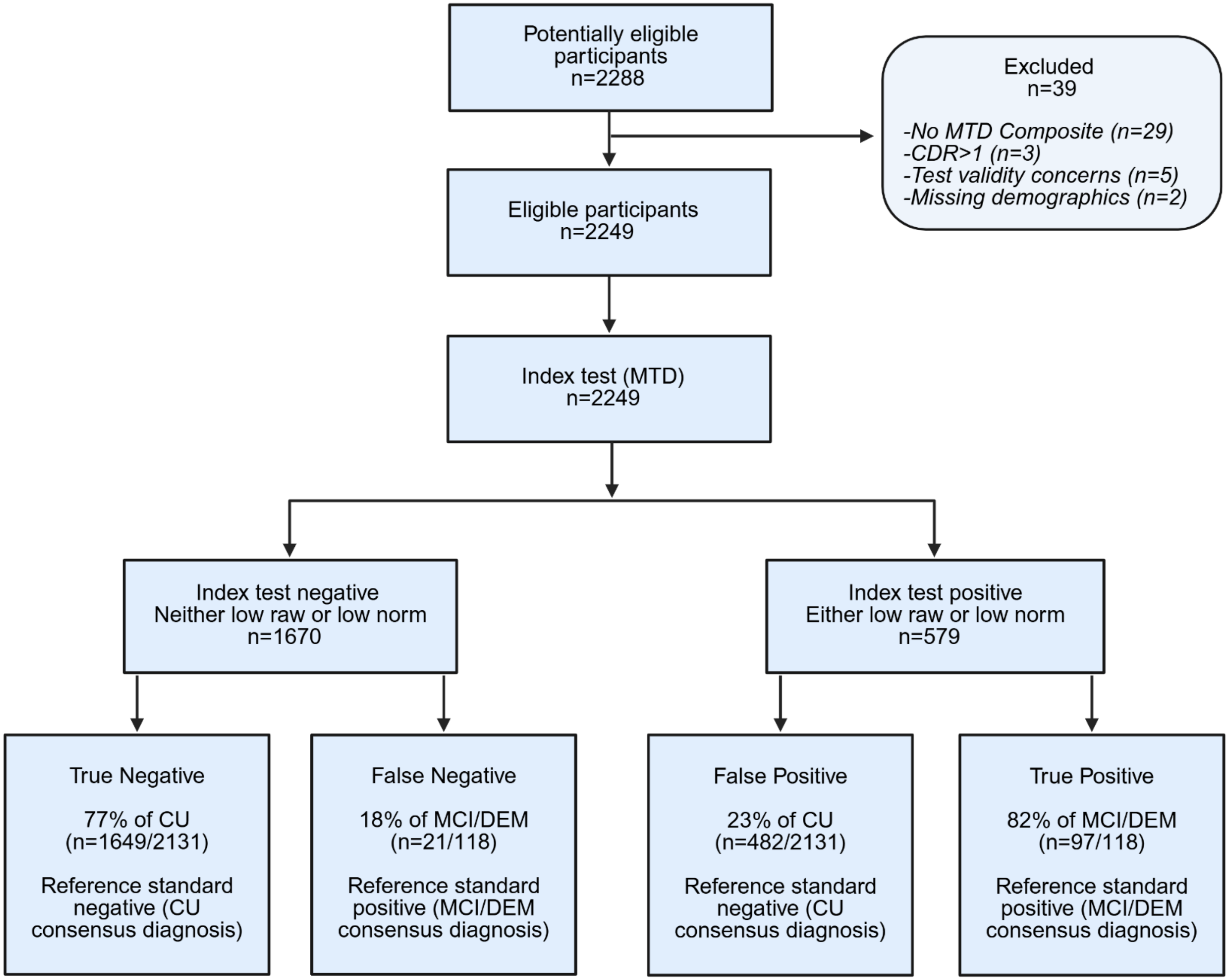


*Note.* Figure with permission of Mayo Foundation for Medical Education and Research; all rights reserved.

**Figure S2.** Sensitivity and specificity of additional MTD subtest scores for differentiating CU vs MCI/mild dementia.

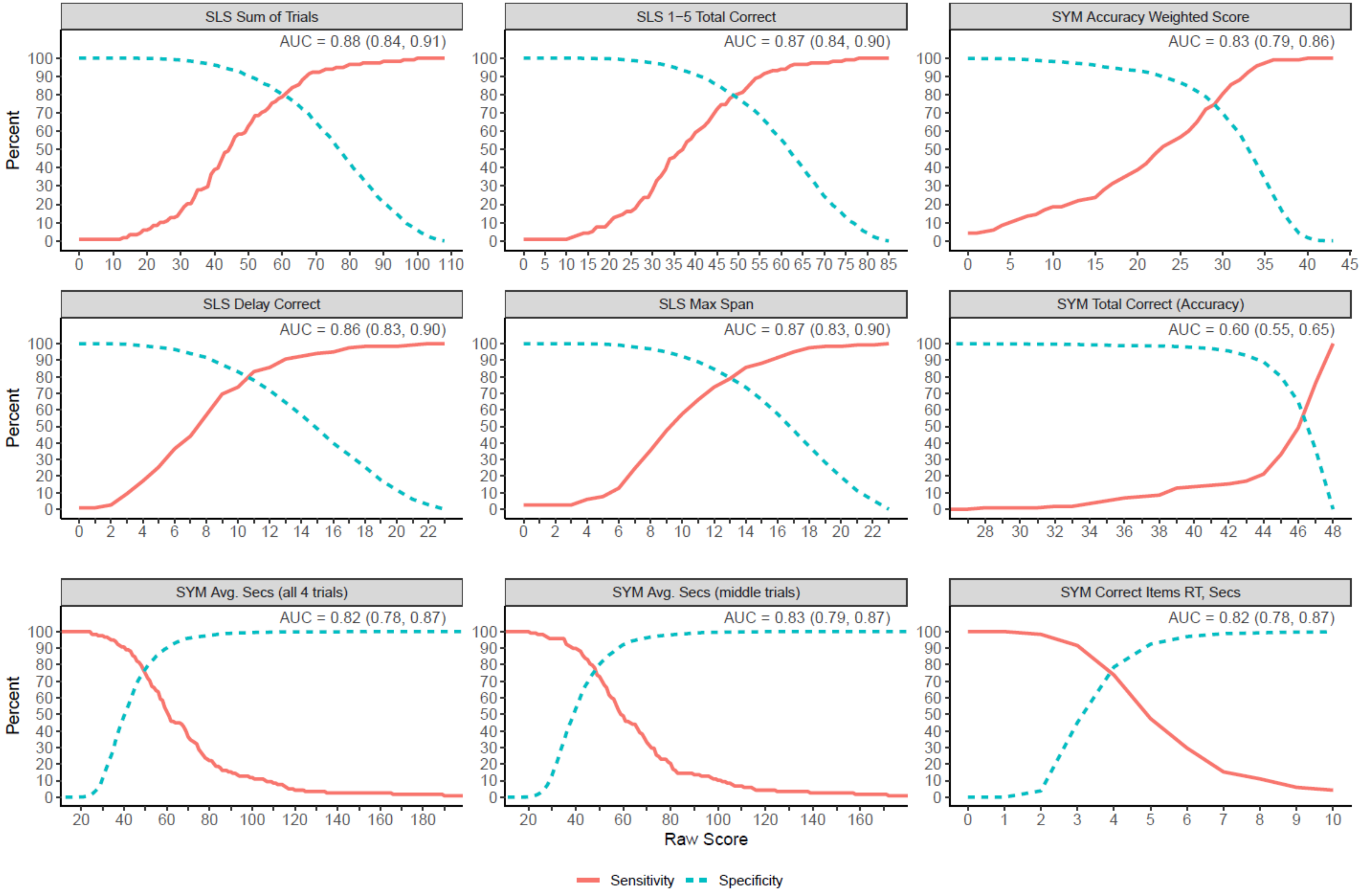

*Note.* AUC = area under the receiver operating characteristic curve; CU = cognitively unimpaired; MCI = mild cognitive impairment; MTD = Mayo Test Drive; SLS = Stricker Learning Span; SLS 1-5 Total = total words correctly recognized across all 5 learning trials; SLS Delay = number of words correctly recognized during the delay trial; SLS Max Span = maximum number of words correctly recognized across any of the 5 learning trials; SLS Sum of Trials = SLS Trials 1-5 Total Correct + SLS Delay Correct; SYM = Symbols Test; SYM Correct Items RT = Symbols Average Response Time (seconds) for Correct Items across all 4 Symbols trials; SYM Avg. Secs = average seconds to complete a trial when averaged across the middle two trials, excluding highest and lowest performances (this measure is included because it may be a more robust option if an interruption disproportionately impacts performance on any single SYM trial). Figure used with permission of Mayo Foundation for Medical Education and Research; all rights reserved.

**Table S1.** *A priori* specified MTD AUCs for MTD raw composite, current observed AUCs (95% Confidence Intervals), and significance of group comparisons.

| Category | Groups | Predicted MTD AUC | Observed MTD AUC | Significance |
|---|---|---|---|---|
| **Clinical Diagnosis** | CU vs MCI/mild dementia [1] | .85 | .89 (.87, .92) | Clinical utility for identifying clinically diagnosed syndromes required for work-up for treatment with monoclonal antibodies |
| **Biomarker-Refined Clinical Diagnosis** | CU A- vs MCI/mild dementia A+ [1] | .85 | .95 (.92, .97) | Clinical utility for MCI/mild dementia due to AD; target population for current treatment with monoclonal antibodies. Other recent digital cognitive diagnostic accuracy studies only report biomarker-refined groups. |
| **Biological AD, independent of diagnosis** | A-T- vs A+T+ | .60 | .76 (.71, .81) | Clinical utility for identifying biological AD. Comparison of remote self-administered MTD to traditional person-administered cognitive measures for differentiating PET biomarker-defined groups, independent of clinical syndrome, to promote use of scalable, remote, self-administered measures. |
| **Preclinical AD** | CU A-T- vs CU A+T+ | .55 | .69 (.62, .75) | Sensitivity to preclinical AD / transitional cognitive decline. Utility for clinical trial enrichment. |

[1] As described in the supplemental method, *a priori*, we hypothesized that the MTD composite would demonstrate excellent discrimination between CU and MCI participants (AUC > .85). Sample size planning targeted approximately 120 MCI participants. If this recruitment target was not achieved, a prespecified alternative analysis combined MCI and mild dementia participants into a broader cognitively impaired group for evaluation of diagnostic accuracy. We implemented this prespecified alternative analysis plan to stay on track with planned analysis timelines for funding and because this also aligns more closely with contemporary monoclonal antibody treatment decision-making pathways, which include both MCI and mild dementia stages. We also provide results for the CU vs MCI comparison.

*Note.* A=brain amyloid (per positron emission tomography, PET); CU=Cognitively Unimpaired; MCI=Mild Cognitive Impairment; T=brain tau (PET). This table is provided to illustrate the *a priori* nature of these analyses as specified in an NIH grant application (1R01AG081955). For these hypotheses specifying a target AUC, if the lower end of the 95% CI did not include the predicted AUC then the null hypothesis was considered rejected and the hypothesis supported. This table is provided in the supplement because AUC values are sample dependent, as evidenced by manuscript Table 2. Table used with permission of Mayo Foundation for Medical Education and Research; all rights reserved.

**Table S2.** Participant demographic, clinical, and neuroimaging characteristics for all participants for participants with versus without amyloid and tau PET imaging.

| Characteristic* | Participants with amyloid & tau PET imaging (N=1224) | Participants without amyloid & tau PET imaging (N=1025) | p-value |
|---|---|---|---|
| Age at MCSA or ADRC visit[1] | 69.0 (11.6) | 70.5 (12.9) | 0.004 |
| Range | 31-98 | 32-101 | |
| Male Sex, N (%) | 590 (48.2%) | 489 (47.7%) | 0.815 |
| Education, years | 15.7 (2.3) | 15.6 (2.3) | 0.354 |
| Range | 6-20 | 8-20 | |
| Race, White, N (%)[1] | 1153 (94.2%) | 952 (92.9%) | 0.098 |
| Ethnicity, Non-Hispanic, N (%)[1] | 1215 (99.3%) | 1009 (98.4%) | 0.045 |
| Non-Hispanic White, N (%) | 1151 (94.0%) | 944 (92.1%) | 0.070 |
| English Second Language, N (%) | 24 (2.0%) | 41 (4.0%) | 0.004 |
| In-person visit to MTD, months | 0.56 (0.51) | 0.60 (0.78) | 0.151 |
| In-person visit to imaging, months | 0.24 (11.62) | - | |
| MTD completed in clinic, N (%) | 10 (0.8%) | 9 (0.9%) | 0.875 |
| Diagnosis, N (%) | | | 0.002 |
| Cognitively Unimpaired | 1144 (93.5%) | 987 (96.3%) | |
| MCI | 61 (5.0%) | 35 (3.4%) | |
| Amnestic[1] | 51 (83.6%) | 27 (77.1%) | |
| Non-amnestic[1] | 8 (13.1%) | 5 (14.3%) | |
| Unknown type | 2 (3.3%) | 3 (8.6%) | |
| Mild Dementia (Probable and Possible AD) | 19 (1.6%) | 3 (0.3%) | |
| CDR Global Score, N (%) | | | 0.078 |
| 0 | 1134 (92.6%) | 965 (94.1%) | |
| 0.5 | 77 (6.3%) | 55 (5.4%) | |
| 1 | 7 (0.6%) | 0 (0.0%) | |
| Unavailable | 6 (0.5%) | 5 (0.5%) | |
| Amyloid PET Centiloid | 28.26 (33.30) | - | |
| Tau PET Meta-ROI SUVR | 1.21 (0.14) | - | |
| MTD Composite Raw Score | 105.5 (22.6) | 104.4 (22.6) | 0.279 |
| Kokmen Short Test of Mental Status | 35.5 (2.5) | 35.5 (2.5) | 0.491 |

| | | | |
|---|---|---|---|
| MMSE/Estimated MMSE from STMS | 28.5 (1.5) | 28.4 (1.4) | 0.342 |
| Mayo-PACC z-score | -0.10 (0.89) | -0.16 (0.86) | 0.100 |
| Global z-score | 0.57 (1.04) | 0.42 (1.04) | 0.001 |

*Mean (standard deviation) except where otherwise noted.
[1] See manuscript Table 1 for additional details.
*Note:* AD = Alzheimer's disease; ADRC = Mayo Clinic Alzheimer's Disease Research Center; CDR = Clinical Dementia Rating Scale; Mayo-PACC z-score = Mayo Preclinical Alzheimer's disease Cognitive Composite z-score (Stricker et al., 2023) calculated on the full baseline MTD dataset in those with a concordant cognitively unimpaired diagnosis; MCI = Mild Cognitive Impairment; MCSA = Mayo Clinic Study of Aging; MMSE = Mini Mental State Examination; MTD = Mayo Test Drive; PET = positron emission tomography; ROI = Region of Interest; STMS = Kokmen Short Test of Mental Status; SUVR = Standard Uptake Volume Ratio. Table used with permission of Mayo Foundation for Medical Education and Research; all rights reserved.

**Table S3.** Logistic regression analyses show the utility of the MTD composite for predicting clinical diagnosis and biomarker-defined groups. The MTD-specific *p*-value for models that include demographics show the independent contribution of MTD beyond age, sex and education.

| *A. Model Performance for Predicting Clinical Diagnosis* | | | | | | |
|---|---|---|---|---|---|---|
| | **A1. CU (n=2131) vs MCI / Mild Dem (n=118)** | | | **A2. CU (n=2131) vs MCI (n=96)** | | |
| Characteristic | Odds Ratio for MTD[1] (95% CI) | *p*-value for MTD | AUC[2] (95% CI) | Odds Ratio for MTD[1] (95% CI) | *p*-value for MTD | AUC[2] (95% CI) |
| **M1:** MTD Composite only | .93 (.92, .94) | <.001 | .893 (.865, .921) | .93 (.92, .94) | <.001 | .883 (.850, .915) |
| **M2:** A/S/E only | - | - | .694 (.649, .740) | - | - | .716 (.667, .766) |
| **M3**: A/S/E + MTD Composite | .93 (.92, .94) | <.001 | .893 (.865, .921) | .94 (.92, .95) | <.001 | .885 (.854, 917) |
| *B. Model Performance for Predicting Biological AD Defined by Abnormal Amyloid PET and Tau PET (A-T- vs A+T+)* | | | | | | |
| | **B.1. All participants (790 A-T- vs 115 A+T+)** | | | **B.2. CU only (764 A-T- vs 80 A+T+)** | | |
| Characteristic | Odds Ratio for MTD[1] (95% CI) | *p*-value for MTD | AUC[2] (95% CI) | Odds Ratio for MTD[1] (95% CI) | *p*-value for MTD | AUC[2] (95% CI) |
| **M1:** MTD Composite only | .96 (.95, .97) | <.001 | .759 (.709, .810) | .97 (.96, .98) | <.001 | .686 (.623, .749) |
| **M2:** A/S/E only | - | - | .820 (.784, .856) | - | - | .837 (.799, .875) |
| **M3**: A/S/E + MTD Comp. | .96 (.95, .97) | <.001 | .857 (.822, .891) | .98 (.97, 1.00) | .02 | .842 (.803, .881) |
| *C. Model Performance for Predicting Abnormal Amyloid (A- vs A+)* | | | | | | |
| | **C.1. All participants (861 A- vs 364 A+)** | | | **C.2. CU only (832 A- vs 313 A+)** | | |
| Characteristic | Odds Ratio for MTD[1] (95% CI) | p-value for MTD | AUC[2] (95% CI) | Odds Ratio for MTD[1] (95% CI) | p-value for MTD | AUC[2] (95% CI) |
| **M1:** MTD Composite only | .97 (.97, .98) | <.001 | .661 (.627, .695) | .98 (.97, .98) | <.001 | .630 (.594, .667) |
| **M2:** A/S/E only | - | - | .764 (.737, .791) | - | - | .768 (.739, .796) |
| **M3**: A/S/E + MTD Comp. | .98 (.98, .99) | <.001 | .777 (.751, .804) | .99 (.98, 1.0) | .04 | .771 (.743, .799) |

[1] MTD odds ratio. Model 1 MTD odds ratio is for MTD only. Model 3 MTD odds ratio adjusts for age, sex, and education. Lower test performance is associated with higher odds of being in the cognitively impaired or biomarker positive group. For example, for the CU vs MCI/DEM comparison (all participants, unadjusted model OR=0.93), each 1-point increase in MTD composite is associated with 7% decreased odds of being MCI/DEM (95% CI 6% - 8%) calculated as $e^{-1*\ln(OR)}$. Because a 1-point change is on a small scale, another example is a -1 SD change using the mean (SD) of the sample - approximately a 23-point increase - is associated with 81% decreased odds of MCI/DEM (95% CI 76% - 85%).

[2] For yes/no responses the AUC is equivalent to concordance, the probability that a randomly selected participant with a yes outcome (cognitively impaired group or biomarker positive group) will have a larger predicted probability than a randomly selected participant with a no outcome (CU group or biomarker negative group).

*Note:* The CU group is the reference group for panel A and the biomarker negative group is the reference group for panels B and C. A- = amyloid PET negative; A+ = amyloid PET positive; AUC = area under the receiver operating characteristic curve; A/S/E = age + sex + education; CI = confidence interval; CU = cognitively unimpaired; Dem = dementia; M = Model; MCI = mild cognitive impairment; MTD = Mayo Test Drive; MTD Comp. = MTD Composite raw score; T- = tau PET negative; T+ = tau PET positive. Table used with permission of Mayo Foundation for Medical Education and Research; all rights reserved.

**Table S4. Model performance of MTD composite compared to in-person cognitive tests in predicting abnormal amyloid and tau PET (A-T- vs A+T+).** Logistic regression analysis predicting A-T- vs. A+T+ for unadjusted models (cognition as the only predictor, on left) and models that also include age, sex, education, in addition to cognition (on right). The *p*-value comparing the AUCs of the remotely administered MTD and in-person cognitive measure provides a direct comparison of test performance for differentiating biomarker-defined groups.

| | **Cognition Only** | | | | **Cognition + Demographics (Age+Sex+Educ)** | | | |
|---|---|---|---|---|---|---|---|---|
| Characteristic | Odds Ratio[1] (95% CI) | *p*-value | AUC[2] (95% CI) | *p*-value comparing AUCs | Cognition Odds Ratio (adj.)[1] (95% CI) | Cog. *p*-value | AUC[2] (95% CI) | *p*-value comparing AUCs |
| **MMSE/est. MMSE (784 A-T- vs 114 A+T+)** | | | | | | | | |
| Demographics only (Age+Sex+Education) | | | | | | | .83 (.79, .86) | |
| MTD Composite | .96 (.95, .97) | <.001 | .76 (.71, .81) | .02 | .97 (.96, .98) | <.001 | .86 (.83, .89) | .30 |
| Est. MMSE | .61 (.54, .69) | <.001 | .70 (.65, .76) | | .68 (.60, .77) | <.001 | .85 (.82, .88) | |
| **STMS (759 A-T- vs 113 A+T+)** | | | | | | | | |
| Demographics only (Age+Sex+Education) | | | | | | | .83 (.79, .86) | |
| MTD Composite | .96 (.95, .97) | <.001 | .76 (.71, .81) | .05 | .97 (.96, .98) | <.001 | .86 (.83, .89) | .20 |
| Kokmen STMS | .75 (.69, .80) | <.001 | .71 (.66, .77) | | .80 (.74, .86) | <.001 | .85 (.82, .89) | |
| **Global-z (705 A-T- vs 86 A+T+)** | | | | | | | | |
| Demographics only (Age+Sex+Education) | | | | | | | .85 (.81, .88) | |
| MTD Composite | .96 (.95, .97) | <.001 | .71 (.65, .77) | .67 | .98 (.96, .99) | .003 | .85 (.81, .89) | .36 |
| Global-z | .47 (.38, .58) | <.001 | .72 (.66, .78) | | .78 (.59, 1.02) | .07 | .85 (.81, .89) | |
| **Mayo-PACC (734 A-T- vs 105 A+T+)** | | | | | | | | |
| Demographics only (Age+Sex+Education) | | | | | | | .83 (.80, .87) | |
| MTD Composite | .96 (.95, .97) | <.001 | .74 (.69, .80) | .71 | .97 (.96, .98) | <.001 | .85 (.82, .89) | .31 |
| Mayo-PACC | .35 (.27, .44) | <.001 | .75 (.70, .81) | | .54 (.40, .71) | <.001 | .85 (.81, .88) | |

[1] Higher odds ratio indicates larger effect, however, MTD and other cognitive measure odds ratios cannot be compared directly with the odds ratio since the scores are on a different scale. AUC data are the focus of test comparisons. Lower test performance is associated with higher odds of being in the biomarker positive group.
[2] For yes/no responses the AUC is equivalent to concordance, the probability that a randomly selected participant with a yes outcome (positive biomarker group) will have a larger predicted probability than a randomly selected participant with a no outcome (negative biomarker group).

*Note.* The biomarker negative group is the reference group (A-T-). A- = amyloid PET negative; A+ = amyloid PET positive; AUC = area under the receiver operating characteristic curve; CI = confidence interval; Educ = education; Global-z = average z-score of 9 in-person-administered neuropsychological measures; STMS = Kokmen Short Test of Mental Status; MMSE = Mini-Mental State Examination, either administered directly or estimated from the STMS; Mayo-PACC = average z-score of AVLT sum of trials + animal fluency + inversed Trails B; MTD = Mayo Test Drive; PET = positron emission tomography; T- = tau PET negative; T+ = tau PET positive. Table used with permission of Mayo Foundation for Medical Education and Research; all rights reserved.

**Table S5.** Subgroup sample sizes, group difference Hedge's G effect sizes, and area under the receiving operating characteristic curves (AUC) with 95% CI for group comparisons for **SLS sum of trials** alone, demographics (age, sex and education) alone, and SLS sum of trials and demographics combined.

| **Outcome** | **Categories** | **n1**[1] | **n2**[2] | **N**[3] | **Effect size**[4] | **SLS AUC**[5] | **Demog. AUC**[6] | **SLS+demog. AUC**[7] | ***p* for SLS**[8] |
|---|---|---|---|---|---|---|---|---|---|
| **a. Clinical Diagnosis** | CU vs MCI/mild DEM | 2131 | 118 | 2249 | -1.66 | .88 (.84, .91) | .69 (.65, .74) | .88 (.85, .91) | <.001 |
| **Subgroup** | CU vs MCI | 2131 | 96 | 2227 | -1.56 | .86 (.82, .90) | .72 (.67, .77) | .87 (.83, .90) | <.001 |
| **Subgroup** | CU vs mild DEM | 2131 | 22 | 2153 | -2.11 | .93 (.89, .98) | .62 (.51, .74) | .93 (.88, .98) | <.001 |
| **Subgroup** | CU A- vs MCI/DEM A+ | 836 | 51 | 887 | -2.19 | .93 (.89, .97) | .80 (.74, .86) | .94 (.90, .98) | <.001 |
| **Subgroup** | CU A- vs MCI A+ | 836 | 34 | 870 | -1.93 | .91 (.85, .96) | .84 (.78, .91) | .92 (.87, .98) | <.001 |
| **b. Biological AD, all** | A-T- vs A+T+ | 790 | 115 | 905 | -1.05 | .74 (.69, .79) | .82 (.78, .86) | .86 (.82, .89) | <.001 |
| **Preclinical AD (CU)** | CU A-T- vs CU A+T+ | 764 | 80 | 844 | -.66 | .66 (.59, .73) | .84 (.80, .88) | .84 (.80, .88) | .007 |
| **AD continuum (CU)** | CU A-T- vs CU A+T- | 764 | 232 | 996 | -.35 | .60 (.55, .64) | .76 (.73, .80) | .77 (.74, .80) | .22 |
| **c. AD path. change** | A- vs A+ | 861 | 364 | 1225 | -.56 | .64 (.61, .68) | .76 (.74, .79) | .78 (.75, .80) | <.001 |
| **AD continuum (CU)** | CU A- vs CU A+ | 836 | 315 | 1151 | -.42 | .61 (.57, .65) | .77 (.74, .80) | .77 (.74, .80) | .01 |

[1] Sample size for the first sample category listed under "Categories."
[2] Sample size for the second sample category listed under "Categories."
[3] Total sample size for the model.
[4] Hedge's G effect size for the group difference in SLS sum of trials raw score (lower performance observed in cognitively impaired and biomarker positive groups).
[5] AUC of the SLS sum of trials; when SLS sum of trials raw score alone is in the model, all *p*'s < .001.
[6] AUC for a model with demographics (age, sex and education) only.
[7] AUC for a model with SLS sum of trials raw score, age, sex, and education.
[8] *p*-value for SLS sum of trials raw score adjusted for age, sex, and education from a logistic regression model.

*Note*. SLS was independent of diagnosis. A- = amyloid PET negative; A+ = amyloid PET positive; AD = Alzheimer's disease; AUC = area under the receiver operating characteristic curve; CI = confidence interval; CU = cognitively unimpaired; DEM = dementia; demog. = demographics; MCI = mild cognitive impairment; path. = pathologic; SLS = Stricker Learning Span sum of trials; T- = tau PET negative; T+ = tau PET positive. Table used with permission of Mayo Foundation for Medical Education and Research; all rights reserved.

**Table S6.** Subgroup sample sizes, group difference Hedge's G effect sizes, and area under the receiving operating characteristic curves (AUC) with 95% CI for group comparisons for **Symbols Test Accuracy Weighted Score (SYM AWS)** alone, demographics (age, sex and education) alone, and SYM AWS and demographics combined.

| **Outcome** | **Categories** | **n1**[1] | **n2**[2] | **N**[3] | **Effect size**[4] | **SYM AUC**[5] | **Demog. AUC**[6] | **SYM+demog. AUC**[7] | ***p* for SYM**[8] |
|---|---|---|---|---|---|---|---|---|---|
| **a. Clinical Diagnosis** | CU vs MCI/mild DEM | 2131 | 118 | 2249 | -1.51 | .83 (.79, .86) | .69 (.65, .74) | .83 (.80, .87) | <.001 |
| **Subgroup** | CU vs MCI | 2131 | 96 | 2227 | -1.43 | .82 (.78, .86) | .72 (.67, .77) | .83 (.79, .86) | <.001 |
| **Subgroup** | CU vs mild DEM | 2131 | 22 | 2153 | -1.98 | .86 (.78, .93) | .62 (.51, .74) | .86 (.79, .94) | <.001 |
| **Subgroup** | CU A- vs MCI/DEM A+ | 836 | 51 | 887 | -2.15 | .90 (.86, .94) | .80 (.74, .86) | .91 (.87, .95) | <.001 |
| **Subgroup** | CU A- vs MCI A+ | 836 | 34 | 870 | -1.99 | .88 (.83, .93) | .84 (.78, .91) | .92 (.87, .98) | <.001 |
| **b. Biological AD, all** | A-T- vs A+T+ | 790 | 115 | 905 | -.99 | .75 (.70, .79) | .82 (.78, .86) | .84 (.81, .87) | <.001 |
| **Preclinical AD (CU)** | CU A-T- vs CU A+T+ | 764 | 80 | 844 | -.60 | .69 (.63, .75) | .84 (.80, .88) | .84 (.80, .87) | .79 |
| **AD continuum (CU)** | CU A-T- vs CU A+T- | 764 | 232 | 996 | -.39 | .62 (.57, .66) | .76 (.73, .80) | .76 (.73, .80) | .31 |
| **c. AD path. change** | A- vs A+ | 861 | 364 | 1225 | -.55 | .66 (.62, .69) | .76 (.74, .79) | .77 (.74, .79) | .03 |
| **AD continuum (CU)** | CU A- vs CU A+ | 836 | 315 | 1151 | -.41 | .63 (.59, .67) | .77 (.74, .80) | .77 (.74, .80) | .49 |

[1] Sample size for the first sample category listed under "Categories."
[2] Sample size for the second sample category listed under "Categories."
[3] Total sample size for the model.
[4] Hedge's G effect size for the group difference in SYM AWS raw score (lower performance observed in cognitively impaired and biomarker positive groups).
[5] AUC of the SYM AWS; when SYM AWS raw score alone is in the model, all *p*'s < .001.
[6] AUC for a model with demographics (age, sex and education) only.
[7] AUC for a model with SYM AWS raw score, age, sex, and education.
[8] *p*-value for SYM AWS raw score adjusted for age, sex, and education from a logistic regression model.

*Note*. The Symbols test was independent of diagnosis. A- = amyloid PET negative; A+ = amyloid PET positive; AD = Alzheimer's disease; AUC = area under the receiver operating characteristic curve; CI = confidence interval; CU = cognitively unimpaired; DEM = dementia; demog. = demographics; MCI = mild cognitive impairment; path. = pathologic; SYM AWS = Symbols Test Accuracy Weighted Score; T- = tau PET negative; T+ = tau PET positive. Table used with permission of Mayo Foundation for Medical Education and Research; all rights reserved.

**Table S7.** Classification accuracy statistics for MTD Composite raw score at various data-derived cutoffs for select group comparisons.

| Group | Cutoff type | ≤ Cutoff | TP | FP | TN | FN | Sensitivity (95% CI) | Specificity (95% CI) |
|---|---|---|---|---|---|---|---|---|
| **CU vs MCI/mild DEM** | **Actual sample sizes** | **Reference** | **118** | **0** | **2131** | **0** | - | - |
| CU vs MCI/mild DEM | Optimal Youden=0.64 | 85 | 92 | 327 | 1804 | 26 | 0.80 (0.73, 0.86) | 0.84 (0.83, 0.86) |
| CU vs MCI/mild DEM | Set 90% sensitivity | 98.28 | 107 | 644 | 1487 | 11 | 0.91 (0.85, 0.96) | 0.70 (0.68, 0.72) |
| CU vs MCI/mild DEM | Set 80% sensitivity | 88.17 | 95 | 396 | 1735 | 23 | 0.81 (0.73, 0.87) | 0.81 (0.80, 0.83) |
| CU vs MCI/mild DEM | Set 90% specificity | 78.35 | 82 | 213 | 1918 | 36 | 0.69 (0.61, 0.77) | 0.90 (0.89, 0.91) |
| CU vs MCI/mild DEM | Set 80% specificity | 89.76 | 97 | 426 | 1705 | 21 | 0.82 (0.75, 0.89) | 0.80 (0.78, 0.82) |
| **CU vs MCI** | **Actual Sample sizes** | **Reference** | **96** | **0** | **2131** | **0** | - | - |
| CU vs MCI | Optimal Youden=0.62 | 90 | 77 | 429 | 1702 | 19 | 0.82 (0.75, 0.90) | 0.80 (0.78, 0.81) |
| CU vs MCI | Set 90% sensitivity | 99.92 | 87 | 694 | 1437 | 9 | 0.91 (0.85, 0.96) | 0.67 (0.66, 0.69) |
| CU vs MCI | Set 80% sensitivity | 89.37 | 77 | 416 | 1715 | 19 | 0.80 (0.73, 0.88) | 0.81 (0.79, 0.82) |
| CU vs MCI | Set 90% specificity | 78.35 | 62 | 213 | 1918 | 34 | 0.65 (0.55, 0.74) | 0.90 (0.89, 0.91) |
| CU vs MCI | Set 80% specificity | 89.76 | 77 | 426 | 1705 | 19 | 0.80 (0.72, 0.88) | 0.80 (0.78, 0.82) |
| **CU A- vs MCI/mild DEM A+** | **Actual Sample sizes** | **Reference** | **51** | **0** | **836** | **0** | | |
| CU A- vs MCI/mild DEM A+ | Optimal Youden=0.74 | 97 | 48 | 181 | 655 | 3 | 0.96 (0.90, 1.00) | 0.78 (0.75, 0.81) |
| CU A- vs MCI/mild DEM A+ | Set 90% sensitivity | 92.26 | 46 | 136 | 700 | 5 | 0.90 (0.82, 0.98) | 0.84 (0.81, 0.86) |
| CU A- vs MCI/mild DEM A+ | Set 80% sensitivity | 82.89 | 41 | 77 | 759 | 10 | 0.80 (0.69, 0.90) | 0.91 (0.89, 0.93) |
| CU A- vs MCI/mild DEM A+ | Set 90% specificity | 84.66 | 42 | 83 | 753 | 9 | 0.82 (0.71, 0.92) | 0.90 (0.88, 0.92) |
| CU A- vs MCI/mild DEM A+ | Set 80% specificity | 95.78 | 47 | 167 | 669 | 4 | 0.92 (0.84, 0.98) | 0.80 (0.77, 0.83) |
| **A-T- vs A+T+** | **Actual Sample sizes** | **Reference** | **115** | **0** | **790** | **0** | | |
| A-T- vs A+T+ | Optimal Youden=0.41 | 97 | 71 | 181 | 609 | 44 | 0.65 (0.57, 0.74) | 0.76 (0.73, 0.79) |
| A-T- vs A+T+ | Set 90% sensitivity | 122.05 | 104 | 557 | 233 | 11 | 0.90 (0.85, 0.96) | 0.29 (0.26, 0.33) |
| A-T- vs A+T+ | Set 80% sensitivity | 111.53 | 92 | 376 | 414 | 23 | 0.80 (0.72, 0.87) | 0.52 (0.49, 0.56) |
| A-T- vs A+T+ | Set 90% specificity | 82.00 | 51 | 79 | 711 | 64 | 0.44 (0.36, 0.54) | 0.90 (0.88, 0.92) |
| A-T- vs A+T+ | Set 80% specificity | 94.67 | 68 | 158 | 632 | 47 | 0.59 (0.50, 0.68) | 0.80 (0.77, 0.83) |
| **CU A-T- vs CU A+T+** | **Actual Sample sizes** | **Reference** | **80** | **0** | **764** | **0** | | |
| CU A-T- vs CU A+T+ | Optimal Youden=0.29 | 112 | 61 | 358 | 406 | 19 | 0.78 (0.69, 0.86) | 0.52 (0.49, 0.55) |
| CU A-T- vs CU A+T+ | Set 90% sensitivity | 125.31 | 72 | 581 | 183 | 8 | 0.90 (0.84, 0.96) | 0.24 (0.21, 0.27) |
| CU A-T- vs CU A+T+ | Set 80% sensitivity | 114.72 | 64 | 409 | 355 | 16 | 0.80 (0.71, 0.89) | 0.47 (0.43, 0.50) |
| CU A-T- vs CU A+T+ | Set 90% specificity | 84.78 | 23 | 76 | 688 | 57 | 0.29 (0.19, 0.39) | 0.90 (0.88, 0.92) |
| CU A-T- vs CU A+T+ | Set 80% specificity | 95.95 | 35 | 152 | 612 | 45 | 0.44 (0.33, 0.55) | 0.80 (0.77, 0.83) |

Note: To rank match Youden's indices to unique, discrete MTD composite scores, model accuracy metrics were applied to the range of possible MTD composite scores (1-150). Each rank below the optimal Youden's J was the highest index value associated with a unique score. Cutoffs were also derived at set values of sensitivity and specificity; the cutoff yielding the value at or greater than the set value is provided. Two by two table information is provided to allow future derivation of any diagnostic accuracy statistics. A- = amyloid PET negative; A+ = amyloid PET positive; CI = confidence interval; CU = cognitively unimpaired; DEM = dementia; FN = false negative; FP = false positive; MCI = mild cognitive impairment; MTD = Mayo Test Drive; T- = tau PET negative; T+ = tau PET positive; TN = true negative; TP = true positive. Table used with permission of Mayo Foundation for Medical Education and Research; all rights reserved.

**Table S8.** Classification accuracy statistics for MTD Composite raw score at post hoc selected candidate raw score cutoffs.

| Group | Cutoff type | < Cutoff | TP | FP | TN | FN | Sensitivity (95% CI) | Specificity (95% CI) |
|---|---|---|---|---|---|---|---|---|
| **CU vs MCI/mild DEM** | **Actual sample sizes** | **Reference** | **118** | **0** | **2131** | **0** | - | - |
| CU vs MCI/mild DEM | Raw score | 80 | 84 | 243 | 1888 | 34 | 0.71 (0.62, 0.79) | 0.89 (0.87, 0.90) |
| CU vs MCI/mild DEM | Raw score | 85 | 92 | 327 | 1804 | 26 | 0.78 (0.69, 0.85) | 0.85 (0.83, 0.86) |
| CU vs MCI/mild DEM | Raw score | 90 | 97 | 429 | 1702 | 21 | 0.82 (0.74, 0.89) | 0.80 (0.78, 0.82) |
| CU vs MCI/mild DEM | Raw score | 95 | 102 | 541 | 1590 | 16 | 0.86 (0.79, 0.92) | 0.75 (0.73, 0.76) |
| CU vs MCI/mild DEM | Raw score | 100 | 108 | 698 | 1433 | 10 | 0.92 (0.85, 0.96) | 0.67 (0.65, 0.69) |
| **CU vs MCI** | **Actual Sample sizes** | **Reference** | **96** | **0** | **2131** | **0** | - | - |
| CU vs MCI | Raw score | 80 | 64 | 243 | 1888 | 32 | 0.67 (0.56, 0.76) | 0.89 (0.87, 0.90) |
| CU vs MCI | Raw score | 85 | 72 | 327 | 1804 | 24 | 0.75 (0.65, 0.83) | 0.85 (0.83, 0.86) |
| CU vs MCI | Raw score | 90 | 77 | 429 | 1702 | 19 | 0.80 (0.71, 0.88) | 0.80 (0.78, 0.82) |
| CU vs MCI | Raw score | 95 | 82 | 541 | 1590 | 14 | 0.85 (0.77, 0.92) | 0.75 (0.73, 0.76) |
| CU vs MCI | Raw score | 100 | 87 | 698 | 1433 | 9 | 0.91 (0.83, 0.96) | 0.67 (0.65, 0.69) |
| **CU A- vs MCI/mild DEM A+** | **Actual Sample sizes** | **Reference** | **51** | **0** | **836** | **0** | | |
| CU A- vs MCI/mild DEM A+ | Raw score | 80 | 39 | 61 | 775 | 12 | 0.76 (0.63, 0.87) | 0.93 (0.91, 0.94) |
| CU A- vs MCI/mild DEM A+ | Raw score | 85 | 42 | 87 | 749 | 9 | 0.82 (0.69, 0.92) | 0.90 (0.87, 0.92) |
| CU A- vs MCI/mild DEM A+ | Raw score | 90 | 44 | 119 | 717 | 7 | 0.86 (0.74, 0.94) | 0.86 (0.83, 0.88) |
| CU A- vs MCI/mild DEM A+ | Raw score | 95 | 46 | 160 | 676 | 5 | 0.90 (0.79, 0.97) | 0.81 (0.78, 0.83) |
| CU A- vs MCI/mild DEM A+ | Raw score | 100 | 49 | 217 | 619 | 2 | 0.96 (0.87, 1.00) | 0.74 (0.71, 0.77) |
| **A-T- vs A+T+** | **Actual Sample sizes** | **Reference** | **115** | **0** | **790** | **0** | | |
| A-T- vs A+T+ | Raw score | 80 | 50 | 69 | 721 | 65 | 0.43 (0.34, 0.53) | 0.91 (0.89, 0.93) |
| A-T- vs A+T+ | Raw score | 85 | 54 | 93 | 697 | 61 | 0.47 (0.38, 0.56) | 0.88 (0.86, 0.90) |
| A-T- vs A+T+ | Raw score | 90 | 63 | 126 | 664 | 52 | 0.55 (0.45, 0.64) | 0.84 (0.81, 0.87) |
| A-T- vs A+T+ | Raw score | 95 | 68 | 165 | 625 | 47 | 0.59 (0.50, 0.68) | 0.79 (0.76, 0.82) |
| A-T- vs A+T+ | Raw score | 100 | 76 | 214 | 576 | 39 | 0.66 (0.57, 0.75) | 0.73 (0.70, 0.76) |
| **CU A-T- vs CU A+T+** | **Actual Sample sizes** | **Reference** | **80** | **0** | **764** | **0** | | |
| CU A-T- vs CU A+T+ | Raw score | 80 | 21 | 54 | 710 | 59 | 0.26 (0.17, 0.37) | 0.93 (0.91, 0.95) |
| CU A-T- vs CU A+T+ | Raw score | 85 | 23 | 77 | 687 | 57 | 0.29 (0.19, 0.40) | 0.90 (0.88, 0.92) |
| CU A-T- vs CU A+T+ | Raw score | 90 | 30 | 109 | 655 | 50 | 0.38 (0.27, 0.49) | 0.86 (0.83, 0.88) |
| CU A-T- vs CU A+T+ | Raw score | 95 | 34 | 146 | 618 | 46 | 0.42 (0.32, 0.54) | 0.81 (0.78, 0.84) |
| CU A-T- vs CU A+T+ | Raw score | 100 | 41 | 192 | 572 | 39 | 0.51 (0.40, 0.63) | 0.75 (0.72, 0.78) |

Note: Select candidate raw score cutoffs were chose after examining Table S7 above. Two by two table information is provided to allow future derivation of any diagnostic accuracy statistics. A- = amyloid PET negative; A+ = amyloid PET positive; CI = confidence interval; CU = cognitively unimpaired; DEM = dementia; FN = false negative; FP = false positive; MCI = mild cognitive impairment; MTD = Mayo Test Drive; T- = tau PET negative; T+ = tau PET positive; TN = true negative; TP = true positive. Table used with permission of Mayo Foundation for Medical Education and Research; all rights reserved.

**Table S9.** Classification accuracy statistics for MTD Composite age/sex/education-adjusted T-scores at various cutoffs.

| Group | Cutoff type | ≤ Cutoff | TP | FP | TN | FN | Sensitivity (95% CI) | Specificity (95% CI) |
|---|---|---|---|---|---|---|---|---|
| **CU vs MCI/mild DEM** | **Actual sample sizes** | **Reference** | **118** | **0** | **2131** | **0** | - | - |
| CU vs MCI/mild DEM | Optimal Youden=0.57 | 42 | 100 | 596 | 1535 | 18 | 0.85 (0.78, 0.92) | 0.72 (0.70, 0.74) |
| CU vs MCI/mild DEM | -1 SD | 40 | 84 | 449 | 1682 | 34 | 0.71 (0.62, 0.79) | 0.79 (0.77, 0.81) |
| CU vs MCI/mild DEM | -1.5 SD | 35 | 65 | 199 | 1932 | 53 | 0.55 (0.46, 0.64) | 0.91 (0.89, 0.92) |
| CU vs MCI/mild DEM | -2 SD | 30 | 42 | 73 | 2058 | 76 | 0.36 (0.27, 0.45) | 0.97 (0.96, 0.97) |
| CU vs MCI/mild DEM | -2.5 SD | 25 | 24 | 21 | 2110 | 94 | 0.20 (0.13, 0.29) | 0.99 (0.98, 0.99) |
| **CU vs MCI** | **Actual Sample sizes** | **Reference** | **96** | **0** | **2131** | **0** | - | - |
| CU vs MCI | Optimal Youden=0.55 | 42 | 80 | 596 | 1535 | 16 | 0.83 (0.75, 0.91) | 0.72 (0.70, 0.74) |
| CU vs MCI | -1 SD | 40 | 66 | 449 | 1682 | 30 | 0.69 (0.58, 0.78) | 0.79 (0.77, 0.81) |
| CU vs MCI | -1.5 SD | 35 | 48 | 199 | 1932 | 48 | 0.50 (0.40, 0.60) | 0.91 (0.89, 0.92) |
| CU vs MCI | -2 SD | 30 | 27 | 73 | 2058 | 69 | 0.28 (0.19, 0.38) | 0.97 (0.96, 0.97) |
| CU vs MCI | -2.5 SD | 25 | 13 | 21 | 2110 | 83 | 0.14 (0.07, 0.22) | 0.99 (0.98, 0.99) |
| **CU A- vs MCI/mild DEM A+** | **Actual Sample sizes** | **Reference** | **51** | **0** | **836** | **0** | | |
| CU A- vs MCI/mild DEM A+ | Optimal Youden=0.67 | 42 | 47 | 212 | 624 | 4 | 0.92 (0.84, 0.98) | 0.75 (0.72, 0.77) |
| CU A- vs MCI/mild DEM A+ | -1 SD | 40 | 41 | 163 | 673 | 10 | 0.80 (0.67, 0.90) | 0.81 (0.78, 0.83) |
| CU A- vs MCI/mild DEM A+ | -1.5 SD | 35 | 34 | 66 | 770 | 17 | 0.67 (0.52, 0.79) | 0.92 (0.90, 0.94) |
| CU A- vs MCI/mild DEM A+ | -2 SD | 30 | 24 | 29 | 807 | 27 | 0.47 (0.33, 0.62) | 0.97 (0.95, 0.98) |
| CU A- vs MCI/mild DEM A+ | -2.5 SD | 25 | 17 | 8 | 828 | 34 | 0.33 (0.21, 0.48) | 0.99 (0.98, 1.00) |
| **A-T- vs A+T+** | **Actual Sample sizes** | **Reference** | **115** | **0** | **790** | **0** | | |
| A-T- vs A+T+ | Optimal Youden=0.31 | 42 | 67 | 214 | 576 | 48 | 0.58 (0.50, 0.67) | 0.73 (0.70, 0.76) |
| A-T- vs A+T+ | -1 SD | 40 | 55 | 166 | 624 | 60 | 0.48 (0.38, 0.57) | 0.79 (0.76, 0.82) |
| A-T- vs A+T+ | -1.5 SD | 35 | 35 | 71 | 719 | 80 | 0.30 (0.22, 0.40) | 0.91 (0.89, 0.93) |
| A-T- vs A+T+ | -2 SD | 30 | 23 | 35 | 755 | 92 | 0.20 (0.13, 0.28) | 0.96 (0.94, 0.97) |
| A-T- vs A+T+ | -2.5 SD | 25 | 17 | 12 | 778 | 98 | 0.15 (0.09, 0.23) | 0.98 (0.97, 0.99) |
| **CU A-T- vs CU A+T+** | **Actual Sample sizes** | **Reference** | **80** | **0** | **764** | **0** | | |
| CU A-T- vs CU A+T+ | Optimal Youden=0.16 | 41 | 31 | 169 | 595 | 49 | 0.39 (0.28, 0.49) | 0.78 (0.75, 0.81) |
| CU A-T- vs CU A+T+ | -1 SD | 40 | 25 | 151 | 613 | 55 | 0.31 (0.21, 0.43) | 0.80 (0.77, 0.83) |
| CU A-T- vs CU A+T+ | -1.5 SD | 35 | 10 | 60 | 704 | 70 | 0.12 (0.06, 0.22) | 0.92 (0.90, 0.94) |
| CU A-T- vs CU A+T+ | -2 SD | 30 | 3 | 28 | 736 | 77 | 0.04 (0.01, 0.11) | 0.96 (0.95, 0.98) |
| CU A-T- vs CU A+T+ | -2.5 SD | 25 | 2 | 8 | 756 | 78 | 0.03 (0.00, 0.09) | 0.99 (0.98, 1.00) |

Note: To rank match Youden's indices to unique, discrete MTD composite scores, model accuracy metrics were applied to the range of possible MTD composite scores (1-100 for T-scores). Each rank below the optimal Youden's J was the highest index value associated with a unique score. Two by two table information is provided to allow future derivation of any diagnostic accuracy statistics. A- = amyloid PET negative; A+ = amyloid PET positive; CI = confidence interval; CU = cognitively unimpaired; DEM = dementia; FN = false negative; FP = false positive; MCI = mild cognitive impairment; MTD = Mayo Test Drive; SD = standard deviation; T- = tau PET negative; T+ = tau PET positive; TN = true negative; TP = true positive. Table used with permission of Mayo Foundation for Medical Education and Research; all rights reserved.

**Table S10.** Classification accuracy statistics for combining raw score and normative score cutoffs: **Mayo Test Drive Composite**.

| Group | Cutoff type | Cutoff | TP | FP | TN | FN | Sensitivity (95% CI) | Specificity (95% CI) |
|---|---|---|---|---|---|---|---|---|
| CU vs MCI/mild DEM | Low Norm, Raw or Both | Any 3 | 97 | 482 | 1649 | 21 | 0.82 (0.74, 0.89) | 0.77 (0.76, 0.79) |
| CU vs MCI/mild DEM | Low Raw | 1. <85 raw | 92 | 327 | 1804 | 26 | 0.78 (0.69, 0.85) | 0.85 (0.83, 0.86) |
| CU vs MCI/mild DEM | Low Norm | 2. <40 T | 80 | 388 | 1743 | 38 | 0.68 (0.59, 0.76) | 0.82 (0.80, 0.83) |
| CU vs MCI/mild DEM | Low Norm and Low Raw | 3. Both | 75 | 233 | 1898 | 43 | 0.64 (0.54, 0.72) | 0.89 (0.88, 0.90) |
| CU vs MCI | Low Norm, Raw or Both | Any 3 | 77 | 482 | 1649 | 19 | 0.80 (0.71, 0.88) | 0.77 (0.76, 0.79) |
| CU vs MCI | Low Raw | 1. <85 raw | 72 | 327 | 1804 | 24 | 0.75 (0.65, 0.83) | 0.85 (0.83, 0.86) |
| CU vs MCI | Low Norm | 2. <40 T | 62 | 388 | 1743 | 34 | 0.65 (0.54, 0.74) | 0.82 (0.80, 0.83) |
| CU vs MCI | Low Norm and Low Raw | 3. Both | 57 | 233 | 1898 | 39 | 0.59 (0.49, 0.69) | 0.89 (0.88, 0.90) |
| CU vs dementia | Low Norm, Raw or Both | Any 3 | 20 | 482 | 1649 | 2 | 0.91 (0.71, 0.99) | 0.77 (0.76, 0.79) |
| CU vs dementia | Low Raw | 1. <85 raw | 20 | 327 | 1804 | 2 | 0.91 (0.71, 0.99) | 0.85 (0.83, 0.86) |
| CU vs dementia | Low Norm | 2. <40 T | 18 | 388 | 1743 | 4 | 0.82 (0.60, 0.95) | 0.82 (0.80, 0.83) |
| CU vs dementia | Low Norm and Low Raw | 3. Both | 18 | 233 | 1898 | 4 | 0.82 (0.60, 0.95) | 0.89 (0.88, 0.90) |
| CU A- vs MCI/mild DEM A+ | Low Norm, Raw or Both | Any 3 | 43 | 158 | 678 | 8 | 0.84 (0.71, 0.93) | 0.81 (0.78, 0.84) |
| CU A- vs MCI/mild DEM A+ | Low Raw | 1. <85 raw | 42 | 87 | 749 | 9 | 0.82 (0.69, 0.92) | 0.90 (0.87, 0.92) |
| CU A- vs MCI/mild DEM A+ | Low Norm | 2. <40 T | 38 | 140 | 696 | 13 | 0.75 (0.60, 0.86) | 0.83 (0.81, 0.86) |
| CU A- vs MCI/mild DEM A+ | Low Norm and Low Raw | 3. Both | 37 | 69 | 767 | 14 | 0.73 (0.58, 0.84) | 0.92 (0.90, 0.94) |
| A-T- vs A+T+ | Low Norm, Raw or Both | Any 3 | 62 | 162 | 628 | 53 | 0.54 (0.44, 0.63) | 0.79 (0.77, 0.82) |
| A-T- vs A+T+ | Low Raw | 1. <85 raw | 54 | 93 | 697 | 61 | 0.47 (0.38, 0.56) | 0.88 (0.86, 0.90) |
| A-T- vs A+T+ | Low Norm | 2. <40 T | 50 | 145 | 645 | 65 | 0.43 (0.34, 0.53) | 0.82 (0.79, 0.84) |
| A-T- vs A+T+ | Low Norm and Low Raw | 3. Both | 42 | 76 | 714 | 73 | 0.37 (0.28, 0.46) | 0.90 (0.88, 0.92) |
| CU A-T- vs CU A+T+ | Low Norm, Raw or Both | Any 3 | 30 | 145 | 619 | 50 | 0.38 (0.27, 0.49) | 0.81 (0.78, 0.84) |
| CU A-T- vs CU A+T+ | Low Raw | 1. <85 raw | 23 | 77 | 687 | 57 | 0.29 (0.19, 0.40) | 0.90 (0.88, 0.92) |
| CU A-T- vs CU A+T+ | Low Norm | 2. <40 T | 22 | 130 | 634 | 58 | 0.28 (0.18, 0.39) | 0.83 (0.80, 0.86) |
| CU A-T- vs CU A+T+ | Low Norm and Low Raw | 3. Both | 15 | 62 | 702 | 65 | 0.19 (0.11, 0.29) | 0.92 (0.90, 0.94) |
| A- vs A+ | Low Norm, Raw or Both | Any 3 | 138 | 178 | 687 | 228 | 0.38 (0.33, 0.43) | 0.79 (0.77, 0.82) |
| A- vs A+ | Low Raw | 1. <85 raw | 119 | 106 | 759 | 247 | 0.33 (0.28, 0.38) | 0.88 (0.85, 0.90) |
| A- vs A+ | Low Norm | 2. <40 T | 105 | 157 | 708 | 261 | 0.29 (0.24, 0.34) | 0.82 (0.79, 0.84) |
| A- vs A+ | Low Norm and Low Raw | 3. Both | 86 | 85 | 780 | 280 | 0.23 (0.19, 0.28) | 0.90 (0.88, 0.92) |
| CU A- vs CU A+ | Low Norm, Raw or Both | Any 3 | 95 | 158 | 678 | 220 | 0.30 (0.25, 0.36) | 0.81 (0.78, 0.84) |
| CU A- vs CU A+ | Low Raw | 1. <85 raw | 77 | 87 | 749 | 238 | 0.24 (0.20, 0.30) | 0.90 (0.87, 0.92) |
| CU A- vs CU A+ | Low Norm | 2. <40 T | 67 | 140 | 696 | 248 | 0.21 (0.17, 0.26) | 0.83 (0.81, 0.86) |
| CU A- vs CU A+ | Low Norm and Low Raw | 3. Both | 49 | 69 | 767 | 266 | 0.16 (0.12, 0.20) | 0.92 (0.90, 0.94) |

*Note*. Normal = MTD composite raw 85+ **and** MTD composite ASE T-score 40+; Low Norm = MTD composite raw 85+ but MTD composite ASE T-score <40; Low Raw = MTD composite ASE T-Score 40+ but MTD composite raw <85; Both Abnormal = MTD composite raw <85 **and** MTD composite ASE T-score <40; Any 3 = either low norm, low raw, or both are met; same as requiring either low raw or low norm; A- = amyloid PET negative; A+ = amyloid PET positive; CI = confidence interval; CU = cognitively unimpaired; DEM = dementia; FN = false negative; FP = false positive; MCI = mild cognitive impairment; MTD = Mayo Test Drive; T- = tau PET negative; T+ = tau PET positive; TN = true negative; TP = true positive. Table used with permission of Mayo Foundation for Medical Education and Research; all rights reserved.

**Table S11**. Classification accuracy statistics for combining raw score and normative score cutoffs: **SLS Sum of Trials**.

| Group | Cutoff type | Cutoff | TP | FP | TN | FN | Sensitivity (95% CI) | Specificity (95% CI) |
|---|---|---|---|---|---|---|---|---|
| CU vs MCI/mild DEM | Low Norm, Raw or Both | Any 3 | 90 | 443 | 1688 | 28 | 0.76 (0.68, 0.84) | 0.79 (0.77, 0.81) |
| CU vs MCI/mild DEM | Low Raw | 1. <57 raw | 86 | 316 | 1815 | 32 | 0.73 (0.64, 0.81) | 0.85 (0.84, 0.87) |
| CU vs MCI/mild DEM | Low Norm | 2. <40 T | 77 | 378 | 1753 | 41 | 0.65 (0.56, 0.74) | 0.82 (0.81, 0.84) |
| CU vs MCI/mild DEM | Low Norm and Low Raw | 3. Both | 73 | 251 | 1880 | 45 | 0.62 (0.52, 0.71) | 0.88 (0.87, 0.90) |
| CU vs MCI | Low Norm, Raw or Both | Any 3 | 71 | 443 | 1688 | 25 | 0.74 (0.64, 0.82) | 0.79 (0.77, 0.81) |
| CU vs MCI | Low Raw | 1. <57 raw | 67 | 316 | 1815 | 29 | 0.70 (0.60, 0.79) | 0.85 (0.84, 0.87) |
| CU vs MCI | Low Norm | 2. <40 T | 60 | 378 | 1753 | 36 | 0.62 (0.52, 0.72) | 0.82 (0.81, 0.84) |
| CU vs MCI | Low Norm and Low Raw | 3. Both | 56 | 251 | 1880 | 40 | 0.58 (0.48, 0.68) | 0.88 (0.87, 0.90) |
| CU vs dementia | Low Norm, Raw or Both | Any 3 | 19 | 443 | 1688 | 3 | 0.86 (0.65, 0.97) | 0.79 (0.77, 0.81) |
| CU vs dementia | Low Raw | 1. <57 raw | 19 | 316 | 1815 | 3 | 0.86 (0.65, 0.97) | 0.85 (0.84, 0.87) |
| CU vs dementia | Low Norm | 2. <40 T | 17 | 378 | 1753 | 5 | 0.77 (0.55, 0.92) | 0.82 (0.81, 0.84) |
| CU vs dementia | Low Norm and Low Raw | 3. Both | 17 | 251 | 1880 | 5 | 0.77 (0.55, 0.92) | 0.88 (0.87, 0.90) |
| CU A- vs MCI/mild DEM A+ | Low Norm, Raw or Both | Any 3 | 40 | 146 | 690 | 11 | 0.78 (0.65, 0.89) | 0.83 (0.80, 0.85) |
| CU A- vs MCI/mild DEM A+ | Low Raw | 1. <57 raw | 40 | 89 | 747 | 11 | 0.78 (0.65, 0.89) | 0.89 (0.87, 0.91) |
| CU A- vs MCI/mild DEM A+ | Low Norm | 2. <40 T | 38 | 130 | 706 | 13 | 0.75 (0.60, 0.86) | 0.84 (0.82, 0.87) |
| CU A- vs MCI/mild DEM A+ | Low Norm and Low Raw | 3. Both | 38 | 73 | 763 | 13 | 0.75 (0.60, 0.86) | 0.91 (0.89, 0.93) |
| A-T- vs A+T+ | Low Norm, Raw or Both | Any 3 | 57 | 153 | 637 | 58 | 0.50 (0.40, 0.59) | 0.81 (0.78, 0.83) |
| A-T- vs A+T+ | Low Raw | 1. <57 raw | 52 | 96 | 694 | 63 | 0.45 (0.36, 0.55) | 0.88 (0.85, 0.90) |
| A-T- vs A+T+ | Low Norm | 2. <40 T | 51 | 136 | 654 | 64 | 0.44 (0.35, 0.54) | 0.83 (0.80, 0.85) |
| A-T- vs A+T+ | Low Norm and Low Raw | 3. Both | 46 | 79 | 711 | 69 | 0.40 (0.31, 0.50) | 0.90 (0.88, 0.92) |
| CU A-T- vs CU A+T+ | Low Norm, Raw or Both | Any 3 | 28 | 135 | 629 | 52 | 0.35 (0.25, 0.46) | 0.82 (0.79, 0.85) |
| CU A-T- vs CU A+T+ | Low Raw | 1. <57 raw | 23 | 81 | 683 | 57 | 0.29 (0.19, 0.40) | 0.89 (0.87, 0.91) |
| CU A-T- vs CU A+T+ | Low Norm | 2. <40 T | 23 | 123 | 641 | 57 | 0.29 (0.19, 0.40) | 0.84 (0.81, 0.86) |
| CU A-T- vs CU A+T+ | Low Norm and Low Raw | 3. Both | 18 | 69 | 695 | 62 | 0.23 (0.14, 0.33) | 0.91 (0.89, 0.93) |
| A- vs A+ | Low Norm, Raw or Both | Any 3 | 129 | 167 | 698 | 237 | 0.35 (0.30, 0.40) | 0.81 (0.78, 0.83) |
| A- vs A+ | Low Raw | 1. <57 raw | 112 | 107 | 758 | 254 | 0.31 (0.26, 0.36) | 0.88 (0.85, 0.90) |
| A- vs A+ | Low Norm | 2. <40 T | 110 | 145 | 720 | 256 | 0.30 (0.25, 0.35) | 0.83 (0.81, 0.86) |
| A- vs A+ | Low Norm and Low Raw | 3. Both | 93 | 85 | 780 | 273 | 0.25 (0.21, 0.30) | 0.90 (0.88, 0.92) |
| CU A- vs CU A+ | Low Norm, Raw or Both | Any 3 | 89 | 146 | 690 | 226 | 0.28 (0.23, 0.34) | 0.83 (0.80, 0.85) |
| CU A- vs CU A+ | Low Raw | 1. <57 raw | 72 | 89 | 747 | 243 | 0.23 (0.18, 0.28) | 0.89 (0.87, 0.91) |
| CU A- vs CU A+ | Low Norm | 2. <40 T | 72 | 130 | 706 | 243 | 0.23 (0.18, 0.28) | 0.84 (0.82, 0.87) |
| CU A- vs CU A+ | Low Norm and Low Raw | 3. Both | 55 | 73 | 763 | 260 | 0.17 (0.13, 0.22) | 0.91 (0.89, 0.93) |

*Note*. Normal = SLS sum of trials raw 57+ **and** SLS Sum of Trials ASE T-score 40+; Low Norm = SLS sum of trials raw 57+ but SLS Sum of Trials ASE T-score <40; Low Raw = SLS Sum of Trials T-Score 40+ but SLS Sum of Trials raw <57, this raw score corresponds to an unadjusted scaled score < 7 in the full normative sample; Both Abnormal = SLS sum of trials raw < 57 **and** SLS Sum of Trials ASE T-score <40; Any 3 = either low norm, low raw, or both are met; same as requiring either low raw or low norm; A- = amyloid PET negative; A+ = amyloid PET positive; CI = confidence interval; CU = cognitively unimpaired; DEM = dementia; FN = false negative; FP = false positive; MCI = mild cognitive impairment; MTD = Mayo Test Drive; T- = tau PET negative; T+ = tau PET positive; TN = true negative; TP = true positive. Table used with permission of Mayo Foundation for Medical Education and Research; all rights reserved.

**Table S12**. Classification accuracy statistics for combining raw score and normative score cutoffs: **SYM Accuracy-Weighted Score**.

| Group | Cutoff type | Cutoff | TP | FP | TN | FN | Sensitivity (95% CI) | Specificity (95% CI) |
|---|---|---|---|---|---|---|---|---|
| CU vs MCI/mild DEM | Low Norm, Raw or Both | Any 3 | 82 | 431 | 1700 | 36 | 0.69 (0.60, 0.78) | 0.80 (0.78, 0.81) |
| CU vs MCI/mild DEM | Low Raw | 1. <25.5 raw | 70 | 304 | 1827 | 48 | 0.59 (0.50, 0.68) | 0.86 (0.84, 0.87) |
| CU vs MCI/mild DEM | Low Norm | 2. <40 T | 63 | 334 | 1797 | 55 | 0.53 (0.44, 0.63) | 0.84 (0.83, 0.86) |
| CU vs MCI/mild DEM | Low Norm and Low Raw | 3. Both | 51 | 207 | 1924 | 67 | 0.43 (0.34, 0.53) | 0.90 (0.89, 0.92) |
| CU vs MCI | Low Norm, Raw or Both | Any 3 | 64 | 431 | 1700 | 32 | 0.67 (0.56, 0.76) | 0.80 (0.78, 0.81) |
| CU vs MCI | Low Raw | 1. <25.5 raw | 55 | 304 | 1827 | 41 | 0.57 (0.47, 0.67) | 0.86 (0.84, 0.87) |
| CU vs MCI | Low Norm | 2. <40 T | 46 | 334 | 1797 | 50 | 0.48 (0.38, 0.58) | 0.84 (0.83, 0.86) |
| CU vs MCI | Low Norm and Low Raw | 3. Both | 37 | 207 | 1924 | 59 | 0.39 (0.29, 0.49) | 0.90 (0.89, 0.92) |
| CU vs dementia | Low Norm, Raw or Both | Any 3 | 18 | 431 | 1700 | 4 | 0.82 (0.60, 0.95) | 0.80 (0.78, 0.81) |
| CU vs dementia | Low Raw | 1. <25.5 raw | 15 | 304 | 1827 | 7 | 0.68 (0.45, 0.86) | 0.86 (0.84, 0.87) |
| CU vs dementia | Low Norm | 2. <40 T | 17 | 334 | 1797 | 5 | 0.77 (0.55, 0.92) | 0.84 (0.83, 0.86) |
| CU vs dementia | Low Norm and Low Raw | 3. Both | 14 | 207 | 1924 | 8 | 0.64 (0.41, 0.83) | 0.90 (0.89, 0.92) |
| CU A- vs MCI/mild DEM A+ | Low Norm, Raw or Both | Any 3 | 37 | 140 | 696 | 14 | 0.73 (0.58, 0.84) | 0.83 (0.81, 0.86) |
| CU A- vs MCI/mild DEM A+ | Low Raw | 1. <25.5 raw | 32 | 76 | 760 | 19 | 0.63 (0.48, 0.76) | 0.91 (0.89, 0.93) |
| CU A- vs MCI/mild DEM A+ | Low Norm | 2. <40 T | 31 | 122 | 714 | 20 | 0.61 (0.46, 0.74) | 0.85 (0.83, 0.88) |
| CU A- vs MCI/mild DEM A+ | Low Norm and Low Raw | 3. Both | 26 | 58 | 778 | 25 | 0.51 (0.37, 0.65) | 0.93 (0.91, 0.95) |
| A-T- vs A+T+ | Low Norm, Raw or Both | Any 3 | 46 | 145 | 645 | 69 | 0.40 (0.31, 0.50) | 0.82 (0.79, 0.84) |
| A-T- vs A+T+ | Low Raw | 1. <25.5 raw | 40 | 79 | 711 | 75 | 0.35 (0.26, 0.44) | 0.90 (0.88, 0.92) |
| A-T- vs A+T+ | Low Norm | 2. <40 T | 36 | 127 | 663 | 79 | 0.31 (0.23, 0.41) | 0.84 (0.81, 0.86) |
| A-T- vs A+T+ | Low Norm and Low Raw | 3. Both | 30 | 61 | 729 | 85 | 0.26 (0.18, 0.35) | 0.92 (0.90, 0.94) |
| CU A-T- vs CU A+T+ | Low Norm, Raw or Both | Any 3 | 19 | 130 | 634 | 61 | 0.24 (0.15, 0.35) | 0.83 (0.80, 0.86) |
| CU A-T- vs CU A+T+ | Low Raw | 1. <25.5 raw | 17 | 68 | 696 | 63 | 0.21 (0.13, 0.32) | 0.91 (0.89, 0.93) |
| CU A-T- vs CU A+T+ | Low Norm | 2. <40 T | 12 | 115 | 649 | 68 | 0.15 (0.13, 0.18) | 0.85 (0.75, 0.92) |
| CU A-T- vs CU A+T+ | Low Norm and Low Raw | 3. Both | 10 | 53 | 711 | 70 | 0.12 (0.06, 0.22) | 0.93 (0.91, 0.95) |
| A- vs A+ | Low Norm, Raw or Both | Any 3 | 109 | 156 | 709 | 257 | 0.30 (0.25, 0.35) | 0.82 (0.79, 0.84) |
| A- vs A+ | Low Raw | 1. <25.5 raw | 93 | 88 | 777 | 273 | 0.25 (0.21, 0.30) | 0.90 (0.88, 0.92) |
| A- vs A+ | Low Norm | 2. <40 T | 77 | 134 | 731 | 289 | 0.21 (0.17, 0.26) | 0.85 (0.82, 0.87) |
| A- vs A+ | Low Norm and Low Raw | 3. Both | 61 | 66 | 799 | 305 | 0.17 (0.13, 0.21) | 0.92 (0.90, 0.94) |
| CU A- vs CU A+ | Low Norm, Raw or Both | Any 3 | 72 | 140 | 696 | 243 | 0.23 (0.18, 0.28) | 0.83 (0.81, 0.86) |
| CU A- vs CU A+ | Low Raw | 1. <25.5 raw | 61 | 76 | 760 | 254 | 0.19 (0.15, 0.24) | 0.91 (0.89, 0.93) |
| CU A- vs CU A+ | Low Norm | 2. <40 T | 46 | 122 | 714 | 269 | 0.15 (0.11, 0.19) | 0.85 (0.83, 0.88) |
| CU A- vs CU A+ | Low Norm and Low Raw | 3. Both | 35 | 58 | 778 | 280 | 0.11 (0.08, 0.15) | 0.93 (0.91, 0.95) |

*Note.* Normal = SYM AWS 25.5+ **and** SYM AWS ASE T-score 40+; Low Norm = SYM AWS 25.5+ but SYM AWS ASE T-score <40; Low Raw = SYM AWS T-Score 40+ but SYM AWS raw < 25.5, this raw score corresponds to an unadjusted scaled score < 7 in the full normative sample; Both Abnormal = SYM AWS raw < 25.5 **and** SYM AWS ASE T-score <40; Any 3 = either low norm, low raw, or both are met; same as requiring either low raw or low norm; A- = amyloid PET negative; A+ = amyloid PET positive; CI = confidence interval; CU = cognitively unimpaired; DEM = dementia; FN = false negative; FP = false positive; MCI = mild cognitive impairment; MTD = Mayo Test Drive; T- = tau PET negative; T+ = tau PET positive; TN = true negative; TP = true positive. Table used with permission of Mayo Foundation for Medical Education and Research; all rights reserved.

**Table S13**. Classification accuracy statistics for combining raw score and normative score cutoffs: **SLS Max Span**.

| Group | Cutoff type | Cutoff | TP | FP | TN | FN | Sensitivity (95% CI) | Specificity (95% CI) |
|---|---|---|---|---|---|---|---|---|
| CU vs MCI/mild DEM | Low Norm, Raw or Both | Any 3 | 91 | 436 | 1695 | 27 | 0.77 (0.68, 0.84) | 0.80 (0.78, 0.81) |
| CU vs MCI/mild DEM | Low Raw | 1. <13 raw | 87 | 329 | 1802 | 31 | 0.74 (0.65, 0.81) | 0.85 (0.83, 0.86) |
| CU vs MCI/mild DEM | Low Norm | 2. <40 T | 76 | 360 | 1771 | 42 | 0.64 (0.55, 0.73) | 0.83 (0.81, 0.85) |
| CU vs MCI/mild DEM | Low Norm and Low Raw | 3. Both | 72 | 253 | 1878 | 46 | 0.61 (0.52, 0.70) | 0.88 (0.87, 0.89) |
| CU vs MCI | Low Norm, Raw or Both | Any 3 | 72 | 436 | 1695 | 24 | 0.75 (0.65, 0.83) | 0.80 (0.78, 0.81) |
| CU vs MCI | Low Raw | 1. <13 raw | 68 | 329 | 1802 | 28 | 0.71 (0.61, 0.80) | 0.85 (0.83, 0.86) |
| CU vs MCI | Low Norm | 2. <40 T | 59 | 360 | 1771 | 37 | 0.61 (0.51, 0.71) | 0.83 (0.81, 0.85) |
| CU vs MCI | Low Norm and Low Raw | 3. Both | 55 | 253 | 1878 | 41 | 0.57 (0.47, 0.67) | 0.88 (0.87, 0.89) |
| CU vs dementia | Low Norm, Raw or Both | Any 3 | 19 | 436 | 1695 | 3 | 0.86 (0.65, 0.97) | 0.80 (0.78, 0.81) |
| CU vs dementia | Low Raw | 1. <13 raw | 19 | 329 | 1802 | 3 | 0.86 (0.65, 0.97) | 0.85 (0.83, 0.86) |
| CU vs dementia | Low Norm | 2. <40 T | 17 | 360 | 1771 | 5 | 0.77 (0.55, 0.92) | 0.83 (0.81, 0.85) |
| CU vs dementia | Low Norm and Low Raw | 3. Both | 17 | 253 | 1878 | 5 | 0.77 (0.55, 0.92) | 0.88 (0.87, 0.89) |
| CU A- vs MCI/mild DEM A+ | Low Norm, Raw or Both | Any 3 | 41 | 146 | 690 | 10 | 0.80 (0.67, 0.90) | 0.83 (0.80, 0.85) |
| CU A- vs MCI/mild DEM A+ | Low Raw | 1. <13 raw | 40 | 95 | 741 | 11 | 0.78 (0.65, 0.89) | 0.89 (0.86, 0.91) |
| CU A- vs MCI/mild DEM A+ | Low Norm | 2. <40 T | 36 | 126 | 710 | 15 | 0.71 (0.56, 0.83) | 0.85 (0.82, 0.87) |
| CU A- vs MCI/mild DEM A+ | Low Norm and Low Raw | 3. Both | 35 | 75 | 761 | 16 | 0.69 (0.54, 0.81) | 0.91 (0.89, 0.93) |
| A-T- vs A+T+ | Low Norm, Raw or Both | Any 3 | 57 | 154 | 636 | 58 | 0.50 (0.40, 0.59) | 0.81 (0.78, 0.83) |
| A-T- vs A+T+ | Low Raw | 1. <13 raw | 51 | 104 | 686 | 64 | 0.44 (0.35, 0.54) | 0.87 (0.84, 0.89) |
| A-T- vs A+T+ | Low Norm | 2. <40 T | 49 | 133 | 657 | 66 | 0.43 (0.33, 0.52) | 0.83 (0.80, 0.86) |
| A-T- vs A+T+ | Low Norm and Low Raw | 3. Both | 43 | 83 | 707 | 72 | 0.37 (0.29, 0.47) | 0.89 (0.87, 0.92) |
| CU A-T- vs CU A+T+ | Low Norm, Raw or Both | Any 3 | 28 | 136 | 628 | 52 | 0.35 (0.25, 0.46) | 0.82 (0.79, 0.85) |
| CU A-T- vs CU A+T+ | Low Raw | 1. <13 raw | 23 | 87 | 677 | 57 | 0.29 (0.19, 0.40) | 0.89 (0.86, 0.91) |
| CU A-T- vs CU A+T+ | Low Norm | 2. <40 T | 22 | 119 | 645 | 58 | 0.28 (0.18, 0.39) | 0.84 (0.82, 0.87) |
| CU A-T- vs CU A+T+ | Low Norm and Low Raw | 3. Both | 17 | 70 | 694 | 63 | 0.21 (0.13, 0.32) | 0.91 (0.89, 0.93) |
| A- vs A+ | Low Norm, Raw or Both | Any 3 | 131 | 167 | 698 | 235 | 0.36 (0.31, 0.41) | 0.81 (0.78, 0.83) |
| A- vs A+ | Low Raw | 1. <13 raw | 113 | 115 | 750 | 253 | 0.31 (0.26, 0.36) | 0.87 (0.84, 0.89) |
| A- vs A+ | Low Norm | 2. <40 T | 106 | 143 | 722 | 260 | 0.29 (0.24, 0.34) | 0.83 (0.81, 0.86) |
| A- vs A+ | Low Norm and Low Raw | 3. Both | 88 | 91 | 774 | 278 | 0.24 (0.20, 0.29) | 0.89 (0.87, 0.91) |
| CU A- vs CU A+ | Low Norm, Raw or Both | Any 3 | 90 | 146 | 690 | 225 | 0.29 (0.24, 0.34) | 0.83 (0.80, 0.85) |
| CU A- vs CU A+ | Low Raw | 1. <13 raw | 73 | 95 | 741 | 242 | 0.23 (0.19, 0.28) | 0.89 (0.86, 0.91) |
| CU A- vs CU A+ | Low Norm | 2. <40 T | 70 | 126 | 710 | 245 | 0.22 (0.18, 0.27) | 0.85 (0.82, 0.87) |
| CU A- vs CU A+ | Low Norm and Low Raw | 3. Both | 53 | 75 | 761 | 262 | 0.17 (0.13, 0.21) | 0.91 (0.89, 0.93) |

*Note.* Normal = SLS Max Span 13+ **and** SLS Max Span ASE T-score 40+; Low Norm = SLS Max Span 13+ but SLS Max Span ASE T-score <40; Low Raw = SLS Max Span T-Score 40+ but SLS Max Span raw < 13, this raw score corresponds to an unadjusted scaled score < 7 in the full normative sample; Both Abnormal: SLS Max Span raw < 13 **and** SLS Max Span ASE T-score <40; Any 3 = either low norm, low raw, or both are met; same as requiring either low raw or low norm; A- = amyloid PET negative; A+ = amyloid PET positive; CI = confidence interval; CU = cognitively unimpaired; DEM = dementia; FN = false negative; FP = false positive; MCI = mild cognitive impairment; MTD = Mayo Test Drive; T- = tau PET negative; T+ = tau PET positive; TN = true negative; TP = true positive. Table used with permission of Mayo Foundation for Medical Education and Research; all rights reserved.

**Table S14**. Classification accuracy statistics for combining raw score and normative score cutoffs: **SLS Delay**.

| Group | Cutoff type | Cutoff | TP | FP | TN | FN | Sensitivity (95% CI) | Specificity (95% CI) |
|---|---|---|---|---|---|---|---|---|
| CU vs MCI/mild DEM | Low Norm, Raw or Both | Any 3 | 90 | 469 | 1662 | 28 | 0.76 (0.68, 0.84) | 0.78 (0.76, 0.80) |
| CU vs MCI/mild DEM | Low Raw | 1. <11 raw | 87 | 363 | 1768 | 31 | 0.74 (0.65, 0.81) | 0.83 (0.81, 0.85) |
| CU vs MCI/mild DEM | Low Norm | 2. <40 T | 75 | 381 | 1750 | 43 | 0.64 (0.54, 0.72) | 0.82 (0.80, 0.84) |
| CU vs MCI/mild DEM | Low Norm and Low Raw | 3. Both | 72 | 275 | 1856 | 46 | 0.61 (0.52, 0.70) | 0.87 (0.86, 0.88) |
| CU vs MCI | Low Norm, Raw or Both | Any 3 | 71 | 469 | 1662 | 25 | 0.74 (0.64, 0.82) | 0.78 (0.76, 0.80) |
| CU vs MCI | Low Raw | 1. <11 raw | 68 | 363 | 1768 | 28 | 0.71 (0.61, 0.80) | 0.83 (0.81, 0.85) |
| CU vs MCI | Low Norm | 2. <40 T | 58 | 381 | 1750 | 38 | 0.60 (0.50, 0.70) | 0.82 (0.80, 0.84) |
| CU vs MCI | Low Norm and Low Raw | 3. Both | 55 | 275 | 1856 | 41 | 0.57 (0.47, 0.67) | 0.87 (0.86, 0.88) |
| CU vs dementia | Low Norm, Raw or Both | Any 3 | 19 | 469 | 1662 | 3 | 0.86 (0.65, 0.97) | 0.78 (0.76, 0.80) |
| CU vs dementia | Low Raw | 1. <11 raw | 19 | 363 | 1768 | 3 | 0.86 (0.65, 0.97) | 0.83 (0.81, 0.85) |
| CU vs dementia | Low Norm | 2. <40 T | 17 | 381 | 1750 | 5 | 0.77 (0.55, 0.92) | 0.82 (0.80, 0.84) |
| CU vs dementia | Low Norm and Low Raw | 3. Both | 17 | 275 | 1856 | 5 | 0.77 (0.55, 0.92) | 0.87 (0.86, 0.88) |
| CU A- vs MCI/mild DEM A+ | Low Norm, Raw or Both | Any 3 | 43 | 152 | 684 | 8 | 0.84 (0.71, 0.93) | 0.82 (0.79, 0.84) |
| CU A- vs MCI/mild DEM A+ | Low Raw | 1. <11 raw | 42 | 111 | 725 | 9 | 0.82 (0.69, 0.92) | 0.87 (0.84, 0.89) |
| CU A- vs MCI/mild DEM A+ | Low Norm | 2. <40 T | 41 | 130 | 706 | 10 | 0.80 (0.67, 0.90) | 0.84 (0.82, 0.87) |
| CU A- vs MCI/mild DEM A+ | Low Norm and Low Raw | 3. Both | 40 | 89 | 747 | 11 | 0.78 (0.65, 0.89) | 0.89 (0.87, 0.91) |
| A-T- vs A+T+ | Low Norm, Raw or Both | Any 3 | 60 | 154 | 636 | 55 | 0.52 (0.43, 0.62) | 0.81 (0.78, 0.83) |
| A-T- vs A+T+ | Low Raw | 1. <11 raw | 56 | 113 | 677 | 59 | 0.49 (0.39, 0.58) | 0.86 (0.83, 0.88) |
| A-T- vs A+T+ | Low Norm | 2. <40 T | 49 | 130 | 660 | 66 | 0.43 (0.33, 0.52) | 0.84 (0.81, 0.86) |
| A-T- vs A+T+ | Low Norm and Low Raw | 3. Both | 45 | 89 | 701 | 70 | 0.39 (0.30, 0.49) | 0.89 (0.86, 0.91) |
| CU A-T- vs CU A+T+ | Low Norm, Raw or Both | Any 3 | 28 | 138 | 626 | 52 | 0.35 (0.25, 0.46) | 0.82 (0.79, 0.85) |
| CU A-T- vs CU A+T+ | Low Raw | 1. <11 raw | 25 | 99 | 665 | 55 | 0.31 (0.21, 0.43) | 0.87 (0.84, 0.89) |
| CU A-T- vs CU A+T+ | Low Norm | 2. <40 T | 19 | 120 | 644 | 61 | 0.24 (0.15, 0.35) | 0.84 (0.82, 0.87) |
| CU A-T- vs CU A+T+ | Low Norm and Low Raw | 3. Both | 16 | 81 | 683 | 64 | 0.20 (0.12, 0.30) | 0.89 (0.87, 0.91) |
| A- vs A+ | Low Norm, Raw or Both | Any 3 | 138 | 170 | 695 | 228 | 0.38 (0.33, 0.43) | 0.80 (0.78, 0.83) |
| A- vs A+ | Low Raw | 1. <11 raw | 123 | 127 | 738 | 243 | 0.34 (0.29, 0.39) | 0.85 (0.83, 0.88) |
| A- vs A+ | Low Norm | 2. <40 T | 113 | 142 | 723 | 253 | 0.31 (0.26, 0.36) | 0.84 (0.81, 0.86) |
| A- vs A+ | Low Norm and Low Raw | 3. Both | 98 | 99 | 766 | 268 | 0.27 (0.22, 0.32) | 0.89 (0.86, 0.91) |
| CU A- vs CU A+ | Low Norm, Raw or Both | Any 3 | 95 | 152 | 684 | 220 | 0.30 (0.25, 0.36) | 0.82 (0.79, 0.84) |
| CU A- vs CU A+ | Low Raw | 1. <11 raw | 81 | 111 | 725 | 234 | 0.26 (0.21, 0.31) | 0.87 (0.84, 0.89) |
| CU A- vs CU A+ | Low Norm | 2. <40 T | 72 | 130 | 706 | 243 | 0.23 (0.18, 0.28) | 0.84 (0.82, 0.87) |
| CU A- vs CU A+ | Low Norm and Low Raw | 3. Both | 58 | 89 | 747 | 257 | 0.18 (0.14, 0.23) | 0.89 (0.87, 0.91) |

*Note.* Normal = SLS Delay raw 11+ **and** SLS Delay ASE T-score 40+; Low Norm = SLS Delay raw 11+ but SLS Delay ASE T-score <40; Low Raw = SLS Delay T-Score 40+ but SLS Delay raw < 11, this raw score corresponds to an unadjusted scaled score < 7 in the full normative sample; Both Abnormal = SLS Delay raw < 11 **and** SLS Delay ASE T-score <40; Any 3 = either low norm, low raw, or both are met; same as requiring either low raw or low norm; A- = amyloid PET negative; A+ = amyloid PET positive; CI = confidence interval; CU = cognitively unimpaired; DEM = dementia; FN = false negative; FP = false positive; MCI = mild cognitive impairment; MTD = Mayo Test Drive; T- = tau PET negative; T+ = tau PET positive; TN = true negative; TP = true positive. Table used with permission of Mayo Foundation for Medical Education and Research; all rights reserved.

**Table S15**. Classification accuracy statistics for combining raw score and normative score cutoffs: SYM Average Seconds (Middle 2 Trials).

| Group | Cutoff type | Cutoff | TP | FP | TN | FN | Sensitivity (95% CI) | Specificity (95% CI) |
|---|---|---|---|---|---|---|---|---|
| CU vs MCI/mild DEM | Low Norm, Raw or Both | Any 3 | 79 | 410 | 1721 | 39 | 0.67 (0.58, 0.75) | 0.81 (0.79, 0.82) |
| CU vs MCI/mild DEM | Low Raw | 1. >55.3 raw | 70 | 271 | 1860 | 48 | 0.59 (0.50, 0.68) | 0.87 (0.86, 0.89) |
| CU vs MCI/mild DEM | Low Norm | 2. <40 T | 66 | 325 | 1806 | 52 | 0.56 (0.46, 0.65) | 0.85 (0.83, 0.86) |
| CU vs MCI/mild DEM | Low Norm and Low Raw | 3. Both | 57 | 186 | 1945 | 61 | 0.48 (0.39, 0.58) | 0.91 (0.90, 0.92) |
| CU vs MCI | Low Norm, Raw or Both | Any 3 | 62 | 410 | 1721 | 34 | 0.65 (0.54, 0.74) | 0.81 (0.79, 0.82) |
| CU vs MCI | Low Raw | 1. >55.3 raw | 56 | 271 | 1860 | 40 | 0.58 (0.48, 0.68) | 0.87 (0.86, 0.89) |
| CU vs MCI | Low Norm | 2. <40 T | 50 | 325 | 1806 | 46 | 0.52 (0.42, 0.62) | 0.85 (0.83, 0.86) |
| CU vs MCI | Low Norm and Low Raw | 3. Both | 44 | 186 | 1945 | 52 | 0.46 (0.36, 0.56) | 0.91 (0.90, 0.92) |
| CU vs dementia | Low Norm, Raw or Both | Any 3 | 17 | 410 | 1721 | 5 | 0.77 (0.55, 0.92) | 0.81 (0.79, 0.82) |
| CU vs dementia | Low Raw | 1. >55.3 raw | 14 | 271 | 1860 | 8 | 0.64 (0.41, 0.83) | 0.87 (0.86, 0.89) |
| CU vs dementia | Low Norm | 2. <40 T | 16 | 325 | 1806 | 6 | 0.73 (0.50, 0.89) | 0.85 (0.83, 0.86) |
| CU vs dementia | Low Norm and Low Raw | 3. Both | 13 | 186 | 1945 | 9 | 0.59 (0.36, 0.79) | 0.91 (0.90, 0.92) |
| CU A- vs MCI/mild DEM A+ | Low Norm, Raw or Both | Any 3 | 34 | 134 | 702 | 17 | 0.67 (0.52, 0.79) | 0.84 (0.81, 0.86) |
| CU A- vs MCI/mild DEM A+ | Low Raw | 1. >55.3 raw | 31 | 68 | 768 | 20 | 0.61 (0.46, 0.74) | 0.92 (0.90, 0.94) |
| CU A- vs MCI/mild DEM A+ | Low Norm | 2. <40 T | 30 | 119 | 717 | 21 | 0.59 (0.44, 0.72) | 0.86 (0.83, 0.88) |
| CU A- vs MCI/mild DEM A+ | Low Norm and Low Raw | 3. Both | 27 | 53 | 783 | 24 | 0.53 (0.38, 0.67) | 0.94 (0.92, 0.95) |
| A-T- vs A+T+ | Low Norm, Raw or Both | Any 3 | 46 | 136 | 654 | 69 | 0.40 (0.31, 0.50) | 0.83 (0.80, 0.85) |
| A-T- vs A+T+ | Low Raw | 1. >55.3 raw | 41 | 70 | 720 | 74 | 0.36 (0.27, 0.45) | 0.91 (0.89, 0.93) |
| A-T- vs A+T+ | Low Norm | 2. <40 T | 37 | 123 | 667 | 78 | 0.32 (0.24, 0.42) | 0.84 (0.82, 0.87) |
| A-T- vs A+T+ | Low Norm and Low Raw | 3. Both | 32 | 57 | 733 | 83 | 0.28 (0.20, 0.37) | 0.93 (0.91, 0.94) |
| CU A-T- vs CU A+T+ | Low Norm, Raw or Both | Any 3 | 18 | 121 | 643 | 62 | 0.23 (0.14, 0.33) | 0.84 (0.81, 0.87) |
| CU A-T- vs CU A+T+ | Low Raw | 1. >55.3 raw | 16 | 59 | 705 | 64 | 0.20 (0.12, 0.30) | 0.92 (0.90, 0.94) |
| CU A-T- vs CU A+T+ | Low Norm | 2. <40 T | 13 | 109 | 655 | 67 | 0.16 (0.09, 0.26) | 0.86 (0.83, 0.88) |
| CU A-T- vs CU A+T+ | Low Norm and Low Raw | 3. Both | 11 | 47 | 717 | 69 | 0.14 (0.07, 0.23) | 0.94 (0.92, 0.95) |
| A- vs A+ | Low Norm, Raw or Both | Any 3 | 102 | 150 | 715 | 264 | 0.28 (0.23, 0.33) | 0.83 (0.80, 0.85) |
| A- vs A+ | Low Raw | 1. >55.3 raw | 89 | 80 | 785 | 277 | 0.24 (0.20, 0.29) | 0.91 (0.89, 0.93) |
| A- vs A+ | Low Norm | 2. <40 T | 81 | 134 | 731 | 285 | 0.22 (0.18, 0.27) | 0.85 (0.82, 0.87) |
| A- vs A+ | Low Norm and Low Raw | 3. Both | 68 | 64 | 801 | 298 | 0.19 (0.15, 0.23) | 0.93 (0.91, 0.94) |
| CU A- vs CU A+ | Low Norm, Raw or Both | Any 3 | 68 | 134 | 702 | 247 | 0.22 (0.17, 0.27) | 0.84 (0.81, 0.86) |
| CU A- vs CU A+ | Low Raw | 1. >55.3 raw | 58 | 68 | 768 | 257 | 0.18 (0.14, 0.23) | 0.92 (0.90, 0.94) |
| CU A- vs CU A+ | Low Norm | 2. <40 T | 51 | 119 | 717 | 264 | 0.16 (0.12, 0.21) | 0.86 (0.83, 0.88) |
| CU A- vs CU A+ | Low Norm and Low Raw | 3. Both | 41 | 53 | 783 | 274 | 0.13 (0.10, 0.17) | 0.93 (0.92, 0.95) |

*Note.* The SYM Middle 2 average seconds score excludes highest and lowest trial times. Excluding highest trial time increases robustness to interruption. Normal = SYM Middle 2 average seconds ≤ 55.3 **and** SYM Middle 2 average seconds ASE T-score 40+; Low Norm = SYM Middle 2 average seconds ≤ 55.3 but SYM Middle 2 average seconds ASE T-score <40; Low Raw = SYM Middle 2 average seconds T-Score 40+ but SYM Middle 2 average seconds > 55.3, corresponds to unadjusted scaled score < 7 in the normative sample; Both Abnormal = SYM Middle 2 average seconds > 55.3 + **and** SYM Middle 2 average seconds ASE T-score <40; Any 3 = either low norm, low raw, or both are met; same as requiring either low raw or low norm; A- = amyloid PET negative; A+ = amyloid PET positive; CI = confidence interval; CU = cognitively unimpaired; DEM = dementia; FN = false negative; FP = false positive; MCI = mild cognitive impairment; MTD = Mayo Test Drive; T- = tau PET negative; T+ = tau PET positive; TN = true negative; TP = true positive. Table used with permission of Mayo Foundation for Medical Education and Research; all rights reserved.

**Table S16**. Classification accuracy statistics for combining raw score and normative score cutoffs: **SYM Accuracy (total correct)**.

| Group | Cutoff type | Cutoff | TP | FP | TN | FN | Sensitivity (95% CI) | Specificity (95% CI) |
|---|---|---|---|---|---|---|---|---|
| CU vs MCI/mild DEM | Low Norm, Raw or Both | Any 3 | 39 | 425 | 1706 | 79 | 0.33 (0.25, 0.42) | 0.80 (0.78, 0.82) |
| CU vs MCI/mild DEM | Low Raw | 1. <46 raw | 39 | 424 | 1707 | 79 | 0.33 (0.25, 0.42) | 0.80 (0.78, 0.82) |
| CU vs MCI/mild DEM | Low Norm | 2. <40 T | 33 | 401 | 1730 | 85 | 0.28 (0.20, 0.37) | 0.81 (0.79, 0.83) |
| CU vs MCI/mild DEM | Low Norm and Low Raw | 3. Both | 33 | 400 | 1731 | 85 | 0.28 (0.20, 0.37) | 0.81 (0.80, 0.83) |
| CU vs MCI | Low Norm, Raw or Both | Any 3 | 30 | 425 | 1706 | 66 | 0.31 (0.22, 0.42) | 0.80 (0.78, 0.82) |
| CU vs MCI | Low Raw | 1. <46 raw | 30 | 424 | 1707 | 66 | 0.31 (0.22, 0.42) | 0.80 (0.78, 0.82) |
| CU vs MCI | Low Norm | 2. <40 T | 24 | 401 | 1730 | 72 | 0.25 (0.17, 0.35) | 0.81 (0.79, 0.83) |
| CU vs MCI | Low Norm and Low Raw | 3. Both | 24 | 400 | 1731 | 72 | 0.25 (0.17, 0.35) | 0.81 (0.80, 0.83) |
| CU vs dementia | Low Norm, Raw or Both | Any 3 | 9 | 425 | 1706 | 13 | 0.41 (0.21, 0.64) | 0.80 (0.78, 0.82) |
| CU vs dementia | Low Raw | 1. <46 raw | 9 | 424 | 1707 | 13 | 0.41 (0.21, 0.64) | 0.80 (0.78, 0.82) |
| CU vs dementia | Low Norm | 2. <40 T | 9 | 401 | 1730 | 13 | 0.41 (0.21, 0.64) | 0.81 (0.79, 0.83) |
| CU vs dementia | Low Norm and Low Raw | 3. Both | 9 | 400 | 1731 | 13 | 0.41 (0.21, 0.64) | 0.81 (0.80, 0.83) |
| CU A- vs MCI/mild DEM A+ | Low Norm, Raw or Both | Any 3 | 17 | 151 | 685 | 34 | 0.33 (0.21, 0.48) | 0.82 (0.79, 0.84) |
| CU A- vs MCI/mild DEM A+ | Low Raw | 1. <46 raw | 17 | 151 | 685 | 34 | 0.33 (0.21, 0.48) | 0.82 (0.79, 0.84) |
| CU A- vs MCI/mild DEM A+ | Low Norm | 2. <40 T | 16 | 146 | 690 | 35 | 0.31 (0.19, 0.46) | 0.83 (0.80, 0.85) |
| CU A- vs MCI/mild DEM A+ | Low Norm and Low Raw | 3. Both | 16 | 146 | 690 | 35 | 0.31 (0.19, 0.46) | 0.83 (0.80, 0.85) |
| A-T- vs A+T+ | Low Norm, Raw or Both | Any 3 | 28 | 145 | 645 | 87 | 0.24 (0.17, 0.33) | 0.82 (0.79, 0.84) |
| A-T- vs A+T+ | Low Raw | 1. <46 raw | 28 | 145 | 645 | 87 | 0.24 (0.17, 0.33) | 0.82 (0.79, 0.84) |
| A-T- vs A+T+ | Low Norm | 2. <40 T | 27 | 141 | 649 | 88 | 0.24 (0.16, 0.32) | 0.82 (0.79, 0.85) |
| A-T- vs A+T+ | Low Norm and Low Raw | 3. Both | 27 | 141 | 649 | 88 | 0.24 (0.16, 0.32) | 0.82 (0.79, 0.85) |
| CU A-T- vs CU A+T+ | Low Norm, Raw or Both | Any 3 | 18 | 137 | 627 | 62 | 0.23 (0.14, 0.33) | 0.82 (0.79, 0.85) |
| CU A-T- vs CU A+T+ | Low Raw | 1. <46 raw | 18 | 137 | 627 | 62 | 0.23 (0.14, 0.33) | 0.82 (0.79, 0.85) |
| CU A-T- vs CU A+T+ | Low Norm | 2. <40 T | 17 | 133 | 631 | 63 | 0.21 (0.13, 0.32) | 0.83 (0.80, 0.85) |
| CU A-T- vs CU A+T+ | Low Norm and Low Raw | 3. Both | 17 | 133 | 631 | 63 | 0.21 (0.13, 0.32) | 0.83 (0.80, 0.85) |
| A- vs A+ | Low Norm, Raw or Both | Any 3 | 79 | 159 | 706 | 287 | 0.22 (0.17, 0.26) | 0.82 (0.79, 0.84) |
| A- vs A+ | Low Raw | 1. <46 raw | 79 | 159 | 706 | 287 | 0.22 (0.17, 0.26) | 0.82 (0.79, 0.84) |
| A- vs A+ | Low Norm | 2. <40 T | 70 | 154 | 711 | 296 | 0.19 (0.15, 0.24) | 0.82 (0.79, 0.85) |
| A- vs A+ | Low Norm and Low Raw | 3. Both | 70 | 154 | 711 | 296 | 0.19 (0.15, 0.24) | 0.82 (0.79, 0.85) |
| CU A- vs CU A+ | Low Norm, Raw or Both | Any 3 | 62 | 151 | 685 | 253 | 0.20 (0.15, 0.25) | 0.82 (0.79, 0.84) |
| CU A- vs CU A+ | Low Raw | 1. <46 raw | 62 | 151 | 685 | 253 | 0.20 (0.15, 0.25) | 0.82 (0.79, 0.84) |
| CU A- vs CU A+ | Low Norm | 2. <40 T | 54 | 146 | 690 | 261 | 0.18 (0.15, 0.20) | 0.83 (0.78, 0.87) |
| CU A- vs CU A+ | Low Norm and Low Raw | 3. Both | 54 | 146 | 690 | 261 | 0.18 (0.15, 0.20) | 0.83 (0.78, 0.87) |

*Note.* Normal = SYM total correct raw 46+ **and** SYM total correct ASE T-score 40+; Low Norm = SYM Total correct raw 46+ but SYM total correct ASE T-score <40; Low Raw = SYM total correct T-Score 40+ but SYM total correct raw < 46, this raw score corresponds to an unadjusted scaled score < 7 in the full normative sample; Both Abnormal = SYM total correct raw < 46 **and** SYM total correct ASE T-score <40; Any 3 = either low norm, low raw, or both are met; same as requiring either low raw or low norm; A- = amyloid PET negative; A+ = amyloid PET positive; CI = confidence interval; CU = cognitively unimpaired; DEM = dementia; FN = false negative; FP = false positive; MCI = mild cognitive impairment; MTD = Mayo Test Drive; T- = tau PET negative; T+ = tau PET positive; TN = true negative; TP = true positive. Table used with permission of Mayo Foundation for Medical Education and Research; all rights reserved.

**Table S17**. Classification accuracy statistics comparing one-stage and two-stage approaches for the MTD Composite raw score across several subgroups.

| Group | Classification Type | TP | IP | FN | TN | IN | FP | Sensitivity | Specificity |
|---|---|---|---|---|---|---|---|---|---|
| CU vs MCI/mild DEM | One-Stage raw score | 92 | 16 | 10 | 1433 | 371 | 327 | 0.78 (0.69, 0.85) | 0.85 (0.83, 0.86) |
| CU vs MCI/mild DEM | Two-Stage raw score | 92 | 16 | 10 | 1433 | 371 | 327 | 0.92 (0.85, 0.96) | 0.85 (0.83, 0.86) |
| CU vs MCI | One-Stage raw score | 72 | 15 | 9 | 1433 | 371 | 327 | 0.75 (0.65, 0.83) | 0.85 (0.83, 0.86) |
| CU vs MCI | Two-Stage raw score | 72 | 15 | 9 | 1433 | 371 | 327 | 0.91 (0.83, 0.96) | 0.85 (0.83, 0.86) |
| CU vs DEM | One-Stage raw score | 20 | 1 | 1 | 1433 | 371 | 327 | 0.91 (0.71, 0.99) | 0.85 (0.83, 0.86) |
| CU vs DEM | Two-Stage raw score | 20 | 1 | 1 | 1433 | 371 | 327 | 0.95 (0.77, 1.00) | 0.85 (0.83, 0.86) |
| CU A- vs MCI/mDEM A+ | One-Stage raw score | 42 | 7 | 2 | 619 | 130 | 87 | 0.82 (0.69, 0.92) | 0.90 (0.87, 0.92) |
| CU A- vs MCI/mDEM A+ | Two-Stage raw score | 42 | 7 | 2 | 619 | 130 | 87 | 0.96 (0.87, 1.00) | 0.90 (0.87, 0.92) |
| A-T- vs A+T+ | One-Stage raw score | 54 | 22 | 39 | 576 | 121 | 93 | 0.47 (0.38, 0.56) | 0.88 (0.86, 0.90) |
| A-T- vs A+T+ | Two-Stage raw score | 54 | 22 | 39 | 576 | 121 | 93 | 0.66 (0.57, 0.75) | 0.88 (0.86, 0.90) |
| CU A-T- vs CU A+T+ | One-Stage raw score | 23 | 18 | 39 | 572 | 115 | 77 | 0.29 (0.19, 0.40) | 0.90 (0.88, 0.92) |
| CU A-T- vs CU A+T+ | Two-Stage raw score | 23 | 18 | 39 | 572 | 115 | 77 | 0.51 (0.40, 0.63) | 0.90 (0.88, 0.92) |
| A- vs A+ | One-Stage raw score | 119 | 71 | 176 | 623 | 136 | 106 | 0.33 (0.28, 0.38) | 0.88 (0.85, 0.90) |
| A- vs A+ | Two-Stage raw score | 119 | 71 | 176 | 623 | 136 | 106 | 0.52 (0.47, 0.57) | 0.88 (0.85, 0.90) |
| CU A- vs CU A+ | One-Stage raw score | 77 | 64 | 174 | 619 | 130 | 87 | 0.24 (0.20, 0.30) | 0.90 (0.87, 0.92) |
| CU A- vs CU A+ | Two-Stage raw score | 77 | 64 | 174 | 619 | 130 | 87 | 0.45 (0.39, 0.50) | 0.90 (0.87, 0.92) |

*Note*. Some One-Stage and Two-Stage sensitivity and specificity reported in Table 3 is repeated here for ease of viewing all information in one table alongside additional subgroup comparisons and including sample sizes for true positive, indeterminate positive, false negative, true negative, indeterminate negative and false positive cases. One-stage and two-stage sensitivity and specificity include all participants in groups of interest. One-Stage Sensitivity = TP / (TP + IP + FN), Two-Stage sensitivity = (TP + IP) / (TP + IP + FN), One-Stage Specificity = TN / (TN + IN + FP), Two-Stage specificity = (TN + IN) / (TN + IN + FP). Table used with permission of Mayo Foundation for Medical Education and Research; all rights reserved. Confusion matrices were based off the following cutoffs:

- MTD raw composite raw scores >= 100 were categorized as belonging to a "normal" or control population
- MTD raw composite raw scores < 85 were categorized as having the abnormal or "positive" condition
- MTD raw composite scores between 85-99, inclusive, were not categorized (indeterminate group represented by Indeterminate Positive (IP) and Indeterminate Negative (IN) columns).
    - The indeterminate group is not excluded from any analyses. In other words, we did not calculate model performances after dropping the indeterminate group as this can lead to biased results and is according to the FDA statistically inappropriate (U.S. Food and Drug Administration. (2007). *Statistical guidance on reporting results from studies evaluating diagnostic tests: Guidance for industry and FDA staff*. U.S. Department of Health and Human Services. FDA guidance document).

**Table S18**. Combination (combo) of raw and norm score approach: classification accuracy statistics comparing one-stage and two-stage approaches for the MTD Composite interpretation guide that blends the combined raw and norm score approaches for the one-stage approach and the raw score only two-stage approach to help reduce the number of indeterminate classifications.

| Group | Classification Type | TP | IP | FN | TN | IN | FP | Sensitivity | Specificity |
|---|---|---|---|---|---|---|---|---|---|
| CU vs MCI/mild DEM | One-Stage combo approach either raw < 85 or T < 40 | 97 | 11 | 10 | 1399 | 250 | 482 | 0.82 (0.74, 0.89) | 0.77 (0.76, 0.79) |
| CU vs MCI/mild DEM | Two-Stage combo approach (+clinical eval if raw 85-99) | 97 | 11 | 10 | 1399 | 250 | 482 | 0.92 (0.85, 0.96) | 0.77 (0.76, 0.79) |
| CU vs MCI | One-Stage combo approach either raw < 85 or T < 40 | 77 | 10 | 9 | 1399 | 250 | 482 | 0.80 (0.71, 0.88) | 0.77 (0.76, 0.79) |
| CU vs MCI | Two-Stage combo approach (+clinical eval if raw 85-99) | 77 | 10 | 9 | 1399 | 250 | 482 | 0.91 (0.83, 0.96) | 0.77 (0.76, 0.79) |
| CU vs DEM | One-Stage combo approach either raw < 85 or T < 40 | 20 | 1 | 1 | 1399 | 250 | 482 | 0.91 (0.71, 0.99) | 0.77 (0.76, 0.79) |
| CU vs DEM | Two-Stage combo approach (+clinical eval if raw 85-99) | 20 | 1 | 1 | 1399 | 250 | 482 | 0.95 (0.77, 1.00) | 0.77 (0.76, 0.79) |
| CU A- vs MCI/mDEM A+ | One-Stage combo approach either raw < 85 or T < 40 | 43 | 6 | 2 | 599 | 79 | 158 | 0.84 (0.71, 0.93) | 0.81 (0.78, 0.84) |
| CU A- vs MCI/mDEM A+ | Two-Stage combo approach (+clinical eval & amyloid PET if raw 85-99) | 43 | 6 | 2 | 599 | 79 | 158 | 0.96 (0.87, 1.00) | 0.81 (0.78, 0.84) |
| A-T- vs A+T+ | One-Stage combo approach either raw < 85 or T < 40 | 62 | 14 | 39 | 557 | 71 | 162 | 0.54 (0.44, 0.63) | 0.79 (0.77, 0.82) |
| A-T- vs A+T+ | Two-Stage combo approach (+amyloid & tau PET if raw 85-99) | 62 | 14 | 39 | 557 | 71 | 162 | 0.66 (0.57, 0.75) | 0.79 (0.77, 0.82) |
| CU A-T- vs CU A+T+ | One-Stage combo approach either raw < 85 or T < 40 | 30 | 11 | 39 | 553 | 66 | 145 | 0.38 (0.27, 0.49) | 0.81 (0.78, 0.84) |
| CU A-T- vs CU A+T+ | Two-Stage combo approach (+amyloid & tau PET if raw 85-99) | 30 | 11 | 39 | 553 | 66 | 145 | 0.51 (0.40, 0.63) | 0.81 (0.78, 0.84) |
| A- vs A+ | One-Stage combo approach either raw < 85 or T < 40 | 138 | 56 | 172 | 603 | 84 | 178 | 0.38 (0.33, 0.43) | 0.79 (0.77, 0.82) |
| A- vs A+ | Two-Stage combo approach (+amyloid PET if raw 85-99) | 138 | 56 | 172 | 603 | 84 | 178 | 0.53 (0.48, 0.58) | 0.79 (0.77, 0.82) |
| CU A- vs CU A+ | One-Stage combo approach either raw < 85 or T < 40 | 95 | 50 | 170 | 599 | 79 | 158 | 0.30 (0.25, 0.36) | 0.81 (0.78, 0.84) |
| CU A- vs CU A+ | Two-Stage combo approach (+amyloid PET if raw 85-99) | 95 | 50 | 170 | 599 | 79 | 158 | 0.46 (0.40, 0.52) | 0.81 (0.78, 0.84) |

*Note*. Some One-Stage and Two-Stage sensitivity and specificity reported in Table 3 is repeated here for ease of viewing all information in one table alongside additional subgroup comparisons and including sample sizes for true positive, indeterminate positive, false negative, true negative, indeterminate negative and false positive cases. One-stage and two-stage sensitivity and specificity include all participants in groups of interest. One-Stage Sensitivity = TP / (TP + IP + FN), Two-Stage sensitivity = (TP + IP) / (TP + IP + FN), One-Stage Specificity = TN / (TN + IN + FP), Two-Stage specificity = (TN + IN) / (TN + IN + FP). T-scores adjusted for age, sex, and education. Table used with permission of Mayo Foundation for Medical Education and Research; all rights reserved. Confusion matrices were based off the following cutoffs:

1. MTD raw composite raw scores >= 100 AND MTD composite T-scores >= 40 were categorized as belonging to a "normal" or control population
2. MTD raw composite raw scores < 85 were categorized as having the abnormal or "positive" condition
3. MTD composite T-scores < 40 were categorized as having the abnormal or "positive" condition
4. MTD raw composite raw scores between 85-99 (AND not classified as positive by the normative score cutoff), inclusive, were not categorized (indeterminate group represented by Indeterminate Positive (IP) and Indeterminate Negative (IN) columns. Because some cases are classified as positive by the normative score cutoff, this reduces the number of indeterminate cases relative to the raw score only two-stage classification approach.
    - The indeterminate group is not excluded from any analyses. In other words, we did not calculate model performances after dropping the indeterminate group as this can lead to biased results and is according to the FDA statistically inappropriate (U.S. Food and Drug Administration. (2007). *Statistical guidance on reporting results from studies evaluating diagnostic tests: Guidance for industry and FDA staff*. U.S. Department of Health and Human Services. FDA guidance document).

cc BY SA

### STARD Checklist

This study is reported in accordance with Standards for Reporting of Diagnostic Accuracy Studies (STARD); dementia-specific reporting considerations were addressed in accordance with the STARDdem Initiative. The checklist below combines the STARD 2015 items with applicable STARDdem considerations.

Noel-Storr AH, McCleery JM, Richard E, Ritchie CW, Flicker L, Cullum SJ, et al. Reporting standards for studies of diagnostic test accuracy in dementia: The STARDdem Initiative. Neurology. 2014;83:364–73.

Bossuyt PM, Reitsma JB, Bruns DE, Gatsonis CA, Glasziou PP, Irwig L, et al. STARD 2015: an updated list of essential items for reporting diagnostic accuracy studies. BMJ. 2015;351:h5527.

http://www.equator-network.org/reporting-guidelines/stard

| Section & Topic | No | Item | Reported on page # |
|---|---|---|---|
| **TITLE OR ABSTRACT** | | | |
| | **1** | Identification as a study of diagnostic accuracy using at least one measure of accuracy<br>(such as sensitivity, specificity, predictive values, or AUC)<br><br>STARDdem: studies reporting a sensitivity/specificity or 2x2 data derivable fall within the scope of STARDdem and should be indexed accordingly | p. 1 (title) and p. 2 (key words) |
| **ABSTRACT** | | | |
| | **2** | Structured summary of study design, methods, results, and conclusions<br>(for specific guidance, see STARD for Abstracts) | p. 2 (Abstract) |
| **INTRODUCTION** | | | |
| | **3** | Scientific and clinical background, including the intended use and clinical role of the index test | p. 3-4 (Introduction) |
| | **4** | Study objectives and hypotheses<br><br>STARDdem: State the research questions or study aims such as estimating diagnostic accuracy or comparing accuracy between tests or across participant groups. Report test purpose: “stand-alone” test or as an addition to other tests or clinical criteria | p. 4-5 (Introduction), p. 9 (Methods) |
| **METHODS** | | | |
| *Study design* | **5** | Whether data collection was planned before the index test and reference standard were performed (prospective study) or after (retrospective study)<br><br>STARDdem: Authors should report the timing of the analysis plan regarding data collection: Was the analysis plan set out in a protocol before index and reference standards were performed? If not, when was the analysis plan created? | p. 2 (Abstract), p. 6 (Methods), Supplemental Methods (Online Supplemental Material) |
| *Participants* | **6** | Eligibility (exclusion/inclusion) criteria<br><br>STARDdem: Key inclusion criteria: (1) demographic, especially age; (2) cognition- or disease-related criteria. Accurate description of the target sample is required including reporting criteria used to define the study population. Report referral pathways, precise locations of patient | p. 9 (Inclusion Criteria) |

CC BY SA

| | | | |
|---|---|---|---|
| | | recruitment, where index test and reference standard were performed. For secondary/tertiary settings, helpful to report the medical subspecialty or hospital department (e.g., psychiatry, neurology). Diagnostic accuracy studies in dementia are often nested within larger cohort studies. If this is the case, then the targeted population for the cohort study and the method of selection into the cohort should be described and/or the parent study cited | |
| | **7** | On what basis potentially eligible participants were identified (such as symptoms, results from previous tests, inclusion in registry)<br><br>STARDdem: Report whether those in intermediate categories (e.g., possible AD or possible DLB) were excluded | p. 9 (Inclusion Criteria), p. 11-12 (Results) |
| | **8** | Where and when potentially eligible participants were identified (setting, location and dates)<br><br>STARDdem: Pertinent particularly to longitudinal (delayed verification) studies, authors should report recruitment dates of the study (not to be confused with recruitment dates of the wider cohort study from which it might be drawn), and<br>the beginning (first participant) and end (last participant) dates of the periods during which the index test(s) and reference standard were performed. | p. 5-6 for setting and location (Method, Participants and Study Design; Table 1 note); p. 9 for dates (Method). |
| | **9** | Whether participants formed a consecutive, random or convenience series<br><br>STARDdem: Participant sampling: Was the study population a consecutive series of participants defined by the selection criteria in items 3 and 4? If not, specify how participants were further selected. | p. 5-6 (Participants and Study Design) |
| *Test methods* | **10a** | Index test, in sufficient detail to allow replication | p. 7-8 (Remote Digital Cognitive Assessment) |
| | **10b** | Reference standard, in sufficient detail to allow replication<br><br>STARDdem: For neuropathologic and clinical reference standards, the diagnostic criteria used should be specified. Where relevant, reference should be made to studies validating the criteria. Report whether standard consensus clinical criteria<br>incorporate the index test (incorporation bias rendering blinding of index test impossible) | p. 6-7 (Neuroimaging Biomarkers, Person-Administered Neuropsychological, Mental Status Screening Measures, and clinical diagnosis criteria) |
| | **11** | Rationale for choosing the reference standard (if alternatives exist) | p. 3-5 (Introduction) |
| | **12a** | Definition of and rationale for test positivity cut-offs or result categories of the index test, distinguishing pre-specified from exploratory | p. 9-10 (Statistical Methods), Table 3. |
| | **12b** | Definition of and rationale for test positivity cut-offs or result categories of the reference standard, distinguishing pre-specified from exploratory | p. 6 (Clinical Characterization, Neuroimaging Biomarkers) |
| | **13a** | Whether clinical information and reference standard results were available to the performers/readers of the index test | p. 6-7 (Clinical Characterization) |
| | **13b** | Whether clinical information and index test results were available to the assessors of the reference standard | p. 6-7 (Clinical Characterization, Neuroimaging biomarkers) |
| | ** | STARDdem: the number, training, and expertise of the person executing and reading the index tests and the reference standard.<br><br>Specify details of administration, which version. Clinical diagnostic criteria: what information was available to inform the diagnoses; how the criteria were applied (e.g., by individual clinicians, by consensus conference, by semiautomated algorithm). Imaging and laboratory tests: specify materials and instruments, including sample handling and concordance with any | p. 6-8 (Clinical Characterization; Neuroimaging Biomarkers; Person-Administered Neuropsychological…; Remote Digital Cognitive Assessment) |

cc BY SA

| | | | |
|---|---|---|---|
| | | harmonization criteria. In new assays, describe all steps in detail. Any particular preparation of participants should be described | |
| | ** | STARDdem: Methods for calculating test reproducibiity if done<br><br>Applies to the reference standard as well as to the index test. Both should be reported/adequately referenced. Report interrater and test-retest reliability of reference standard as established in the study being reported, rather than simply referring to other studies in which reproducibility has been established. The training that image readers receive should be carefully described. Studies in which the accuracy of "majority" judgments are reported should also report data for the minority judgments. Reports of the impact of training should clearly describe the characteristics of the sample used for training and whether it is representative of the group to which the test will be applied | Not assessed in the current diagnostic-accuracy analysis but test-retest reliability of the index test was recently reported in detail in an overlapping sample, as described on p. 8. |
| *Analysis* | **14** | Methods for estimating or comparing measures of diagnostic accuracy | p. 9-11 (Statistical Methods) |
| | **15** | How indeterminate index test or reference standard results were handled | p. 7 (Neuroimaging biomarkers), p. 9 (Inclusion Criteria), p. 9-12 (Statistical Method, Participant Characteristics); Tables 3, S17, S18 |
| | **16** | How missing data on the index test and reference standard were handled | p. 9 (Inclusion Criteria), p. 9-12 (Statistical Method, Participant Characteristics). P. 13-14 (Results, Comparison with traditional in-person cognitive measures) |
| | **17** | Any analyses of variability in diagnostic accuracy, distinguishing pre-specified from exploratory | p. 4-5, p. 9-12 (Statistical Method), p. 12-14 (Results), Table 2, Supplementary tables. |
| | **18** | Intended sample size and how it was determined | Supplemental methods |
| **RESULTS** | | | |
| *Participants* | **19** | Flow of participants, using a diagram | Figure S1 |
| | **20** | Baseline demographic and clinical characteristics of participants | Table 1, p. 11-12 |
| | **21a** | Distribution of severity of disease in those with the target condition | Table 1, p. 11-12 |
| | **21b** | Distribution of alternative diagnoses in those without the target condition | Not systematically characterized because the primary reference-negative group was defined as cognitively unimpaired rather than by alternate clinical diagnoses. However, p. 12 reports the percentage of participants who are A+, for those with amyloid PET available. Table 1 also reports MCI subtype information. |
| | **22** | Time interval and any clinical interventions between index test and reference standard | No clinical interventions were part of this study. Other potential non-study clinical treatments that could have occurred were not tracked. Time intervals between MTD and the in-person visit and between MTD and PET imaging are provided in Table 1. |
| *Test results* | **23** | Cross tabulation of the index test results (or their distribution) by the results of the reference standard | Supplementary tables |
| | **24** | Estimates of diagnostic accuracy and their precision (such as 95% confidence intervals) | Table 2, Figure 1, Figure 2 |
| | **25** | Any adverse events from performing the index test or the reference standard | Supplemental Results |
| **DISCUSSION** | | | |
| | **26** | Study limitations, including sources of potential bias, statistical uncertainty, and generalisability | p. 19-22 |

| | | | |
|---|---|---|---|
| | **27** | Implications for practice, including the intended use and clinical role of the index test<br><br>STARDdem: Discuss differences in age and comorbidity between the study population and the patients typically seen in clinical practice. Discuss whether the reported data demonstrate “added” or “incremental” value of the index test over and above other routine diagnostic tests. Identify stage of development of the test (e.g., proof of concept; defining accuracy in a typical spectrum of patients). Discuss the further research needed to be done to make test applicable to population in whom likely to be applied in practice | p.20-22 |
| **OTHER INFORMATION** | | | |
| | **28** | Registration number and name of registry | This diagnostic accuracy study was not registered in a clinical study registry. |
| | **29** | Where the full study protocol can be accessed | The full study protocol is not publicly available but other manuscripts have described study procedures in detail, particularly Patel. et al., 2024 and Hughes et al., 2026. The prospectively specified aims and analysis framework are described in the Supplemental Methods, and the funded project is listed in NIH Reporter. |
| | **30** | Sources of funding and other support; role of funders | p.23 (Acknowledgements). The funders had no role in study design, data collection, analysis, interpretation, manuscript preparation, or the decision to submit the work. |

CC BY SA
